# Time-resolved predictability of end-of-therapy outcome and relapse after cure in Phase 3 tuberculosis trials

**DOI:** 10.64898/2026.09.15.26362605

**Authors:** Daniel Garger, Jamison H. Burks, Nicholas I. Paton, Derek J. Sloan, Gareth Maher-Edwards, Fabian Theis, Michael P. Menden, Francesco Paolo Casale

**Affiliations:** Institute of Computational Biology, Helmholtz Zentrum München - German Research Center for Environmental Health, Neuherberg, Germany; Department of Biology, Ludwig Maximilian University of Munich, Munich, Germany; Institute of AI for Health, Helmholtz Zentrum München - German Research Center for Environmental Health, Neuherberg, Germany; Infectious Diseases Translational Research Programme, National University of Singapore, Singapore; London School of Hygiene and Tropical Medicine, London, UK; Department of Pulmonology, Radboud University Medical Centre, Nijmegen, The Netherlands; School of Medicine, University of St Andrews, St Andrews, UK; Global Health Medicines R&D, GSK, London, UK; School of Computation, Information and Technology, Technical University of Munich, Garching, Germany; TUM School of Life Sciences, Technical University of Munich, Munich, Germany; Department of Biochemistry and Pharmacology, Bio21 Molecular Science and Biotechnology Institute, The University of Melbourne, Melbourne, VIC, Australia; Helmholtz Pioneer Campus, Helmholtz Zentrum München - German Research Center for Environmental Health, Neuherberg, Germany

## Abstract

Relapse after apparently successful tuberculosis (TB) therapy remains difficult to predict, and how relapse risk evolves throughout treatment remains unclear. Using harmonised clinical data from two Phase 3 trials (2,918 participants), we performed time-resolved prediction of end-of-therapy (EOT) outcomes and post-treatment relapse by training models using tabular data at monthly intervals from baseline to therapy end. Prediction of EOT outcomes improved after month 3 (ROC-AUC up to 0.84), driven by sputum-smear and solid culture. In contrast, relapse prediction among participants with favourable EOT outcomes and completed follow-up improved only modestly through month 3 (ROC-AUC 0.58-0.63) before declining, with age, sex, clinical symptoms and bacterial burden contributing most strongly to prediction. Models trained on large language model-derived embeddings, a more flexible representation of the same variables, matched tabular relapse models in performance throughout therapy, and outperformed tabular EOT outcome models at months 3 and 4 (ΔROC-AUC: 0.12 and 0.14), with longitudinal modelling and sparse variable inclusion only improving relapse prediction at months 4-6 (ΔROC-AUC: 0.09, 0.19 and 0.12), however with reduced interpretability. Models incorporating data after baseline provided incremental improvements in post-treatment relapse risk stratification compared with baseline cavitation and sputum smear alone (4-month relapse-free survival: 94.8%/78.5% for model-derived low/high risk groups, vs. 91.4%/82.9% for baseline easy-/hard-to-treat groups). Overall, these findings suggest that while routine clinical data collected after baseline can improve post-treatment risk stratification, it offers limited predictive value for relapse, underscoring the need for relapse-specific biomarkers.

## Introduction

Tuberculosis (TB) remains a leading infectious cause of death, with an estimated 1.23 million deaths in 2024^1^. Despite treatment success rates of 88% for drug-susceptible and 71% for drug-resistant TB, relapse after apparently successful therapy remains a major challenge, occurring at 2.26 per 100 person-years and accounting for most post-treatment recurrences^1,2^.

Participants with favourable end-of-therapy (EOT) outcomes from standard-of-care TB therapy ^3^ are typically considered cured and discharged from active care ^4^, despite relapse remaining a real and ongoing concern. A risk stratification tool for the post-treatment period could substantially improve how these participants are managed clinically ^3^. Current stratification approaches divide participants into easy-and hard-to-treat (ETT/HTT) groups using baseline bacterial burden, cavitation, or extensive pulmonary involvement, but were validated against composite unfavourable outcomes rather than relapse alone ^5,6^. Whether these strata capture relapse risk among participants who complete therapy successfully is therefore unknown, and relapse mechanisms in ETT participants - unlike HTT relapse, which is largely driven by cavitary disease - remain to be elucidated ^7,8^. Previous relapse predictions using routine clinical data have achieved only moderate predictive performance (ROC-AUC ≈ 0.6-0.8) ^5,7,9–16^, suggesting that relapse remains difficult to predict from these variables alone. This difficulty may reflect biological persistence of *Mycobacterium tuberculosis* ^17^ together with low relapse rates after treatment. It may also reflect limitations of routine monitoring variables: sputum smear microscopy has limited sensitivity, culture-based testing is contamination-prone, costly and slow, and weight gain can be confounded by food access and inconsistent calibration^18–22^.

Although previous studies have identified broadly consistent baseline predictors (including male sex, older age, high sputum smear grade, low body mass index (BMI), HIV co-infection, and cavitation) and on-treatment predictors (including two-month culture conversion and treatment adherence) associated with relapse or composite unfavourable outcomes ^5,7,9–14,23,24^, they have generally evaluated prediction at isolated time points. Consequently, it remains unclear how the prognostic information contained in routine clinical data evolves during treatment, whether longitudinal measurements meaningfully extend relapse predictability, or when clinically useful prognostic information becomes available. Emerging studies of host serum proteins further suggest that relapse-associated prognostic information may itself change over the course of treatment ^15,16^.

These observations highlight the need to understand how relapse risk prediction and variable contributions evolve during treatment, and could clarify whether current limitations primarily reflect modelling approaches or the information content of routinely collected clinical measurements. We therefore aimed to assess this evolution, evaluate whether longitudinal data, additional sparse variables, and flexible modelling approaches improve prediction, and determine whether this approach can inform post-treatment relapse risk stratification. To address this question, we therefore analysed harmonised individual-participant data from OFLOTUB ^25^ and REMoxTB ^26^, two phase 3 trials evaluating the potential for treatment shortening by comparing standard 6-month regimens to quinolone-containing 4-month regimens, sourced from TB-PACTS ^27^ (2,918 participants, **Table 1**, **Figure 1A**). Using a sliding-window framework, we quantify how the predictability of EOT outcomes and post-treatment relapse of participants with favourable EOT outcomes and follow-up completion evolves during treatment (**Figure 1B**). We further evaluate whether additional longitudinal clinical information and alternative data representations improve relapse predictability, while assessing their implications for biological interpretation and post-treatment risk stratification (**Figure 1C**). For this we train models on raw clinical data using methods capable of using single-timepoint (L1-regularised logistic regression, LR; XGBoost) or sequential data (long short-term memory, LSTM), comparing them to models trained on large language model (LLM-)derived embeddings, which allow for sparse variable inclusion and longitudinal modelling. Together, these analyses define the temporal structure and limits of tuberculosis relapse predictability from routine clinical data, motivating the development of relapse-specific biomarkers to improve post-treatment risk assessment.

**Figure 1.**
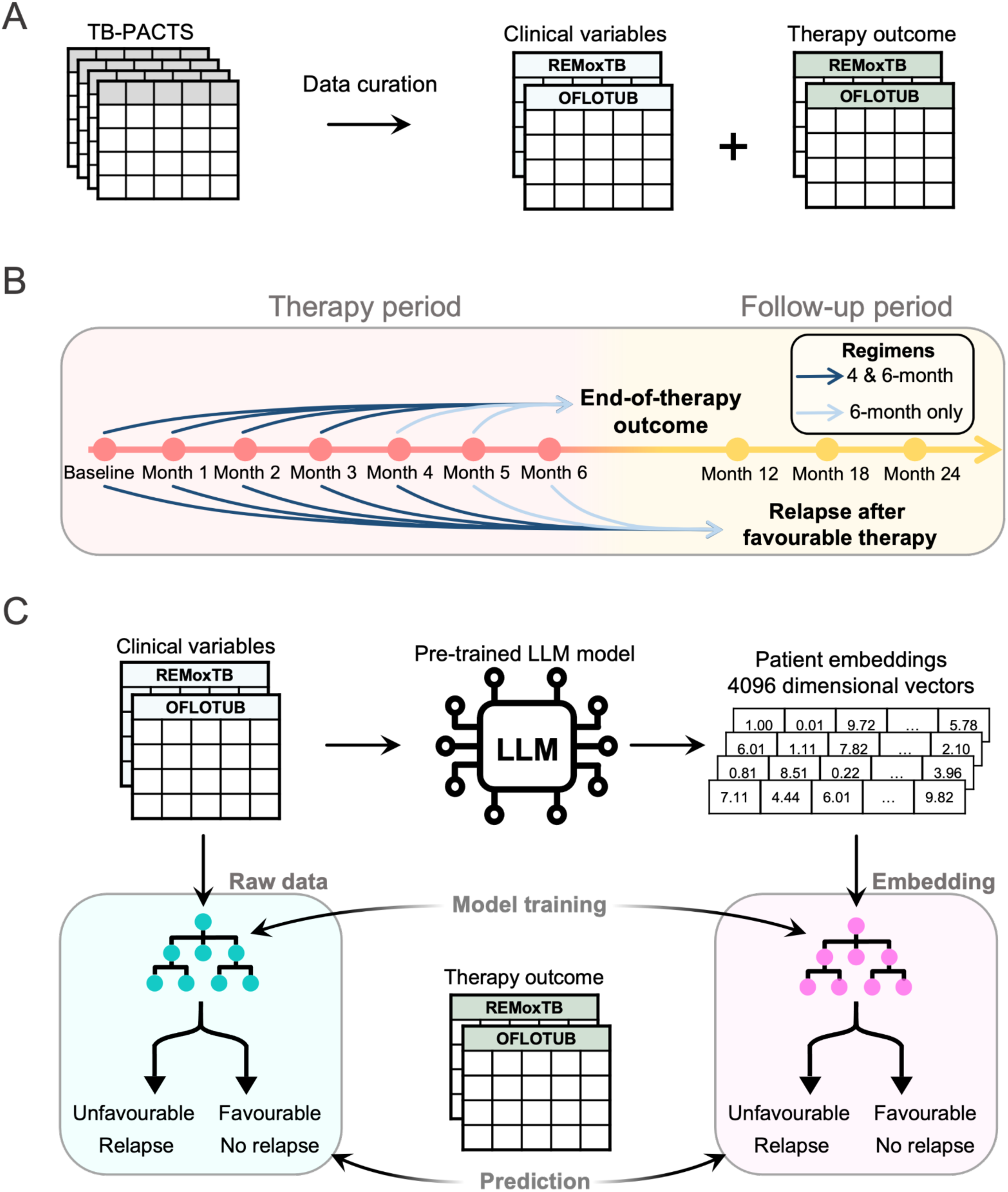
Cohort construction and time-resolved prediction framework. (**A**) Overview of cohort construction from the TB-PACTS repository. Raw clinical trial data were curated to extract aligned longitudinal clinical variables and adjudicated therapy outcomes. From the available studies, the Phase 3 REMoxTB and OFLOTUB trials were selected for analysis. (**B**) Time-resolved evaluation design using a sliding-window approach. At each monthly cutoff, separate models were trained on data available up to that timepoint, to predict end-of-therapy (EOT) outcome (top timeline) and relapse during post-treatment follow-up among participants with a favourable EOT outcome and follow-up completion (bottom timeline). Arrow colour indicates participant inclusion by regimen duration (dark blue: 4-month regimens; light blue: 6-month regimens). (**C**) Modelling representations evaluated in this study. Left: prediction using raw tabular clinical and treatment data. Right: embedding-based approach in which a pre-trained large language model (LLM) transforms participant data into fixed-length numerical embeddings, which are then used for model training and prediction for both EOT outcome and relapse.

**Table 1.** Number of participants used for analysis. Number of participants included in the analysis by study, treatment arm, and prediction task (end-of-therapy outcome and relapse following favourable therapy). Bracketed proportions reflect fractions within each arm and prediction task, the bottom row reflects overall proportions across all arms. For relapse prediction, only participants with a favourable end-of-therapy outcome and completed follow-up were considered.

|  |  | End-of-therapy outcome |  | Relapse after favourable therapy |  |
| --- | --- | --- | --- | --- | --- |
| Study | Arm | Favourable | Unfavourable | No relapse | Relapse |
| REMoxTB | 2EHRZ/4HR | 449 (97.4%) | 12 (2.6%) | 402 (96.4%) | 15 (3.6%) |
|  | 2EMRZ/2MR | 473 (95.9%) | 20 (4.1%) | 367 (82.8%) | 76 (17.2%) |
|  | 2MHRZ/2MHR | 462 (97.1%) | 14 (2.9%) | 382 (88.6%) | 49 (11.4%) |
| OFLOTUB | Control | 712 (95.0%) | 37 (5.0%) | 544 (93.5%) | 38 (6.5%) |
|  | Gatifloxacin | 723 (97.7%) | 17 (2.3%) | 536 (86.6%) | 83 (13.4%) |
|  | Sum | 2818 (96.6%) | 100 (3.4%) | 2231 (89.5%) | 261 (10.5%) |

## Results

### Time-resolved predictability of end-of-therapy outcomes and relapse

We employed a sliding-window framework in which separate models were trained at each monthly cutoff using only data available up to that point (**Figure 1B**). Models were trained on a shared variable set across trials (“common variables”; **Table 2**), with repeated stratified splits to account for class imbalance. For non-sequential models, we compared two input representations: the most recent visit before each cutoff (“last visit in period”) and concatenation of all prior visits (“last visits concatenated”). Full details are provided in the **Methods** and **Supplementary Methods**. For EOT outcome prediction, last-visit models showed stable performance from baseline to month 3, with mean ROC-AUC values ranging from 0.53 to 0.61 (**Figure 2A**). Performance improved at later cutoffs, reaching ROC-AUC values of 0.77 for LR and 0.84 for XGBoost at month 5. Concatenating prior visits produced similar trends with only marginal improvements at isolated time points (**Supplementary Figure 1A**). Within-arm performance evaluation showed comparable early performance in all treatment arms, with both 6-month regimens exhibiting similar improvements at later time points. (**Supplementary Figure 2,3**).

**Figure 2.**
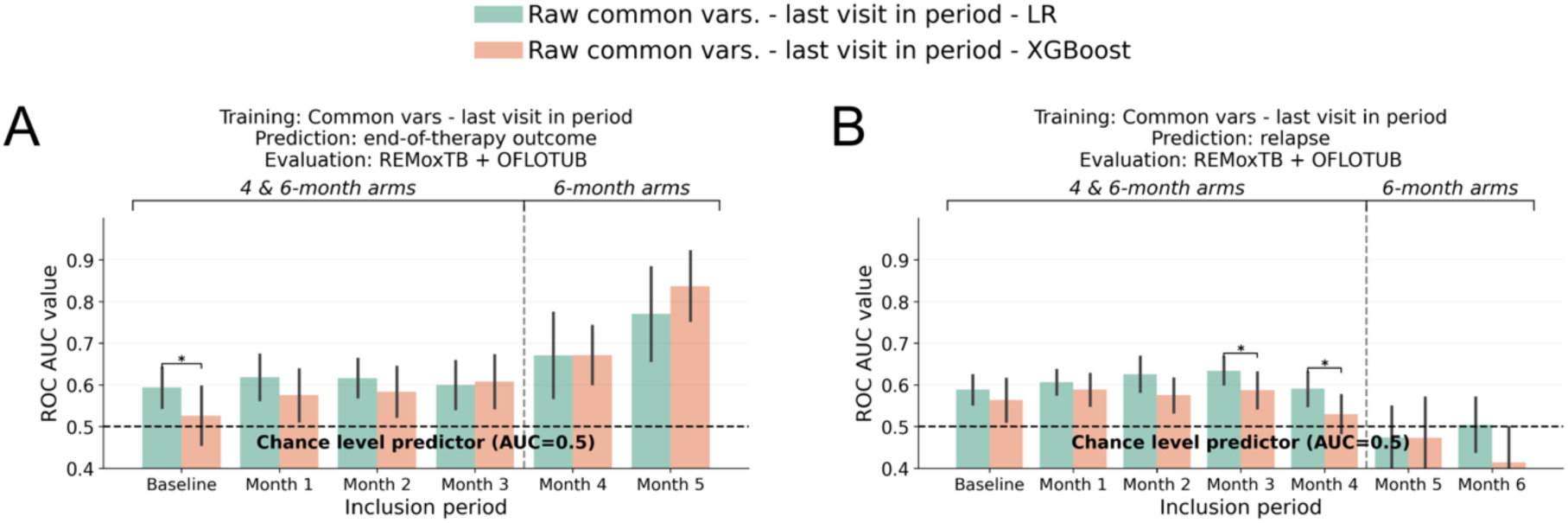
Time-resolved prediction performance using conventional clinical models. (**A**) End-of-therapy (EOT) outcome prediction and (**B**) relapse prediction using logistic regression (LR) and XGBoost models trained on raw tabular clinical and treatment data. Models were trained at monthly inclusion cutoffs from baseline to month 5 and month 4 for EOT outcome and relapse prediction respectively, using common clinical and treatment variables across studies from the most recent visit prior to each cutoff (“last visit in period”) and evaluated across the combined REMoxTB and OFLOTUB cohorts. The vertical dashed line marks where 4-month arms complete treatment and drop from the analysis, leaving 6-month arms only. Bars indicate mean predictive performance (ROC-AUC), with error bars showing the standard deviation across 25 test sets from repeated 25×5 cross-validation. The dashed horizontal line denotes chance-level performance (ROC-AUC = 0.5). Corrected resampled t-tests ^28^ were used to compare setups within periods, accounting for the correlation between performance estimates induced by overlapping test sets across the 25 random train-test splits, with only significant comparisons shown after Benjamini-Hochberg correction (*: adj. p<0.05). The dashed line indicates chance-level performance (AUC = 0.5)

**Table 2.**
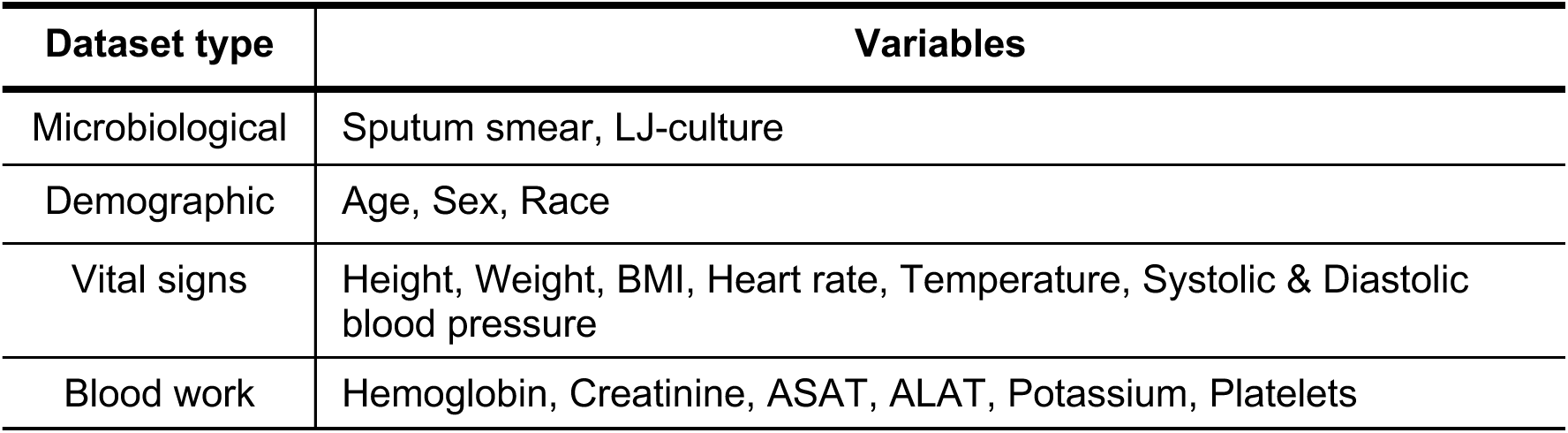
General overview of variables used for analysis. Table containing common variables selected across REMoxTB and OFLOTUB for analysis. LJ: Löwenstein-Jensen; BMI: body-mass-index; ASAT: aspartate aminotransferase; ALAT: alanine aminotransferase.

| Dataset type | Variables |
| --- | --- |
| Microbiological | Sputum smear, LJ-culture |
| Demographic | Age, Sex, Race |
| Vital signs | Height, Weight, BMI, Heart rate, Temperature, Systolic & Diastolic blood pressure |
| Blood work | Hemoglobin, Creatinine, ASAT, ALAT, Potassium, Platelets |

In contrast, relapse prediction exhibited a markedly different temporal pattern. In the better-performing LR model, discrimination improved only modestly from baseline to month 3 (mean ROC-AUC 0.58-0.63) before declining towards chance-level performance at months 5 and 6 (**Figure 2B**). This pattern was consistent across input representations (**Supplementary Figure 1B**). Within-arm evaluation showed similar temporal dynamics, with most treatment arms peaking around months 2-3 before declining. This pattern was most pronounced in the REMoxTB 2EHRZ/4HR arm, whereas the 2EMRZ/2MR arm maintained relatively stable performance throughout follow-up (**Supplementary Figure 4**).

Sequential LSTM models did not improve discrimination for either outcome, achieving mean ROC-AUC values between 0.5 and 0.6 across time points (**Supplementary Figure 1C,D**). Overall, non-sequential LR and XGBoost models consistently outperformed LSTMs, and incorporating multiple prior visits did not provide systematic gains over using only the most recent visit. Given their comparable or superior discrimination and substantially lower computational complexity, last-visit LR and XGBoost models were carried forward for subsequent analyses.

### Temporal evolution of clinical predictors of end-of-therapy outcomes and relapse

To characterise the clinical factors underlying time-resolved prediction, we computed SHAP values ^29^ from the best-performing conventional models, focusing on last-visit logistic regression models (**Supplementary Figure 5**). Across both outcomes, demographic, microbiological, laboratory, and symptomatic variables contributed to prediction, but their relative importance evolved markedly over the course of treatment.

For EOT outcome prediction, markers of bacterial burden increasingly dominated prediction as treatment progressed. Sputum-based microbiological measures, including Lowenstein-Jensen (LJ) culture status and sputum smear grade, contributed little early in therapy but increasingly shifted predictions toward Unfavourable outcomes from month 2 onward, reaching maximal influence near therapy completion (**Figure 3A**; **Supplementary Figure 6-12**). By month 5, the strongest single driver of Unfavourable outcome prediction was a high sputum smear grade.

**Figure 3.**
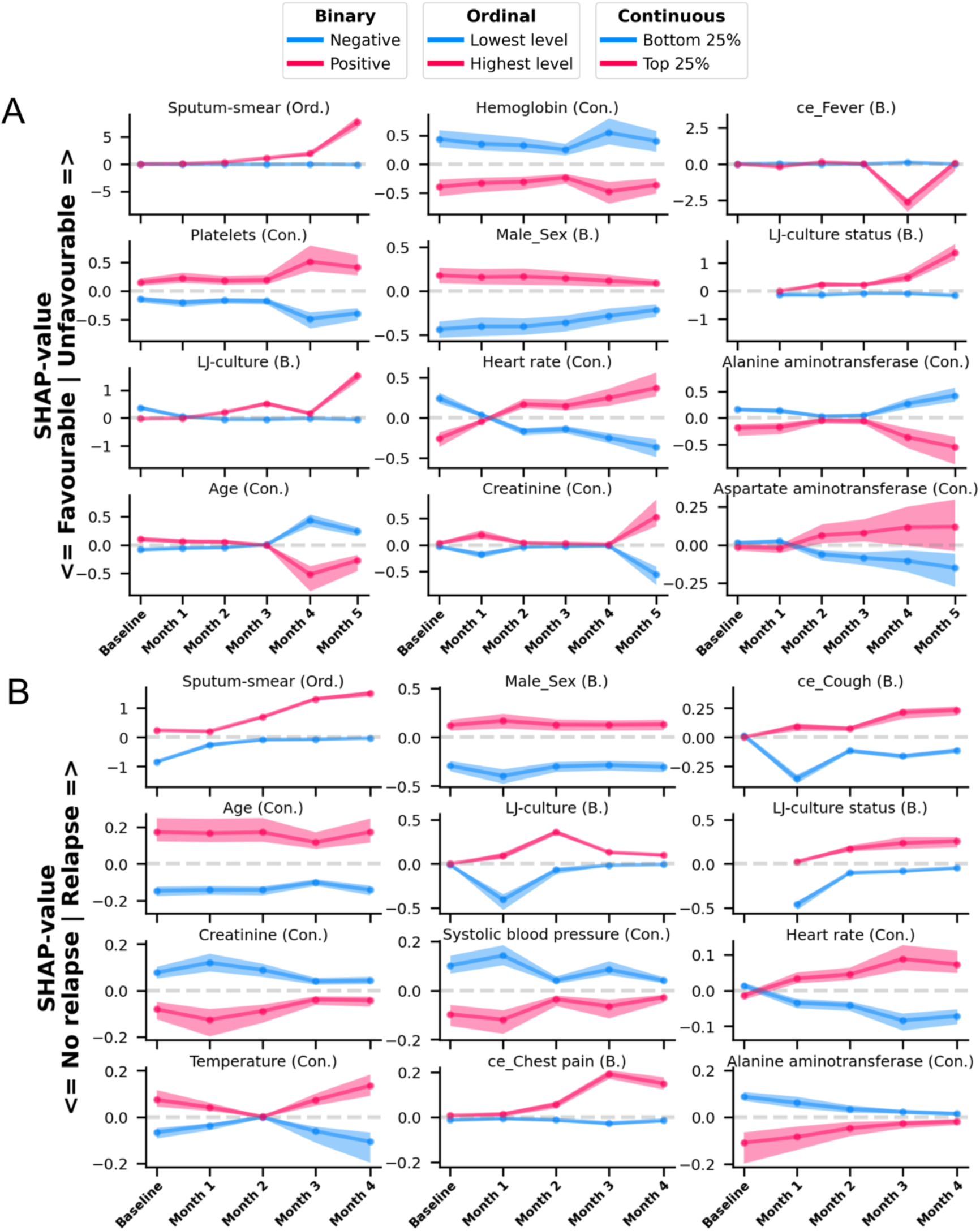
Population-level SHAP value trajectories across therapy periods for selected clinical variables. Population-level SHAP value trajectories from last-visit raw-data models for end-of-treatment (EOT) outcome prediction (A) and relapse prediction (B). Variables were selected based on consistently large extreme SHAP values across cross-validation splits and time points. For relapse prediction, time points with non-random model performance are shown (baseline-month 4, see Figure 2B). Participants were stratified by the distribution of the corresponding feature values at each time point: continuous variables were divided into quantile-based strata (bottom 25% and top 25%), while binary and ordinal variables were stratified into two groups (negative vs positive; lowest vs highest level). Lines represent the median SHAP value, and shaded bands indicate the interquartile range (25-75%) of SHAP values within each stratum at a given time point. Line and band colours encode the feature value strata (see legend). The horizontal dashed line denotes zero contribution to the model prediction. Positive SHAP values indicate increased contribution towards positive label (Unfavourable/Relapse), whereas negative values indicate decreased contribution. B.: binary; Ord.: ordinal; Con.: continuous; ce: clinical event; LJ-culture: Löwenstein-Jensen culture; BMI: body mass index.

Host-related factors exhibited more heterogeneous temporal patterns. Higher BMI was consistently associated with Favourable outcomes, whereas age, height, and weight exerted stronger influence early in therapy and attenuated or reversed later. Male sex showed a persistent association with Unfavourable outcomes. Among laboratory variables, elevated platelet counts and higher levels of creatinine and aspartate aminotransferase (ASAT) were associated with Unfavourable predictions, whereas higher hemoglobin, systolic blood pressure, and alanine aminotransferase (ALAT) were associated with Favourable outcomes. Several variables, including heart rate, potassium, and diastolic blood pressure, exhibited time-dependent reversals in attribution direction (**Supplementary Figure 6**).

In contrast to EOT prediction, relapse prediction among participants with Favourable EOT outcomes and follow-up completion exhibited a distinct temporal profile (**Supplementary Figure 13-20**). Microbiological markers increased in influence during the first months of treatment, with LJ-culture positivity and high sputum-smear grade showing the strongest associations with relapse risk between months 1 and 4 (**Figure 3B**). Host-related factors played a comparatively larger role than in EOT prediction: male sex, higher platelet counts, older age, elevated ASAT, higher temperature, and persistent symptoms, including sweating, cough, and chest pain, consistently shifted predictions toward relapse, with age and sex exhibiting relatively stable effect sizes across periods. Higher BMI and systolic blood pressure were generally associated with reduced relapse risk. Notably, elevated creatinine and ALAT levels were also associated with lower relapse risk. Potassium and diastolic blood pressure again exhibited time-dependent reversals in attribution direction, but in the opposite direction to those observed in EOT outcome prediction, whereas hemoglobin, despite its stable association with Favourable EOT outcome, reversed direction over time for relapse prediction.

### LLM-based representations improve relapse discrimination but reduce interpretability

To benchmark alternative representations of routine clinical data, we generated fixed-length embeddings from structured clinical inputs using the pre-trained biomedical LLM BioMistral-7B ^30^ and trained logistic regression and XGBoost classifiers on these embeddings (**Figure 1C**; **Methods**). Embeddings were generated from three input configurations: (i) common variables from the most recent visit, matching the raw model input; (ii) common variables from all visits up to each cutoff; and (iii) all available variables from all visits up to each cutoff.

For EOT outcome prediction, embedding-based models produced modest, setup-dependent improvements over raw clinical models. Models using common-variable embeddings including last or all visits performed best at later time points, achieving ROC-AUC values of 0.74-0.88 after month 2, outperforming raw last-visit models, particularly in the OFLOTUB Control arm (**Figure 4A**; **Supplementary Figure 21, 22**).

**Figure 4.**
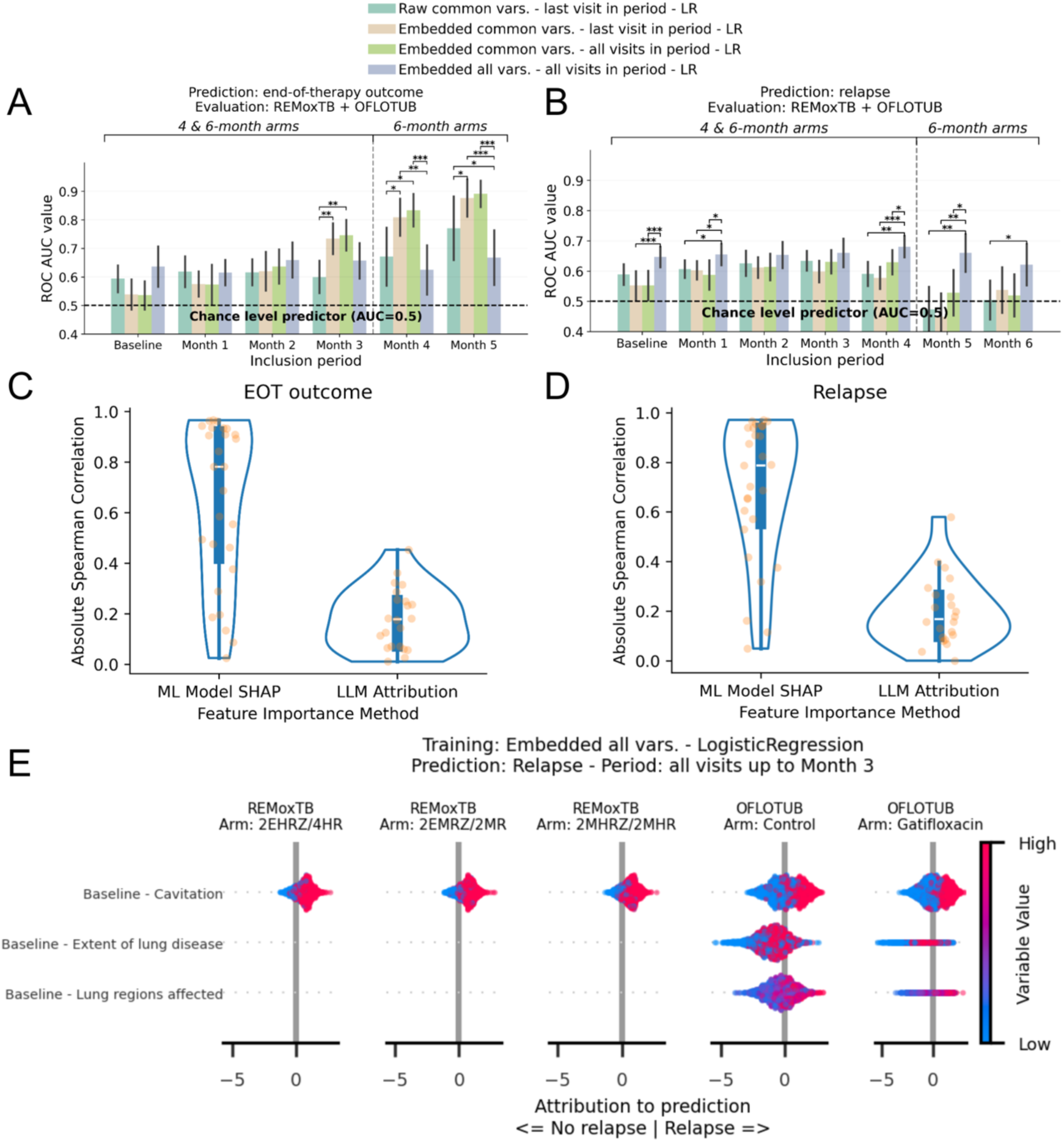
Performance-interpretability trade-off using LLM-based representations for prediction. Predictive performance of logistic regression (LR) models predicting (**A**) end-of-therapy (EOT) outcome and (**B**) relapse. Models were trained at monthly inclusion cutoffs from baseline to month 5 and month 6 for EOT outcome and relapse prediction respectively, with raw data models using common variables from the most recent visit, embedding-based models using LLM representations of the same setup, or embeddings of common variables, or all available variables from all prior visits, as indicated by the legend above. The vertical dashed line marks where 4-month arms complete treatment and drop from the analysis, leaving 6-month arms only. Bars represent mean ROC-AUC across 25 test set splits; error bars indicate standard deviation. Corrected resampled t-tests ^28^ were used to compare setups within periods, accounting for the correlation between performance estimates induced by overlapping test sets across the 25 random train-test splits, with only significant comparisons shown after Benjamini-Hochberg correction (*: adj. p<0.05, **: adj. p<0.01, ***: adj. p<0.001). The dashed line indicates chance-level performance (AUC = 0.5). (**C-D**) Distribution of absolute Spearman correlations (|ρ|) between clinical variable values and their impact measures (SHAP value for raw models; Integrated Gradient attributions for LLM-based models) for end-of-therapy outcome (**C**) and relapse prediction (**D**). Lower |ρ| in LLM-based models indicates weaker monotonic alignment between variable magnitude and model contribution. (**E**) Integrated gradients attributions for selected chest X-ray variables in a relapse prediction model trained on embeddings of all variables across all visits up to month 3. These variables showed both high attribution scores and strong positive correlations with their values, enhancing interpretability. Each point represents a participant stratified by treatment arm, with colour indicating the variable value. Positive values indicate contributions toward relapse.

In contrast, relapse prediction showed more consistent benefit from embeddings incorporating all variables across visits. These models partially mitigated the late-treatment decline in discrimination observed with raw clinical models and outperformed last-visit models from month 4 onward, particularly in participants receiving the OFLOTUB Control regimen (**Figure 4B; Supplementary Figure 23**).

To assess interpretability, we applied integrated gradients ^31^ to attribute embedding-based predictions back to original clinical variables (**Methods**). Although embedding-based models relied on many of the same clinical variables identified by SHAP, including sputum microbiology, demographic characteristics, and laboratory measures (EOT outcome: **Supplementary Figure 24-29;** relapse**: Supplementary Figure 30-36**), the alignment between variable values and their attribution scores was less monotonic than in raw clinical models, reflected in lower absolute Spearman correlations (**Figure 4C,D**; **Supplementary Tables 1-4**). Among the few variables with stronger monotonic relationships, some contributed little to overall predictions (e.g. smoking history and isoniazid resistance) or showed substantial temporal (e.g. LJ-culture, sputum-smear) or between-arm variability (e.g. age, male sex; **Supplementary Figure 24-29; Supplementary Figure 37-43**).

Chest X-ray variables, including cavitation, disease extent, and the number of affected lung regions, were a notable exception. These features showed both strong positive monotonic associations with relapse and substantial contributions to model predictions (**Figure 4E**; **Supplementary Figure 42, 43**), consistent with previous studies ^5,6,32,33^.

### Model predictions reveal limited and data-input-dependent relapse risk stratification

We next evaluated whether incorporating on-treatment data improved post-treatment relapse risk stratification among participants with favourable EOT outcomes and follow-up completion (**Methods**). We compared model-derived risk groups with the established easy-versus hard-to-treat (ETT/HTT) severity stratification proposed by Imperial et al. ^5^, based on baseline cavitation and sputum-smear grade. Participants were classified into low-and high-risk groups using averaged model predictions up to month 4. Kaplan-Meier relapse estimates were compared separately for pooled 4-and 6-month treatment arms, with follow-up right-censored at 12 months after EOT to maximise the number of participants at risk while capturing most relapse events (**Methods**).

ETT versus HTT stratification separated relapse risk in the 4-month cohort (relapse-free survival 91.4% (n = 721) vs. 82.9% (n = 426); χ² = 20.02, p = 7.67 × 10^-6^), but not in the 6-month cohort (χ² = 2.68, p = 0.10), indicating that baseline severity markers did not generalise across treatment durations (**Figure 5A**; **Supplementary Figure 44A**). Survival curves diverged early and remained separated throughout follow-up in the 4-month cohort.

**Figure 5.**
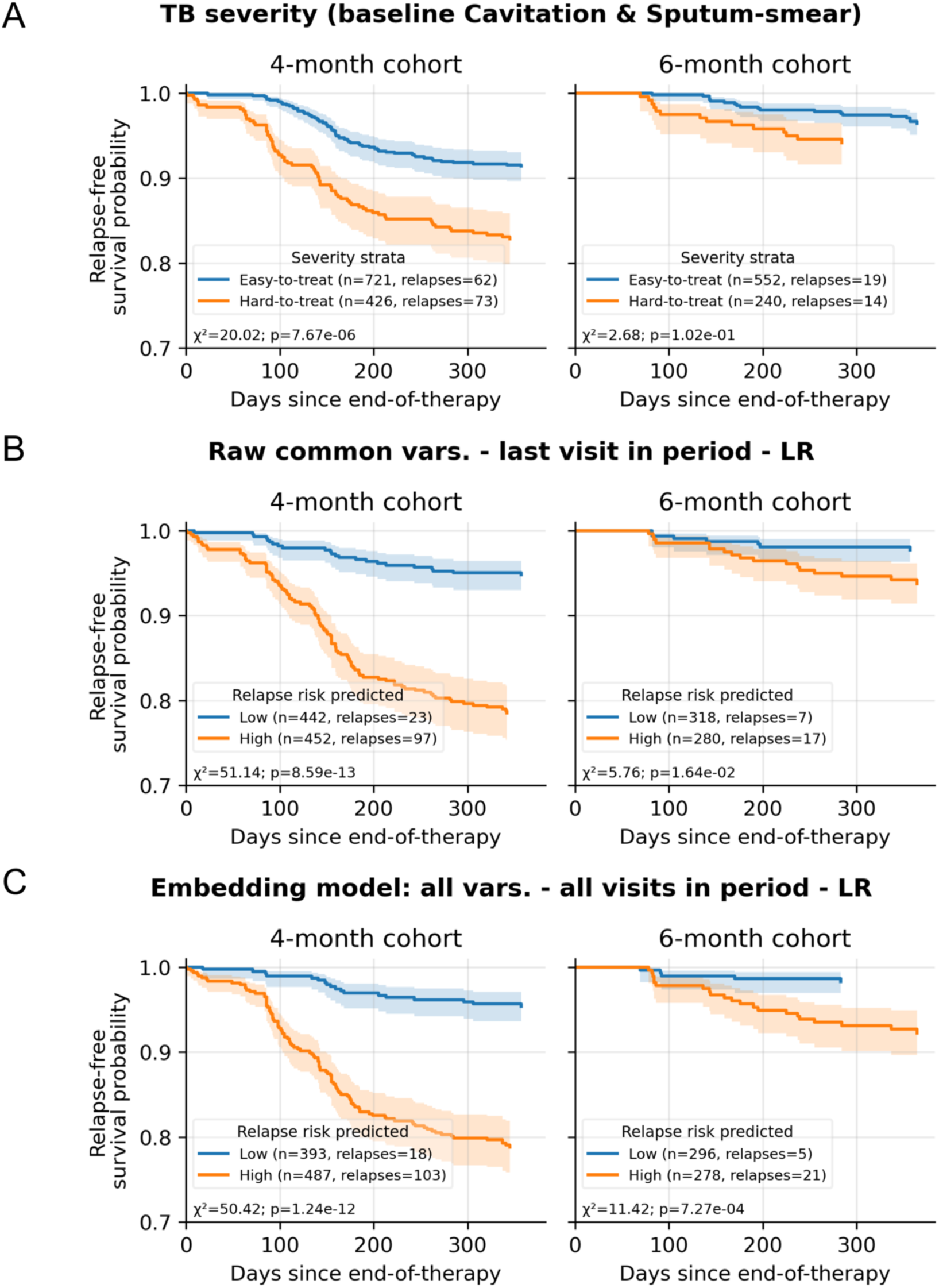
Relapse-free survival by baseline and model-based participant stratification across treatment durations. Relapse-free survival was estimated as a function of time since end-of-therapy across subgroups stratified by treatment duration (4-or 6-month). Subgroups were defined as easy-versus hard-to-treat according to Imperial et al. ^5^ (**A**), or as low-versus high-risk based on predictions from raw last-visit models (**B**), or LLM-based models incorporating all variables across all visits (C). Survival probabilities were estimated using inverse probability-of-study weighted Kaplan-Meier analysis, effect sizes and statistical significance are reported as chi-squared (χ²) values and raw p-values, respectively. The number of participants and relapse events per subgroup are indicated in the subplot legends. LR: logistic regression.

Low versus high risk groups derived from raw clinical models using on-treatment information achieved significant separation in the 4-month cohort (94.8% (n = 442) vs. 78.5% (n = 452); χ² = 51.14, p = 8.6 × 10^-13^), with early divergence maintained throughout follow-up (**Figure 5B; Supplementary Figure 44B**). Significant separation was also observed in the 6-month cohort (97.7% (n = 318) versus 93.8% (n = 280); χ² = 5.76, p = 1.64 × 10^-2^), although the absolute difference between groups was smaller.

Low versus high risk groups of embedding-based models incorporating all variables across visits also achieved significant separation in both the 4-month (95.4% (n = 393) vs. 78.9% (n = 487); χ² = 50.42, p = 1.24 × 10⁻^12^) and 6-month cohorts (98.3% (n = 296) vs. 92.3% (n = 278); χ² = 11.42, p = 7.27 × 10^-4^; **Figure 5C**). As with the raw models, separation emerged early in the 4-month cohort, whereas survival curves separated only later during follow-up in the 6-month cohort.

Imperial et al. demonstrated the non-inferiority of 4-month versus 6-month treatment at 24 months after start-of-therapy among participants within the easy-to-treat severity phenotype, using composite unfavourable outcomes (relapse, treatment failure, death, lost to follow-up) ^5^. We evaluated whether its reported non-inferiority of 4-month versus 6-month regimens could be reproduced in our cohort, restricted to participants with favourable EOT outcomes and completed follow-up, using relapse at 12 months post-EOT as the endpoint. Following the analytical framework of Imperial et al. and including Rifaquin ^34^ participants, we compared relapse risk using a 3% non-inferiority margin, instead of the original 6% (**Methods**).

Interaction analysis showed no significant interaction between ETT/HTT groups and regimen duration (4 versus 6 months; p = 0.45). However, non-inferiority of the 4-month regimen was not supported within the ETT group (risk difference 7.1 percentage points; 90% CI, 4.5-9.4; **Supplementary Figure 45**).

## Discussion

Our study shows that routinely collected clinical data contain only limited prognostic information for tuberculosis relapse. Whereas prediction of end-of-therapy (EOT) outcomes improves as treatment progresses, relapse predictability increases only modestly during the first months of therapy and does not substantially improve with continued routine clinical monitoring. The declining performance at months 5 and 6 partly reflects our data inclusion approach: at these timepoints, only 6-month arm participants were included, where lower relapse rates limited learning.

This divergence likely reflects fundamental differences in the biology captured by routine clinical data. Whereas EOT outcomes become increasingly predictable through contemporaneous microbiological measures of bacterial burden, relapse prediction relies more on stable host characteristics and gains comparatively little from repeated microbiological or laboratory measurements. These observations suggest that routine clinical variables capture only part of the biology underlying relapse, leaving much of the prognostic information inaccessible to conventional clinical monitoring. Notably, bacterial burden and laboratory markers played a smaller relative role than in EOT prediction, while invariant host characteristics (age, sex) and clinical symptoms (cough, chest pain) gained relative importance. Some of the identified relapse-associated variables warrant closer interpretation, as they may seem counterintuitive. Lower ALAT and creatinine, rather than higher, were associated with relapse, while higher ASAT predicted relapse risk. As low ALAT and creatinine levels are established markers of reduced muscle mass and frailty ^35–39^, these associations may reflect poorer physiological reserve in patients who subsequently relapsed. Because ASAT, unlike ALAT, also reflects extrahepatic tissue injury, this pattern may indicate systemic stress, consistent with its reported prognostic role in infectious diseases ^40,41^.

Models based on LLM-derived embeddings matched or exceeded raw model performance, with effects differing by prediction label: additional data reduced EOT prediction performance but improved relapse prediction, partially mitigating late-treatment performance decline. These gains however came at the cost of substantially reduced interpretability. Improvements were most pronounced in the 4-month treatment cohort, whereas embedding-based models including longitudinal and sparse data provided greater discrimination in the lower-relapse 6-month cohort.

Models incorporating data beyond baseline improved post-treatment relapse risk stratification compared to ETT/HTT stratification using cavitation and sputum-smear at baseline. Improvements were most pronounced in the 4-month treatment cohort, whereas embedding-based models including longitudinal and sparse data provided greater discrimination in the lower-relapse 6-month cohort. This discrimination could eventually inform clinical monitoring: flagging high-risk participants for closer follow-up, enabling earlier relapse diagnosis, and reducing both individual health consequences and transmission to close contacts.

Previous work demonstrated non-inferiority of 4-month versus 6-month treatment in the ETT groups using a composite unfavourable outcome. In our cohort of participants with a favourable EOT outcome and completed follow-up, using relapse as the endpoint, 4-month treatment was associated with a higher-than-acceptable relapse risk at 12 months post-EOT among ETT participants. This suggests that even easy-to-treat participants may benefit from longer therapy when relapse prevention is the priority.

Several limitations should be considered. Analyses were restricted to two Phase 3 trials, and residual protocol heterogeneity despite harmonisation may have influenced variable availability. Low relapse rates, particularly in the 6-month regimens, limited statistical power at later time points. In addition, because these trials predated the widespread adoption of Xpert MTB/RIF, microbiological monitoring relied on smear microscopy and solid culture ^42^, which may underrepresent residual low-burden disease. Together, these limitations highlight the need for more sensitive biomarkers, including lipoarabinomannan (LAM), mycobacterial load assays (MBLA) and rRNA synthesis ratio (RS ratio), which may better capture residual bacterial burden and improve relapse prediction and post-treatment relapse risk stratification^43–46^.

In conclusion, relapse predictability is inherently constrained by Mtb persistence, low relapse rates, and limited variable informativeness of routinely collected clinical data. Together, these findings suggest that the primary limitation lies in the *information content* of routine clinical data *rather than the modelling approach*. Despite these limits, incorporating routine clinical data after baseline improved post-treatment relapse risk stratification beyond ETT/HTT stratification - which uses cavitation and sputum-smear at baseline - indicating that routine clinical data retain utility even though their predictive information is limited.

## Methods

### Preprocessing

#### Definition of prediction tasks

We defined two binary prediction tasks: 1) End-of-therapy (EOT) outcome using labels as defined in TB-PACTS (Favourable/Unfavourable), and 2) Relapse among participants with a Favourable EOT outcome, (individuals who completed follow-up, excluding those who experienced death, loss to follow-up, acquired drug resistance, or reinfection during follow-up). Participants with a confirmed relapse at any follow-up timepoint were labelled Relapse; participants with favourable outcomes at all follow-up timepoints (and TB-1021 participants with missing month-12 outcomes but favourable month-18 outcomes without treatment) were labelled No relapse. Restricting both tasks to using data from the initial treatment period (4 or 6-months) avoided confounding by retreatment for relapse.

#### Study selection for analysis

We selected two phase 3 trials with available EOT outcomes and sufficient common variables across studies: TB-1021 (REMoxTB ^26^) and TB-1022 (OFLOTUB ^25^), comprising 2,918 participants. Variable selection is described in the Supplementary Methods. Both studies were designed to evaluate the potential for treatment shortening by comparing standard 6-month regimens to quinolone-containing 4-month regimens. A third trial, Rifaquin ^34^ pursued a similar aim but was excluded from the main analysis due to insufficient overlap in variables with the other two studies. Outcome measurements were available at end of therapy (4 or 6 months after therapy start for both studies, depending on regimen) and at follow-up (REMoxTB: 12 and 18 months; OFLOTUB: 18 and 24 months).

#### Preprocessing of microbiological test results

Microbiological test results were harmonised across studies. OFLOTUB sputum smear grading was aligned to the REMoxTB reporting system using the REMoxTB laboratory manual’s conversion chart, as in Imperial et al. ^5^. For visits with multiple measurements of the same method (sputum smear, LJ culture and, for REMoxTB, MGIT), we derived a single result by taking the mode, assigning positivity in case of ties. For multiple positive sputum smear tests the highest grade was used. For multiple positive MGIT measurements the average was taken. REMoxTB MGIT results identified as contaminated by blood-agar validation cultures were removed, and false-positive MGIT and LJ-culture results were corrected to negative following Murphy et al. ^47^. REMoxTB’s protocol-defined culture-negative status ^26^ was used to derive LJ-culture status and MGIT-culture status variables. For OFLOTUB, LJ-culture status was set equal to the LJ-culture result, as applying the REMoxTB algorithm was not feasible due to lower testing frequency.

#### Variable selection and data imputation

Non-sparse common variables were selected for each study configuration (REMoxTB only, OFLOTUB only, or pooled) by optimising the trade-off between variable count and participant retention. Missing numerical laboratory results were interpolated within the therapy period and forward-filled thereafter. Extreme potassium values (>12 mg/L) were replaced with the mean of all values ≤ 12 mg/L. OFLOTUB microbiology variables not measured at every visit were forward-filled. Race data, missing for all OFLOTUB participants, were set to “Black” as all study sites were in Africa ^5^.

#### Definition of prediction periods and participant selection per period

We employed a sliding-window approach for both prediction tasks, training separate models at each monthly cutoff using only data available up to that point. The Baseline period spanned 30 days before the therapy induction to day 5 of therapy. Subsequent monthly periods used therapy-day cutoffs (Month 1: day 31; Month 2: day 62; Month 3: day 93; Month 4: day 125; Month 5: day 155; Month 6: day 156 and beyond). Participants who did not complete at least 80 days of therapy were excluded from all analyses. For periods Month 4 and later, only participants treated beyond the previous period’s cutoff plus 5 days were included, to accommodate variability in visit timing.

Because EOT outcome labels were determined at Month 4 (4-month regimens) or Month 6 (6-month regimens) from microbiological results that were also predictors, including the EOT timepoint would render prediction trivial. For EOT prediction, we therefore excluded this timepoint, restricting inclusion to Baseline-Month 3 for 4-month and Baseline-Month 5 for 6-month regimens. For relapse prediction, as the outcome is determined after EOT, microbiological results at EOT do not confound prediction, all timepoints were therefore included (Baseline-Month 4 and Baseline-Month 6, respectively). Visit-window rules are in Supplementary Methods.

#### LLM-based embedding of raw clinical data

We additionally represented participant data as embeddings from the pre-trained biomedical LLM BioMistral-7B, without fine-tuning, as pre-trained LLMs can accommodate sparse variables, are well-suited for sequential modelling, and could potentially utilise knowledge accumulated during its pre-training on millions of biomedical articles ^30,48–51^. For each participant, tabular data were serialised into a single string consisting of an instruction prompt followed by modality-prefixed substrings (e.g., “Microbiological test results of participant:”), each appended with therapy day, value, and unit. The string was tokenised and passed through BioMistral-7B; the token-axis mean of the final hidden state yielded a 4096-dimensional embedding per participant, used as input for downstream models ^52^. The instruction prompt and implementation details are in Supplementary Methods.

#### Experimental setups of input data

Four input setups were defined for each prediction period: (i) **common variables - last visit in period**: non-sparse imputed variables from the most recent visit before the cutoff (raw and LLM approaches), enabling direct comparison of the effect of LLM embedding; (ii) **common variables - last visits concatenated**: as (i), but with continuous variables included separately for each prior visit (e.g., heart rate at Month 1, Month 2, etc.), allowing non-sequential models to capture temporal trends (raw); (iii) **common variables - all visits in period**: non-sparse imputed variables from all visits up to the cutoff, testing if full visit history improved prediction in sequential models (LSTM and LLM); (iv) **all variables - all visits in period**: all non-sparse and sparse variables from all visits up to the cutoff, without imputation (LLM only).

For the Baseline period, the data window spanned 30 days before therapy induction to day 5; missing values were forward-and backward-filled, with the backward window extended to 30 days for participants still missing non-sparse variables (mostly OFLOTUB). Values closest to therapy day 1 were then used as Baseline. Two Baseline setups were applied: one restricted to non-sparse variables (raw approach plus the corresponding LLM setups), and one including all clinical variables (LLM “all variables - all visits in period” setup only).

### Model training and evaluation

#### General overview of model training pipeline

For each combination of input type, prediction period, prediction task, and experimental setup, we trained logistic regression (L1-regularised) and XGBoost as established clinical prediction methods, alongside bidirectional long short-term memory (LSTM) models as a sequential baseline ^53–57^. As LSTMs require dense inputs, the latter were only trained on the “common variables - all visits in period” setup. To account for variability in performance estimates and class imbalance, we generated 25 external training-testing splits stratified by outcome label and study. Hyperparameter search was performed on the training splits, and final models were evaluated on the corresponding test sets. Details on LSTM structure, hyperparameter search, model training and testing are in Supplementary Methods.

### Model interpretation

#### SHAP-value calculation

We computed SHAP values ^29^ for the raw-data logistic regression and XGBoost models on test-set predictions, LSTM models were excluded due to poor performance and computational cost. For logistic regression we used a linear explainer with imputation-based masking on training data, for XGBoost we used a tree explainer with interventional feature perturbation, training data as background. Both choices account for correlations among input variables.

#### Attribution of variables to model output using integrated gradients method

To attribute LLM-embedding model predictions back to clinical variables, we applied integrated gradients (IG) ^31^ to the logistic regression models trained on embeddings. XGBoost was excluded as IG required a differentiable pipeline. The baseline input consisted of the instruction prompt and modality introductions only, with variable names, values, and units removed. Attributions were computed on the token-embedding layer of BioMistral-7B with 24 interpolation steps. For each token, attributions were averaged across the 4096 embedding dimensions. Per-variable attributions were obtained by summing across the variable’s name and value tokens. Positive values indicated contributions toward the positive label (Unfavourable or Relapse). Attributions were standardised within each of the 25 CV splits for cross-split comparability, implementation details are in **Supplementary Methods**.

#### Correlation between input variables and SHAP values and integrated gradients attributions

For each prediction period, we computed correlations between raw input values and their impact metric (SHAP for raw-data models, IG attributions for embedding-based models) within each test set of the 25 splits, stratified by treatment arm: Spearman for continuous variables, point-biserial for binary variables. P-values were corrected within each period using the Benjamini-Hochberg procedure (FDR). For each variable we recorded the number of splits with FDR < 0.05 and aggregated statistics (mean FDR, mean correlation, mean absolute impact metric) across splits; full results are in **Supplementary Tables 1, 2** (SHAP) and **Supplementary Tables 3, 4** (IG). IG attributions for selected variables in the “all variables - all visits” relapse setup are shown in **Supplementary Figures 24-43**, variable selection criteria are described in the **Supplementary Methods**.

### Subgroup analysis in relapse prediction cohort

#### Relapse-free survival estimation within participant subgroups

To assess relapse-risk stratification, we assigned participants to low-and high-risk groups using prediction probabilities from the last-visit raw-data models and the all-variables/all-visits embedding-based models. Because participants could appear in multiple of the 25 splits, we first computed a single per-period prediction probability per participant by averaging across all splits in which they appeared. Hierarchical clustering of these per-participant trajectories revealed temporally consistent low-and high-prediction subclusters (**Supplementary Figure 46**), supporting aggregation across periods. We therefore used the average prediction probability across baseline to month 4 (chosen to maximise participant inclusion) as the final risk score.

Arm-specific thresholds were applied to account for arm-level differences in relapse rates: low-risk ≤ 30th and high-risk ≥ 70th quantile of training-set averages, applied to test-set participants for both models. As an established comparator, we also applied the easy-and hard-to-treat (ETT/HTT) classification of Imperial et al. ^5^, based on baseline cavitation and sputum smear grade. Subgroups were analysed within regimen-duration cohorts (4-or 6-month), following Imperial et al. For 4-month regimens, end-of-therapy was defined as the last study day of non-placebo drug administration. Survival was right-censored at 12 months post-EOT, capturing the majority of relapses while maximising the number of participants, given heterogeneous follow-up durations (REMoxTB: 18 months from start of therapy; OFLOTUB: 24 months from EOT). Relapse-free survival was estimated using inverse probability-of-study weighted Kaplan-Meier analysis ^58^.

#### Non-inferiority analysis of easy-to-treat participants using relapse after favourable EOT outcome

We assessed whether the non-inferiority of 4-month versus 6-month treatment in easy-to-treat participants ^5^ also holds in our cohort restricted to participants with favourable EOT outcomes and completed follow-up, using relapse alone rather than a composite outcome. We pre-specified a non-inferiority margin appropriate for relapse as standalone outcome, as the 6% margin used by REMoxTB and OFLOTUB ^25,26^ was designed for composite unfavourable outcomes (relapse, treatment failure, death, lost to follow-up), with control-arm event rates of approximately 13-20% ^59^. Relapse rates under standard 6-month therapy are however typically 3-5% with an absolute increase of 9-10% on shortening to 4 months ^60–62^. Following regulatory guidance to preserve ≥50% of the active-control effect ^63^, we chose a more conservative 3% margin. To match Imperial et al.’s pooled analysis, we additionally included Rifaquin participants ^34^, classifying those with a favourable per-protocol outcome and no subsequent relapse as no relapse after cure (n=470), and those with a recorded relapse during follow-up as relapse after cure (n=33).

Following Imperial et al.: interaction was tested using Cox regression with subgroup (easy-or hard-to-treat), regimen duration, and their interaction term ^64^. Non-inferiority was assessed by estimating the relapse risk difference at 24 months after start of therapy, between subgroups within regimen-duration cohorts, using inverse probability-of-study weighted Kaplan-Meier estimates. Risk differences were computed across 500 stratified bootstrap samples (stratified by study and regimen duration); non-inferiority was determined by comparing the upper bound of the two-sided 90% confidence interval to the pre-specified 3% margin.

## Data Availability

All data produced in the present study are available upon reasonable request to the authors

## Declarations

## Competing interests

G.M.-E. is a full-time employee of GSK and holds GSK stock/share options. N.I.P. has received funding to his institution from Janssen for sequencing work on the TRUNCATE-TB trial and honoraria from Janssen for speaking at symposia and advisory board participation; serves as unpaid chair of the Data Safety Monitoring Boards for the TB Alliance NC-009 and NC-010 trials; and is Chief Investigator on the EU-funded PARADIGM4TB trial (payment to institution), which tests anti-tuberculosis drug combinations including drugs from GSK, TB Alliance, Otsuka, Janssen, and LMU. D.J.S. and M.P.M. declare no competing interests related to this manuscript.

## Use of Artificial Intelligence

In the preparation of this manuscript, we utilised the large language model GPT-4 (https://chat.openai.com/) for editing assistance, including language polishing and clarification of text. While this tool assisted in refining the manuscript’s language, it was not used to generate contributions to the original research, data analysis, or interpretation of results. All final content decisions and responsibilities rest with the authors.

## Acknowledgements

Daniel Garger and Jamison H. Burks received funding from the Innovative Medicines Initiative 2 Joint Undertaking (JU) under grant agreement No. 101007873 (UNITE4TB). Francesco Paolo Casale was funded by the Free State of Bavaria’s High-Tech Agenda through the Institute of AI for Health (AIH).

## Authors’ contributions

D.G. conducted all data preprocessing and analyses, with contributions from J.H.B. D.G. and J.H.B. jointly led data interpretation and manuscript development. N.I.P., D.J.S., and G.M.-E. contributed to study design and interpretation of results, with N.I.P. providing key input on study positioning and overall framing. D.G., J.H.B., and F.P.C. wrote the manuscript, with input from all authors. M.P.M. and F.P.C. conceptualised and supervised the study, with support from F.T.

## Code availability

The TB-PACTS dataset is publicly available upon application at https://c-path.org/tools-platforms/tb-pacts/. Code for the preprocessing and analysis performed in this study is available at https://github.com/gargerd/outcome_prediction.

