## Supplementary methods for "Time-resolved predictability of end-of-therapy outcome and relapse after cure in Phase 3 tuberculosis trials"

### Data harmonisation

TB-PACTS data were obtained from the C-Path data repository [1], as of August 2021. After reviewing the different data modalities within TB-PACTS, we selected the following domains for our analysis: adverse events, clinical events, concomitant medications, drug accountability, demographics, disposition, exposure, laboratory and microbiology test results, medical history, microbiology susceptibility, respiratory findings, subject visit, substance use, vital signs, outcomes. Prior to analysis, we harmonised these domains across all 25 studies included in TB-PACTS. Detailed descriptions of the studies and their corresponding pseudonyms are provided in **Supplementary Table 5**.

#### *Laboratory test results*

Within each study, continuous laboratory variables were already converted to consistent measurement units by the TB-PACTS curators. To ensure harmonisation across studies, we further standardised all continuous laboratory variables to common variables. Some laboratory variables were reported partly as continuous and partly as categorical. For these variables, if more than 50% of observations were categorical, we converted continuous values to categorical values based on the corresponding categorical levels. Conversely, if continuous values outweighed the categorical values, we converted the categorical values to continuous values based on the categorical levels. Where possible, categorical variables were assigned ordinal levels. Detailed descriptions of the harmonisation procedures for all laboratory variables are provided in the Jupyter Notebook used for laboratory test preprocessing (*5_laboratory_results.ipynb*).

Missing measurement days were imputed using either the measurement dates of other laboratory variables from the same or subject visit data, when available. If only the week or month of measurement was recorded, the exact day was derived from the reported week or month information. For measurements recorded at “screening” or during the “pretreatment epoch” with missing exact dates, the measurement day was assigned as day 0.

#### *Medical terms in adverse and clinical events, medical history and concomitant medication*

Because data were collected across multiple study sites, medical terminology describing diagnoses in adverse events, clinical events, medical history and concomitant medication showed substantial variability. Many terms referred to the same condition, often further complicated by typographical errors and the use of abbreviations. In total, more than 12,000 unique medical terms were identified across all studies. Due to time constraints, we therefore prioritised the standardisation of the most frequently occurring diagnosis terms within these four domains, with the goal of maximising coverage across terms and participants.

For medical history records, conditions were assumed to be absent unless their presence was explicitly recorded as “Yes.” For adverse and clinical events, toxicity grades as well as event start and end days were extracted.

#### *Respiratory findings*

Because most participants had binary labels (“Yes” or “No”) indicating the presence of cavitation, categorical variables describing cavitation extent in greater detail were converted to binary indicators where applicable to ensure consistency across studies. For additional descriptors of pulmonary tuberculosis, categorical variables were assigned ordinal levels, incorporating laterality information when available. When the specific side of the affected lung was not recorded, findings were classified as unilateral or bilateral. Missing measurement days were imputed using the same approach applied to laboratory test results.

#### *Microbiology susceptibility*

Inconclusive test results were excluded, including measurements from the same day with discordant susceptibility outcomes. Where possible, we derived a *susceptible concentration* and a *resistant concentration* for each tested strain. If a strain was susceptible at all tested drug concentrations, the susceptible concentration was defined as the lowest tested concentration (or the only concentration if a single test was performed). Conversely, if a strain was resistant at all tested concentrations, the resistant concentration was defined as the highest tested concentration (or the only concentration if a single test was performed). In cases where resistance was observed at lower concentrations and susceptibility at higher concentrations, the susceptible concentration was defined as the lowest concentration at which susceptibility was observed, and the resistant concentration as the highest concentration at which resistance was observed.

### Reconstruction of concomitant medication

TB-PACTS contains information on concomitant non-study medications administered during the study period, along with diagnosis terms describing the indication for each medication. These diagnosis terms were preprocessed separately together with adverse events, clinical events, and medical history data.

During extraction of concomitant medication names, we encountered challenges similar to those observed for medical terminology, including multiple names for the same active ingredient, abbreviations, and typographical errors. Owing to time constraints, we therefore limited the analysis to the most frequently administered concomitant medications.

Medication names were standardised based on their active ingredients, in part using the *drugstandards* Python package (Bernauer, 2017). We additionally extracted the duration of drug administration, imputing the final day of application using therapy completion data when available. Routes of administration and dosing frequencies were also extracted and standardised.

Using this information, we reconstructed concomitant medication administration at a daily resolution. Because exact administration dates were not recorded, doses were distributed across the treatment period according to the recorded dosing frequencies. This resulted in a table in which rows correspond to application days, columns represent drugs (stratified by route of administration), and values indicate daily doses. From these daily doses, cumulative drug exposure over the administration period was subsequently calculated.

### Preprocessing of final dataset for analysis

#### *Selection of phase 3 studies in TB-PACTS*

The preprocessing steps described above were applied to all available studies in TB-PACTS, including phase 1, 2, and 3 trials. However, because the objective of our analysis was to predict therapy outcomes using data pooled across multiple studies, we subsequently restricted our focus to phase 3 trials with available therapy outcome labels. Accordingly, only the following phase 3 studies were included in the final analysis: TB-1018, TB-1020, TB-1021, TB-1022, and TB-1030.

#### *Outcome label extraction*

For study TB-1018, outcome labels were extracted at the end-of-therapy and 24-month follow-up timepoints. Information on the analysis approach used for outcome definition (per-protocol or intention-to-treat) was not available.

For study TB-1020, only outcome labels from the per-protocol analysis at the 18-month follow-up timepoint were available and were therefore extracted. Information on the medium used for outcome determination was not reported.

For study TB-1021, outcome labels from the per-protocol analysis were extracted at the end-of-therapy and 12-month follow-up timepoints. For the 18-month follow-up, outcome labels determined using liquid culture were selected. For participants with no available liquid culture result at month 18, solid medium results were taken instead.

For study TB-1022, outcome labels were extracted at the end-of-therapy, 18-month, and 24-month follow-up timepoints. The analysis approach used for outcome definition (per-protocol or intention-to-treat) was not reported.

For study TB-1030, only outcome labels derived from the intention-to-treat analysis were available. Outcome labels were extracted at the end-of-therapy, 12-month, and 30-month follow-up timepoints. Information on the medium used for outcome determination was not reported.

#### *Detecting common variables across studies*

Different phase 3 studies measured partially overlapping sets of variables, resulting in a small subset of variables with broad coverage across studies and a larger subset with substantial data sparsity. To systematically identify variable sets suitable for downstream analysis, we employed a data-driven variable selection approach.

First, we constructed a data-availability matrix with participants as rows and variables as columns. Each entry was set to *True* if data for a given variable were recorded for a participant at least once during the therapy period, and *False* otherwise. Participants were then clustered into five groups based on similarity in their data-availability profiles. All possible combinations of these clusters were subsequently considered.

For each cluster combination, we evaluated only participants belonging to the selected clusters and calculated, for each variable, the number of participants with available data. Variables were ranked according to data availability, and a sequential selection procedure was applied: starting with the variable with the highest availability, we iteratively retained only participants with data for the current variable and all previously selected variables.

Finally, for each cluster combination, the trade-off between the number of retained participants and the number of commonly available variables was visualised using bar plots. This procedure was repeated separately for outcome labels available across multiple studies (end-of-therapy, 12-, 18-, and 24-month outcomes), yielding multiple candidate subsets of participants and variables for subsequent analyses.

### Additional details on model training, testing and interpretation

#### *Handling variability in visit timing*

To account for variability in visit timing between Month 2 and Month 5, if a participant's last visit occurred more than 20 days before the cutoff but another visit took place within 10 days after the cutoff, the later visit was used. This rule was also applied at Month 4 for participants in the 4-month regimen arm.

#### *LLM-based embedding extraction*

For each participant, tabular raw data were first converted into a single string. Each string began with an instruction prompt:

*[INST] The following data originates from a patient with pulmonary tuberculosis, participating in a Phase 3 clinical trial. Please summarise the condition of the patient. [/INST]*

This prompt leveraged the instruction-based training of BioMistral-7B. Data from different modalities were then appended iteratively, each prefixed with an introductory substring (e.g., "Microbiological test results of participant:"), followed by the therapy day, variable values, and measurement units. The final participant string consisted of the instruction prompt followed by all data modality substrings included in the given experimental setup.

Using the transformers package, the base BioMistral-7B model (BioMistral/BioMistral-7B) was loaded in 16-bit float precision. Each participant string was tokenised using the corresponding BioMistral-7B tokeniser, then fed into the model. The last hidden state output was extracted, and the row-wise mean of this output was computed to obtain a 4096-dimensional embedding vector for the participant. This embedding procedure was repeated for all prediction periods and all experimental setups, yielding individual participant embedding vectors for every combination.

#### *Long short-term memory (LSTM) model structure*

We implemented a bidirectional LSTM model with attention-based pooling to capture sequential patterns in longitudinal participant data. The model processes each participant's time series using an LSTM network, generating a hidden representation for each time step. An attention mechanism assigns weights to these hidden states, producing a context vector as a weighted sum of all time points. This context vector is then passed through a series of fully connected layers, with a softmax activation at the output to produce class probabilities for the two binary prediction tasks: EOT outcome and relapse prediction.

#### *Hyperparameter selection*

Hyperparameter search spaces were: logistic regression, regularisation strength $C \in \{ 0.01, 0.1, 1, 10\}$; XGBoost, for EOT out outcome prediction: *n_estimators* ∈ {200, 500}, *max_depth* ∈ {3, 5, 7}, *eta* ∈ {0.05, 0.1}, *subsample* ∈ {0.7, 0.9}, *colsample_bytree* ∈ {0.7, 0.9}, for relapse prediction: *n_estimators* ∈ {100, 200}, *max_depth* ∈ {2, 3}, *min_child_weight* ∈ {20, 40}, *subsample* ∈ {0.8}, *colsample_bytree* ∈ {0.05, 0.1, 0.2}, *learning_rate* ∈ {0.05, 0.1}; LSTM, *num_epochs* ∈ {100, 200}, *num_lstm_layers* ∈ {1, 2, 4}, *lstm_hidden_size* ∈ {64, 128}.

Hyperparameter tuning was performed separately within each of the 25 external splits. Training data were further divided into five internal training-validation folds, stratified by outcome label and study. Continuous variables were standardised, binary variables encoded as 0/1, and categorical variables one-hot encoded. Outcome labels were weighted inversely to their normalised frequency in the internal training set. For XGBoost and LSTM, hyperparameters were selected by grid search using mean ROC-AUC across the five validation folds. ROC-AUC was used throughout for its robustness to class imbalance [2].

#### *Training and testing of final models*

Final models were trained on each of the 25 eternal splits using the selected hyperparameters, with class weights set inversely to label frequency of the training set. The standard scaler was fitted on training data only and applied to the corresponding test set to prevent leakage. Test-set predicted probabilities were used to compute ROC-AUC.

#### *Attribution of variables to model output using integrated gradients method*

To address limitations in LLM-embedding interpretation [3], we applied the Integrated Gradients (IG) method [4], which interpolates feature attributions between a baseline input and the actual input, computing gradients of the model output with respect to the inputs along this interpolation path. Because (IG) requires a fully differentiable pipeline, it was applied only to logistic regression models trained on embeddings, not to XGBoost models.

The baseline input was constructed by removing variable names, values, and units from the input sentences, leaving only the instruction and data modality introduction substrings. Attribution calculations were performed using Captum's *LayerIntegratedGradients*() on the token embedding layer of BioMistral-7B, with 24 interpolation steps.

To create a fully differentiable pipeline, for each of the 25 CV-split test sets we followed the same steps as during model training: the last hidden layer for each input sentence was averaged along the token axis to produce the embedding, standardised using a scaler fitted on the training data, and fed to the trained logistic regression model. This produced an attribution matrix of shape *(number of tokens × 4096)* for each input sentence. Positive values indicated contributions toward the positive label (Unfavourable or Relapse), while negative values indicated contributions toward the negative label.

To obtain a single attribution per token, we averaged across the 4096 embedding dimensions. Variable-level attributions were obtained by summing token attributions corresponding to each variable's name and value. Since attributions were derived separately from 25 CV-split models, we standardised the attribution values within each split to ensure comparability, and reported the standardised values.

#### *Variable prioritisation for LLM-embedding interpretation*

For the "all variables - all visits" setup, which yielded a large number of variables, we applied three complementary selection strategies based on the aggregated statistics in **Supplementary Tables 3** and **4**, retaining the top 40 unique variables per strategy:

1. **Highest attribution scores**: Variables with mean >15 participants per arm across splits, ranked by mean absolute attribution.
2. **Consistent correlations between input and attributions**: Variables with statistically significant correlations (FDR < 0.05) in >20 of 25 splits and mean >15 participants per arm, ranked by mean absolute attribution.
3. **Sparse variable capture**: Variables with limited availability (mean 2-15 participants per arm) not well-represented at other timepoints, ranked by mean absolute attribution.
