## Supplementary figures for "Time-resolved predictability of end-of-therapy outcome and relapse after cure in Phase 3 tuberculosis trials"

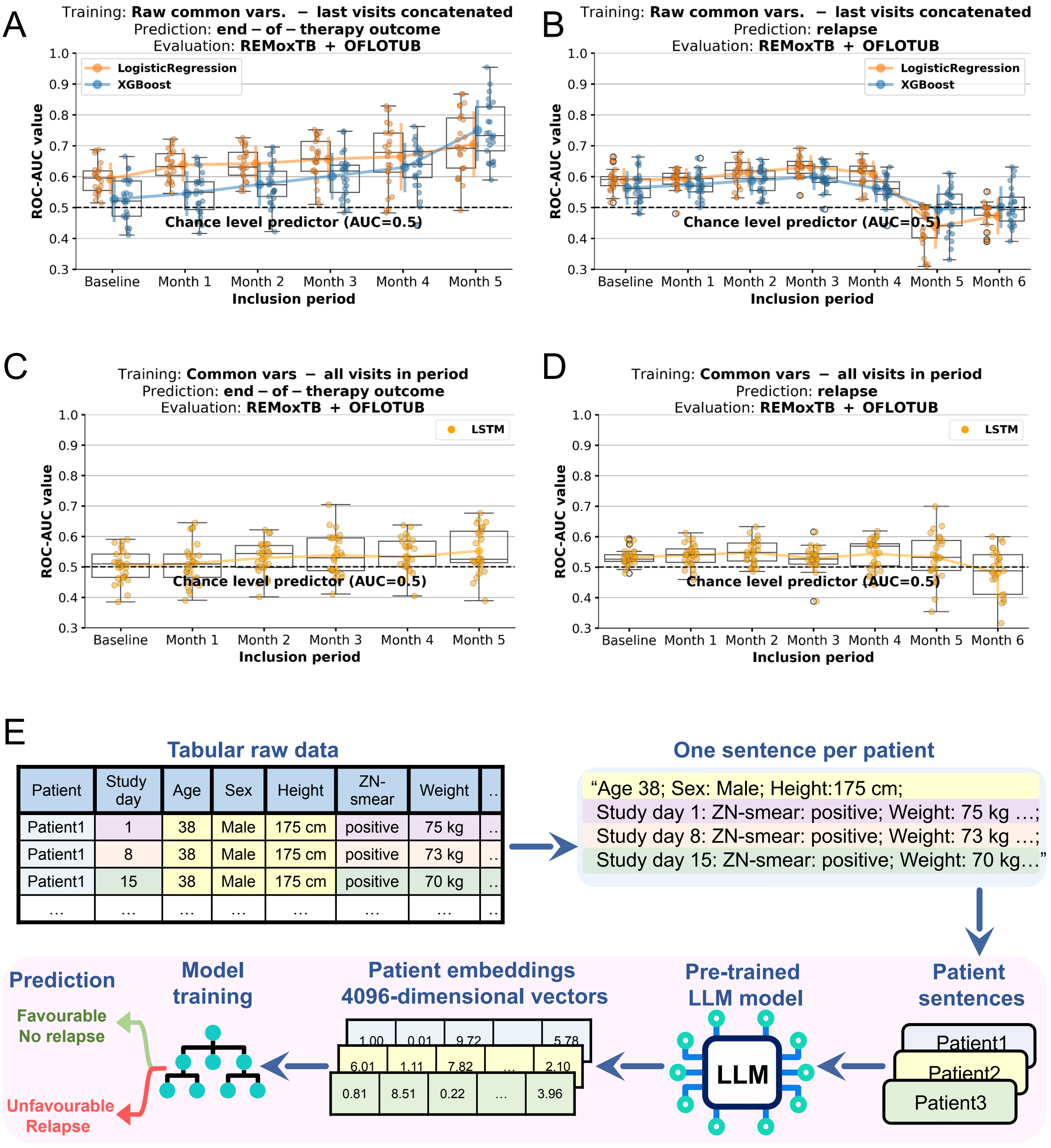


**Supplementary Figure 1: Prediction performance of models trained with sequential data approaches and overview of embedding steps. (A)** End-of-therapy (EOT) outcome prediction and (**B**) relapse prediction using logistic regression (LR) and XGBoost models trained on raw tabular clinical and treatment data. Models were trained at monthly inclusion cutoffs from baseline to month 5 and month 6 for EOT outcome and relapse prediction respectively, using data formed by concatenating all prior "last visit in period" values up to each cutoff ("last visits concatenated"), including common clinical and treatment variables across studies. **(C)** End-of-therapy (EOT) outcome prediction and (**D**) relapse prediction using long short-term memory (LSTM) model, trained on raw clinical and treatment data. Models were trained at monthly inclusion cutoffs from baseline to month 5 and month 6 for EOT outcome and relapse prediction respectively, using sequential data from all prior visits to the cutoff. Evaluation for models in (A-D) was performed across the combined REMoxTB and OFLOTUB cohorts. Bars indicate mean predictive performance (ROC-AUC), with error bars showing the standard deviation across 25 test sets from repeated 25×5 cross-validation. The dashed horizontal line denotes chance-level performance (ROC-AUC = 0.5). (**E**) Overview of embedding steps. Raw tabular data were converted into sentences and fed into BioMistral-7B. Mean pooling of the last hidden layer produced a 4096-dimensional patient embedding, which was then used for training and prediction.

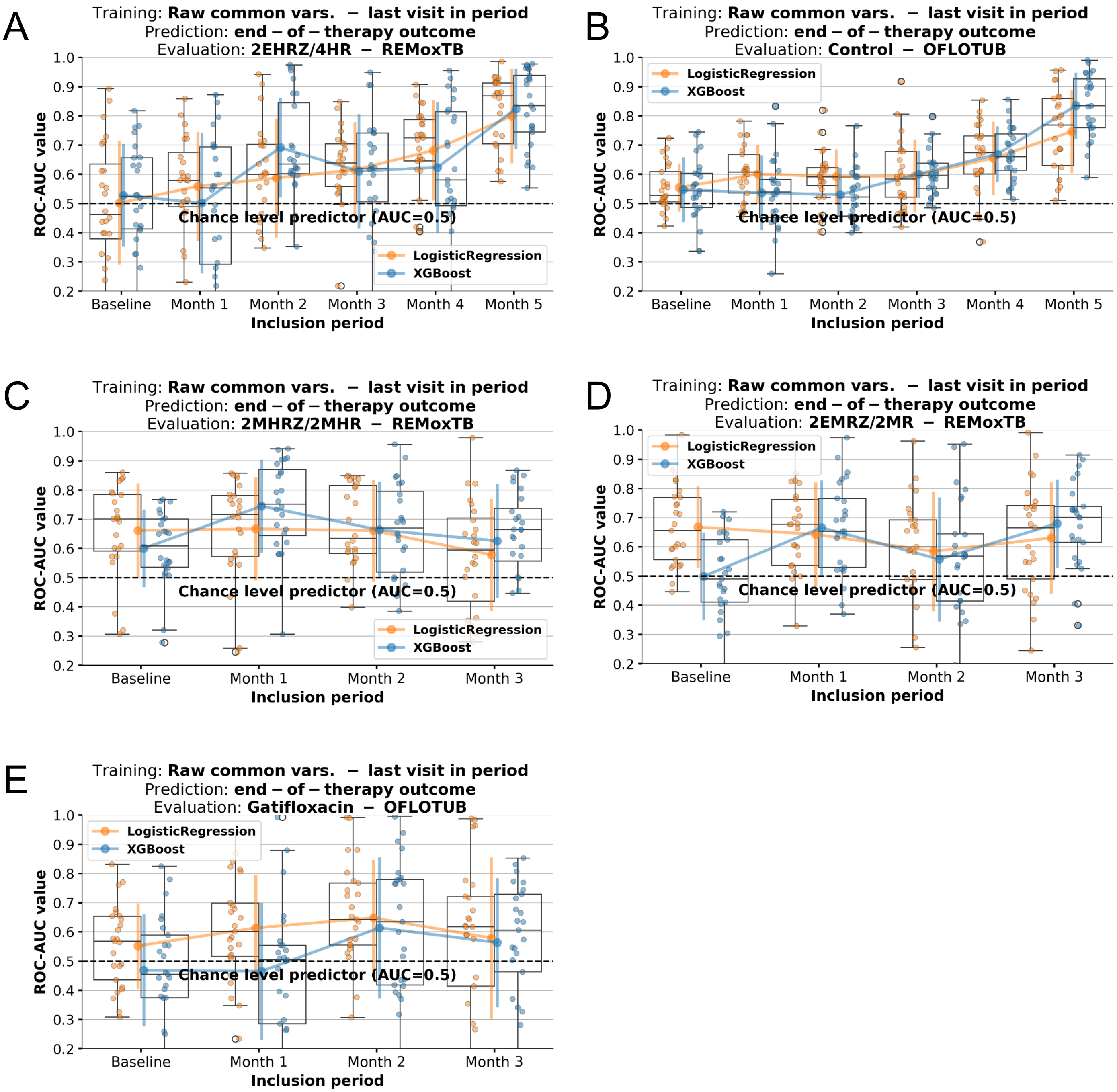


**Supplementary Figure 2**: **Within arm prediction performance of end-of-therapy outcome models trained with last-visit raw data including common variables.** End-of-therapy (EOT) outcome prediction using logistic regression (LR) and XGBoost models trained on raw tabular clinical and treatment data. Models were trained at monthly inclusion cutoffs from baseline to month 3 and month 5 for EOT outcome prediction, using common clinical and treatment variables across studies from the most recent visit prior to each cutoff ("last visit in period"). (**A-E**) panels show the within-arm performance of models evaluated on patients from the corresponding treatment arms included in the analysis. Corresponding arm and study names are indicated above the subplots. Bars indicate mean predictive performance (ROC-AUC), with error bars showing the standard deviation across 25 test sets from repeated 25×5 cross-validation. The dashed horizontal line denotes chance-level performance (ROC-AUC = 0.5).

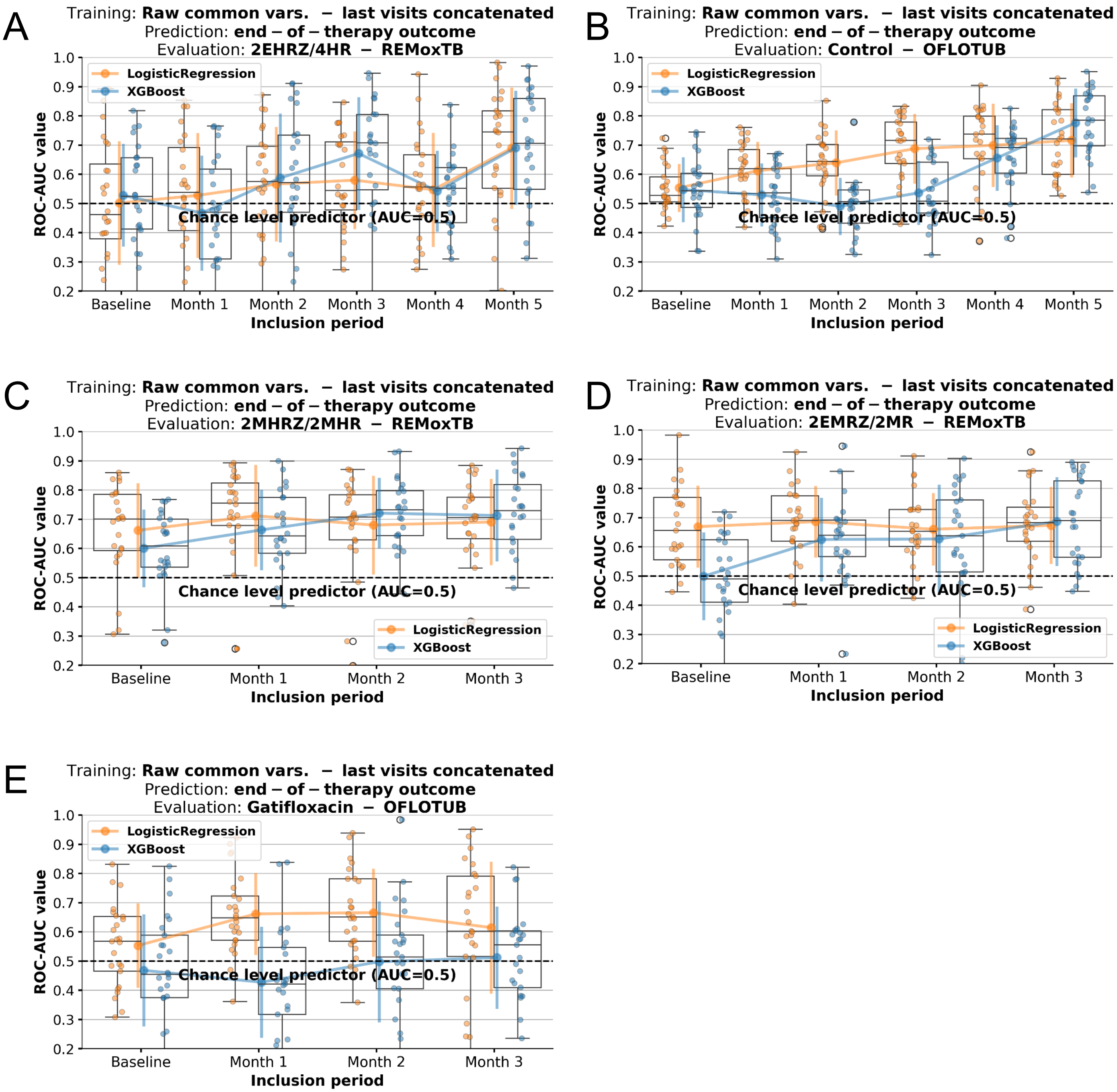


**Supplementary Figure 3**: **Within arm prediction performance of end-of-therapy outcome models trained with last-visits concatenated raw data.** End-of-therapy (EOT) outcome prediction using logistic regression (LR) and XGBoost models trained on raw tabular clinical and treatment data. Models were trained at monthly inclusion cutoffs from baseline to month 3 and month 5 for EOT outcome prediction, using data formed by concatenating all prior "last visit in period" values up to each cutoff ("last visits concatenated"), including common clinical and treatment variables across studies. (**A-E**) panels show the within-arm performance of models evaluated on patients from the corresponding treatment arms included in the analysis. Corresponding arm and study names are indicated above the subplots. Bars indicate mean predictive performance (ROC-AUC), with error bars showing the standard deviation across 25 test sets from repeated 25×5 cross-validation. The dashed horizontal line denotes chance-level performance (ROC-AUC = 0.5).

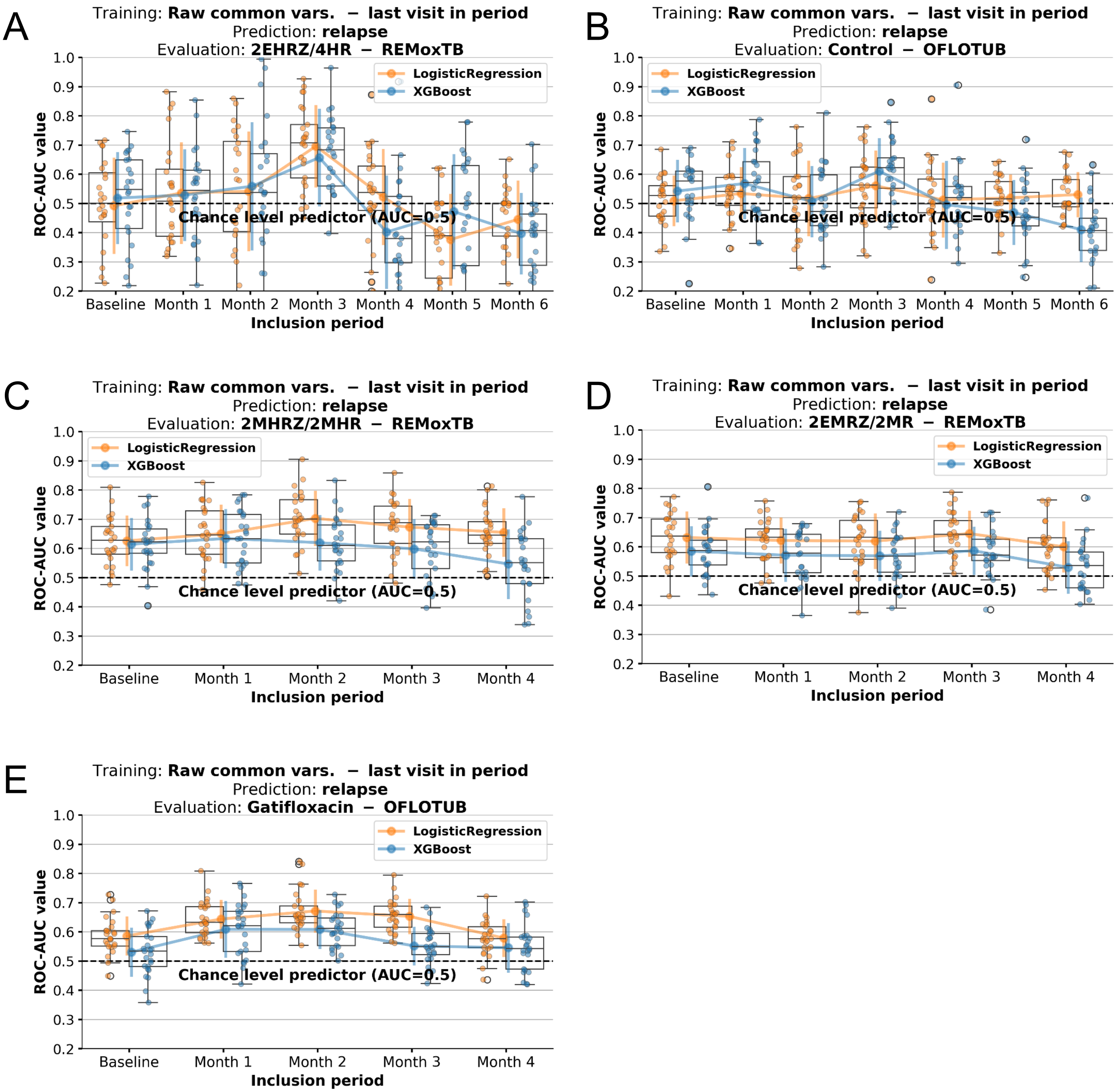


**Supplementary Figure 4**: **Within arm prediction performance of relapse prediction models trained with last-visit raw data.** Relapse outcome prediction using logistic regression (LR) and XGBoost models trained on raw tabular clinical and treatment data. Models were trained at monthly inclusion cutoffs from baseline to month 5 and month 6 for relapse prediction, using common clinical and treatment variables across studies from the most recent visit prior to each cutoff ("last visit in period"). (**A-E**) panels show the within-arm performance of models evaluated on patients from the corresponding treatment arms included in the analysis. Corresponding arm and study names are indicated above the subplots. Bars indicate mean predictive performance (ROC-AUC), with error bars showing the standard deviation across 25 test sets from repeated 25×5 cross-validation. The dashed horizontal line denotes chance-level performance (ROC-AUC = 0.5).

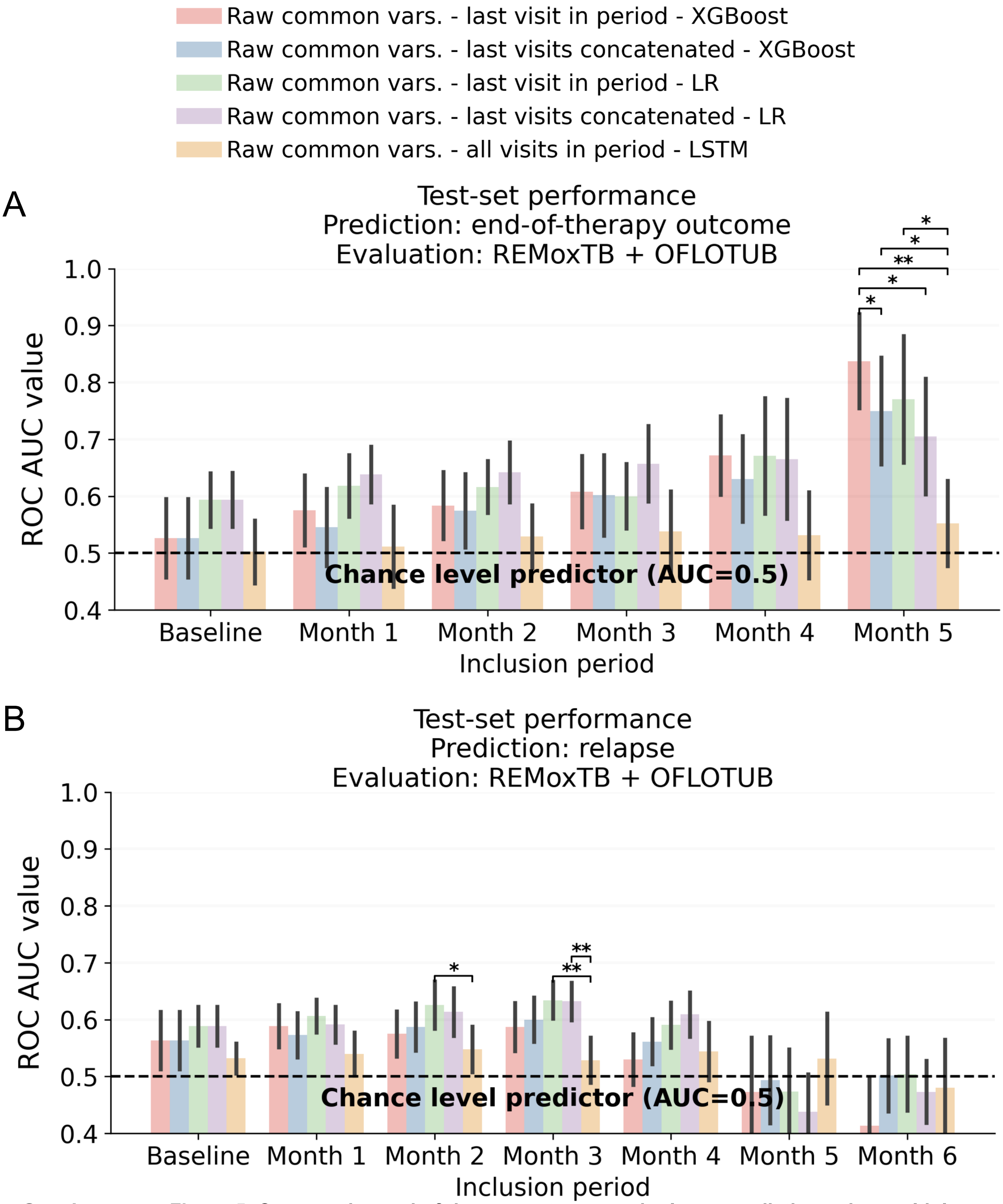


Supplementary Figure 5: **Comparative end-of-therapy outcome and relapse prediction using multiple raw data approches.** Predictive performance (ROC-AUC) of XGBoost, logistic regression (LR) and long short-term memory (LSTM) models trained on two input setups: raw tabular data of common clinical and treatment variables across studies, taken from the last visit in each period ("Raw common vars. – last visit in period"); or using data formed by concatenating all prior "last visit in period" values up to each cutoff ("last visits concatenated"). Subplot (**A**) shows results of end-of-therapy outcome, (**B**) results of relapse prediction. Evaluation for all models included in the plot was performed across the combined REMoxTB and OFLOTUB cohorts. Legend above shows colour coding of the input setup-model combinations used for prediction. Bars represent mean ROC-AUC across 25 test set splits; error bars indicate standard deviation. Corrected resampled t-tests (Nadeau & Bengio, 2003) were used to compare setups within periods, accounting for the correlation between performance estimates induced by overlapping test sets across the 25 random train-test splits, with only significant comparisons shown after Benjamini-Hochberg correction (*: adj. p<0.05, **: adj. p<0.01, ***: adj. p<0.001). The dashed line indicates chance-level performance (AUC = 0.5)

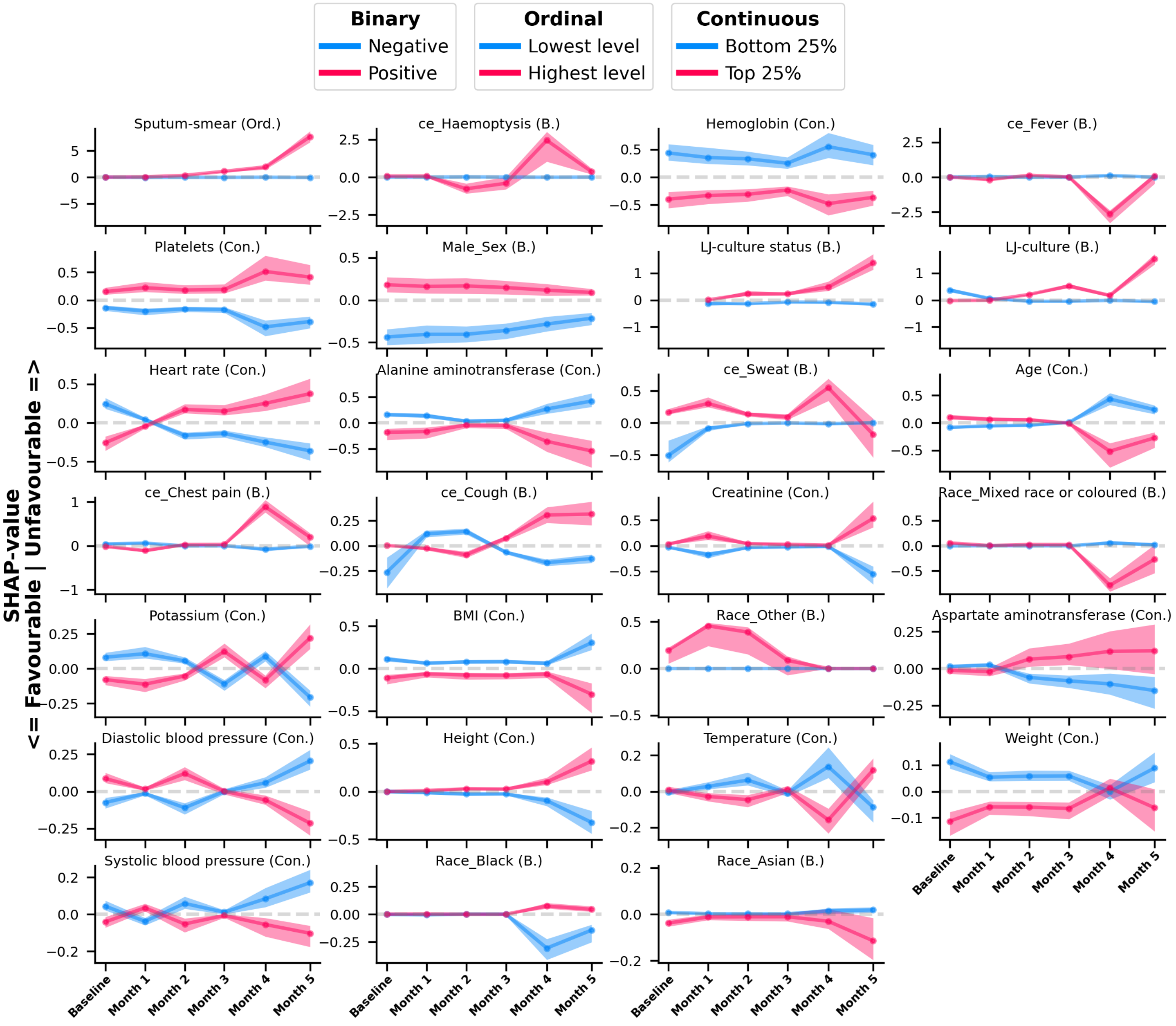


**Supplementary Figure 6**: **Population-level SHAP value trajectories across therapy periods for all clinical variables used for end-of-therapy outcome prediction**. Population-level SHAP value trajectories from last-visit raw-data models for end-of-treatment (EOT) outcome prediction. Plots depict SHAP value trajectories of variables not shown in Figure 3. Patients were stratified by the distribution of the corresponding feature values at each time point: continuous variables were divided into quantile-based strata (bottom 25% and top 25%), while binary and ordinal variables were stratified into two groups (negative vs positive; lowest vs highest level). Lines represent the median SHAP value, and shaded bands indicate the interquartile range (25–75%) of SHAP values within each stratum at a given time point. Line and band colours encode the feature value strata (see legend). The horizontal dashed line denotes zero contribution to the model prediction. Positive SHAP values indicate increased contribution towards Unfavourable label whereas negative values indicate decreased contribution. B.: binary; Ord.: ordinal; Con.: continuous; ce: clinical event; LJ-culture: Löwenstein-Jensen culture; BMI: body mass index.

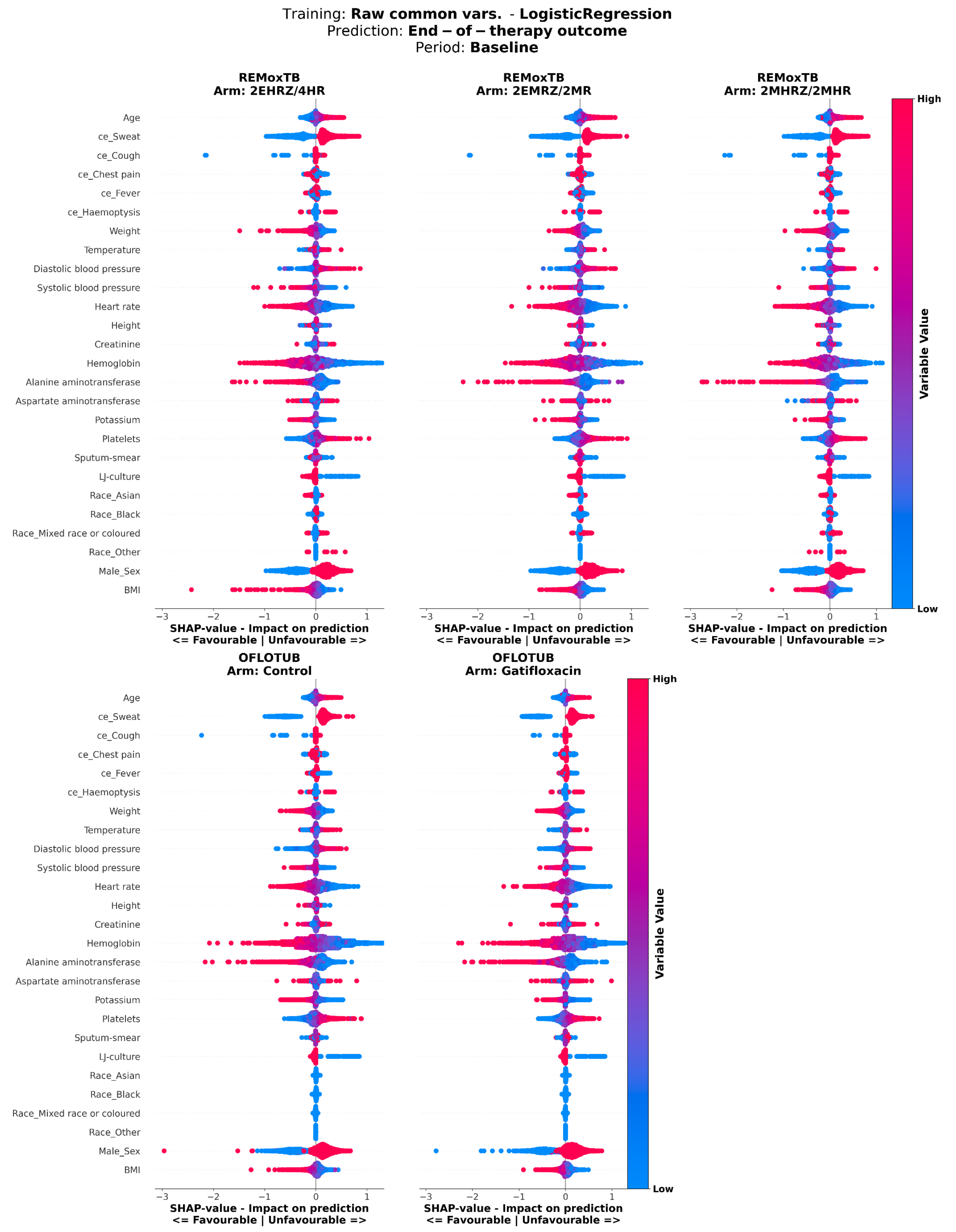


**Supplementary Figure 7**: **SHAP value analysis of logistic regresssion model predicting end-of-therapy outcome, using raw common variable data at baseline.** SHAP values of logistic regression models predicting end-of-therapy outcome, trained on common clinical and treatment variables across studies at baseline. SHAP values were calculated on test set patients. The y-axis shows variables included in the model; the x-axis shows SHAP values, where positive values indicate greater contribution to the positive label (Unfavourable), and negative values indicate lower contribution. Each dot represents a patient, with dot colour indicating the normalized value of the variable for that patient. Panels show patients stratified by treatment arm, with arm and study name indicated above each panel. mh.: medical history; ce: clinical event, BMI: body mass index.

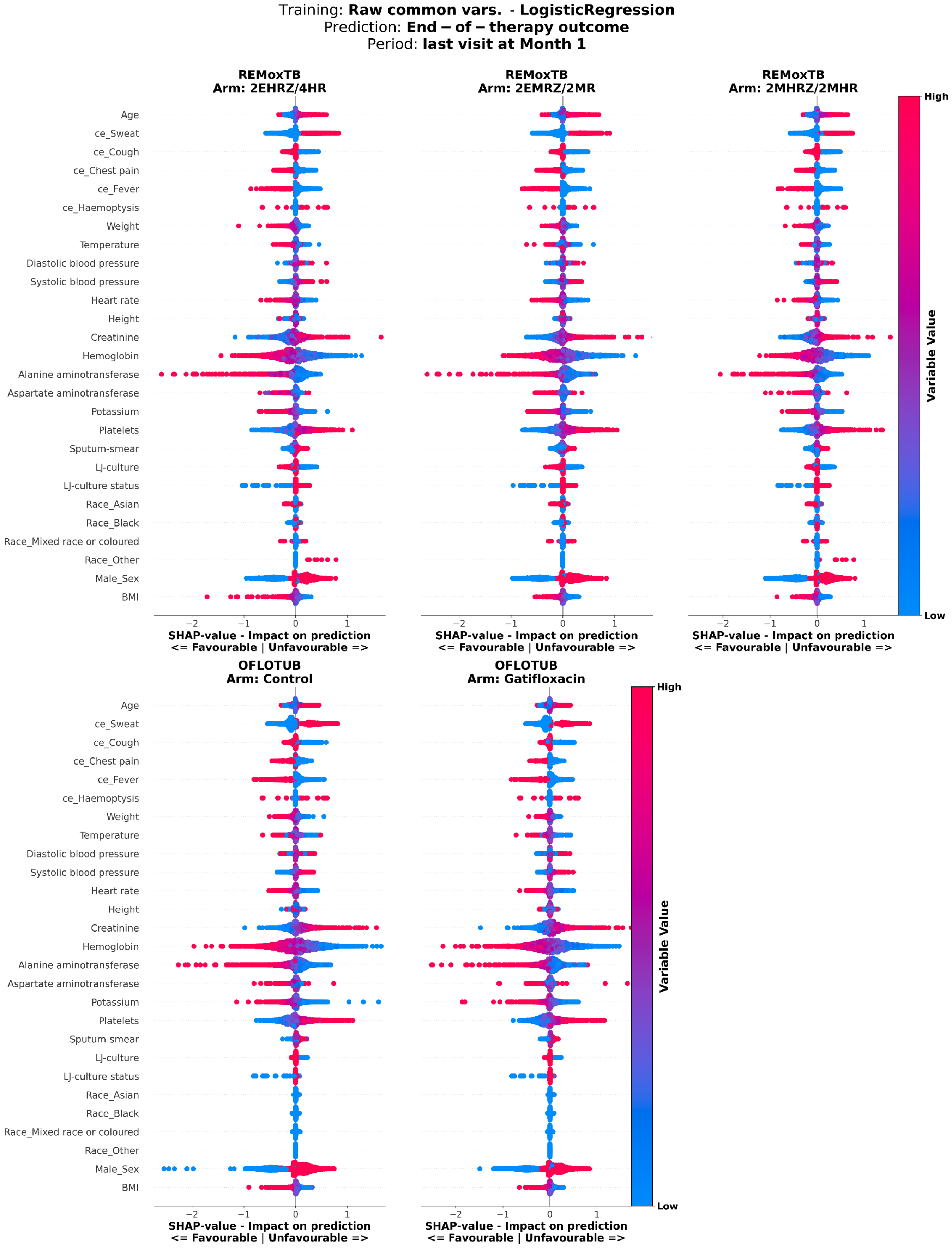


**Supplementary Figure 8**: **SHAP value analysis of logistic regresssion model predicting end-of-therapy outcome, using raw common variable data from last visit at month 1.** SHAP values of logistic regression models predicting end-of-therapy outcome, trained on common clinical and treatment variables across studies from the last visit at month 1. SHAP values were calculated on test set patients. The y-axis shows variables included in the model; the x-axis shows SHAP values, where positive values indicate greater contribution to the positive label (Unfavourable), and negative values indicate lower contribution. Each dot represents a patient, with dot colour indicating the normalized value of the variable for that patient. Panels show patients stratified by treatment arm, with arm and study name indicated above each panel. mh.: medical history; ce: clinical event. , BMI: body mass index.

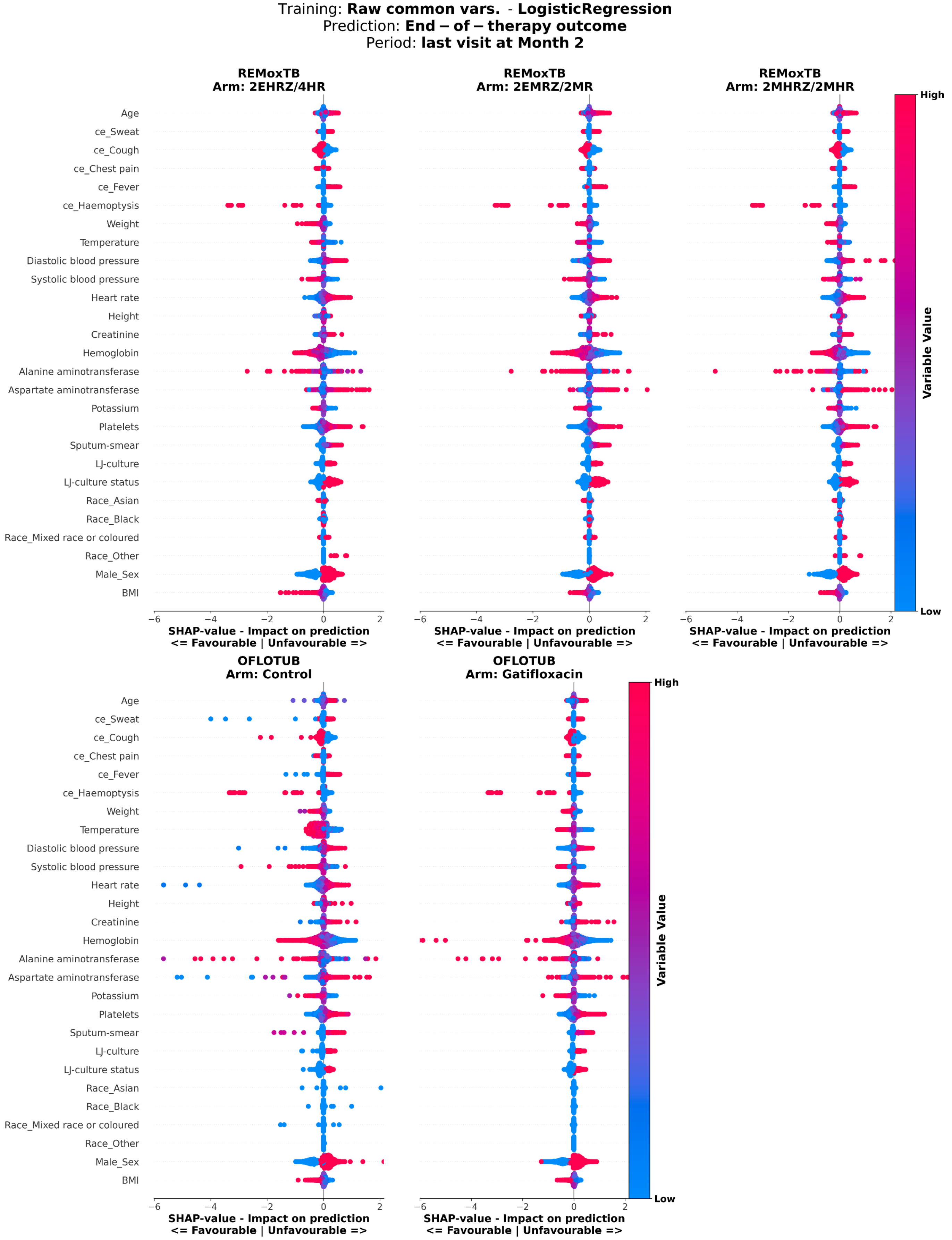


**Supplementary Figure 9**: **SHAP value analysis of logistic regresssion model predicting end-of-therapy outcome, using raw common variable data from last visit at month 2.** SHAP values of logistic regression models predicting end-of-therapy outcome, trained on common clinical and treatment variables across studies from the last visit at month 2. SHAP values were calculated on test set patients. The y-axis shows variables included in the model; the x-axis shows SHAP values, where positive values indicate greater contribution to the positive label (Unfavourable), and negative values indicate lower contribution. Each dot represents a patient, with dot colour indicating the normalized value of the variable for that patient. Panels show patients stratified by treatment arm, with arm and study name indicated above each panel. mh.: medical history; ce: clinical event. , BMI: body mass index.

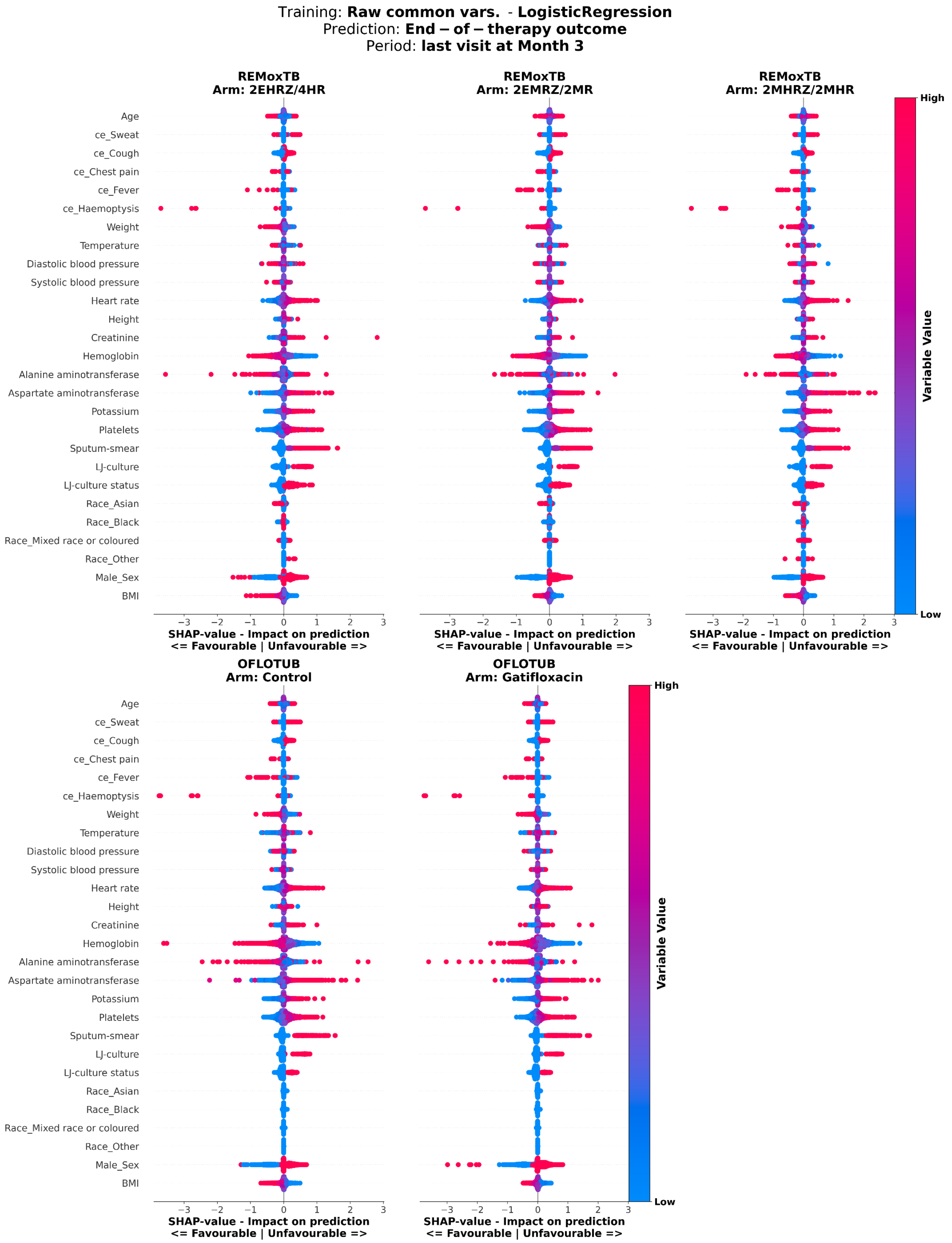


**Supplementary Figure 10**: **SHAP value analysis of logistic regresssion model predicting end-of-therapy outcome, using raw common variable data from last visit at month 3.** SHAP values of logistic regression models predicting end-of-therapy outcome, trained on common clinical and treatment variables across studies from the last visit at month 3. SHAP values were calculated on test set patients. The y-axis shows variables included in the model; the x-axis shows SHAP values, where positive values indicate greater contribution to the positive label (Unfavourable), and negative values indicate lower contribution. Each dot represents a patient, with dot colour indicating the normalized value of the variable for that patient. Panels show patients stratified by treatment arm, with arm and study name indicated above each panel. mh.: medical history; ce: clinical event. , BMI: body mass index.

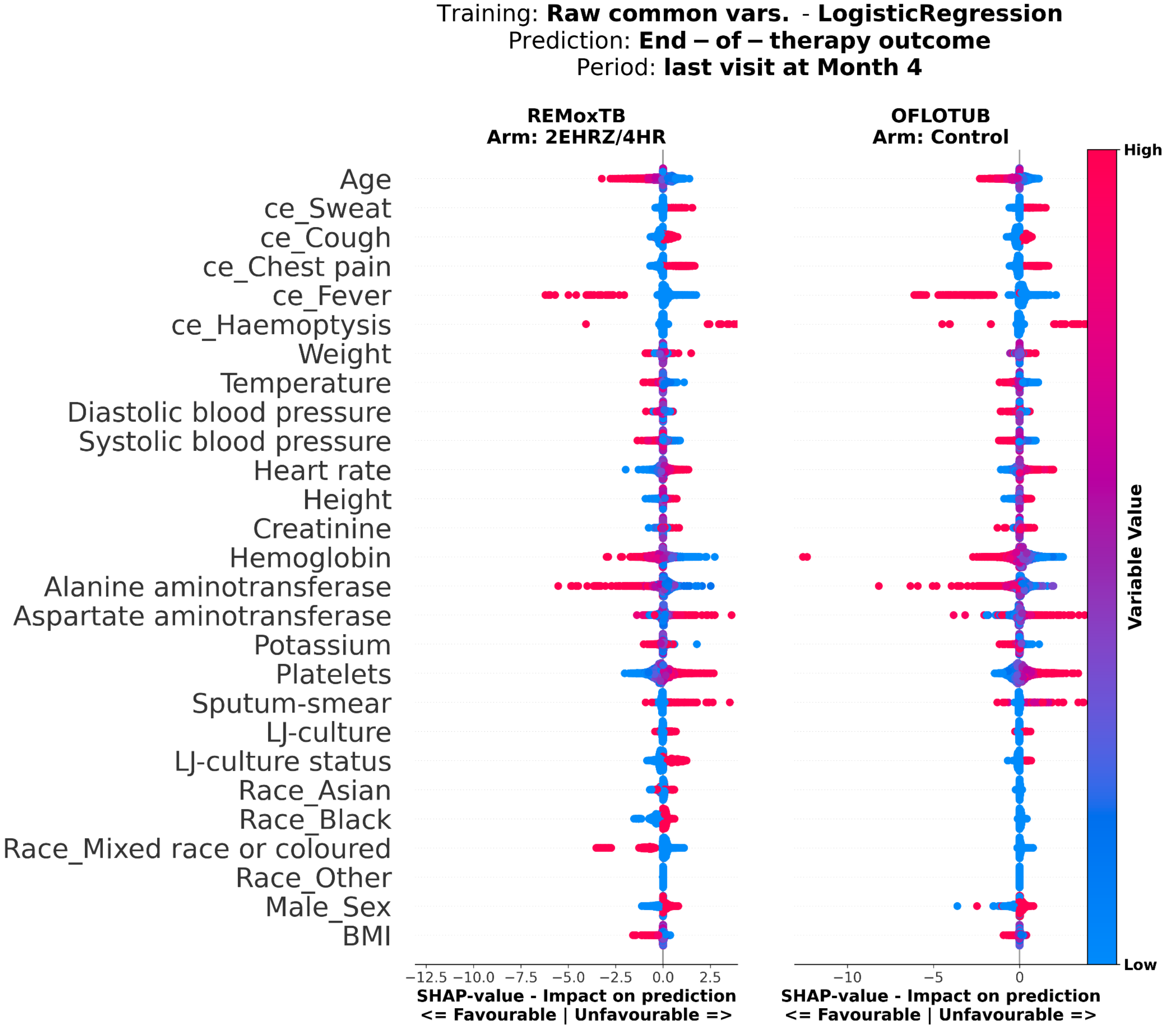


**Supplementary Figure 11**: **SHAP value analysis of logistic regresssion model predicting end-of-therapy outcome, using raw common variable data from last visit at month 4.** SHAP values of logistic regression models predicting end-of-therapy outcome, trained on common clinical and treatment variables across studies from the last visit at month 4. SHAP values were calculated on test set patients. The y-axis shows variables included in the model; the x-axis shows SHAP values, where positive values indicate greater contribution to the positive label (Unfavourable), and negative values indicate lower contribution. Each dot represents a patient, with dot colour indicating the normalized value of the variable for that patient. Panels show patients stratified by treatment arm, with arm and study name indicated above each panel. mh.: medical history; ce: clinical event. , BMI: body mass index.

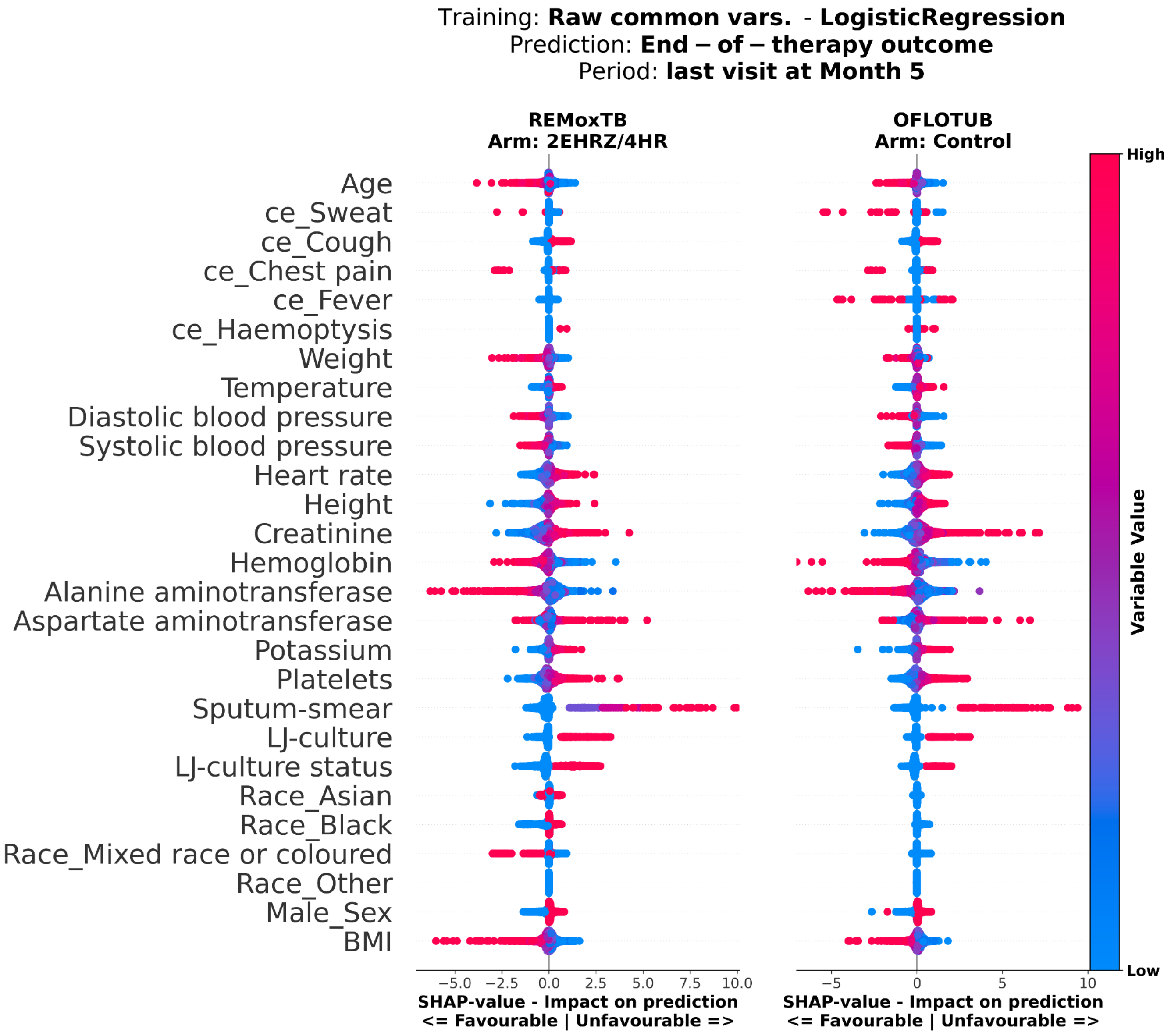


**Supplementary Figure 12**: **SHAP value analysis of logistic regresssion model predicting end-of-therapy outcome, using raw common variable data from last visit at month 5.** SHAP values of logistic regression models predicting end-of-therapy outcome, trained on common clinical and treatment variables across studies from the last visit at month 5. SHAP values were calculated on test set patients. The y-axis shows variables included in the model; the x-axis shows SHAP values, where positive values indicate greater contribution to the positive label (Unfavourable), and negative values indicate lower contribution. Each dot represents a patient, with dot colour indicating the normalized value of the variable for that patient. Panels show patients stratified by treatment arm, with arm and study name indicated above each panel. mh.: medical history; ce: clinical event. , BMI: body mass index.

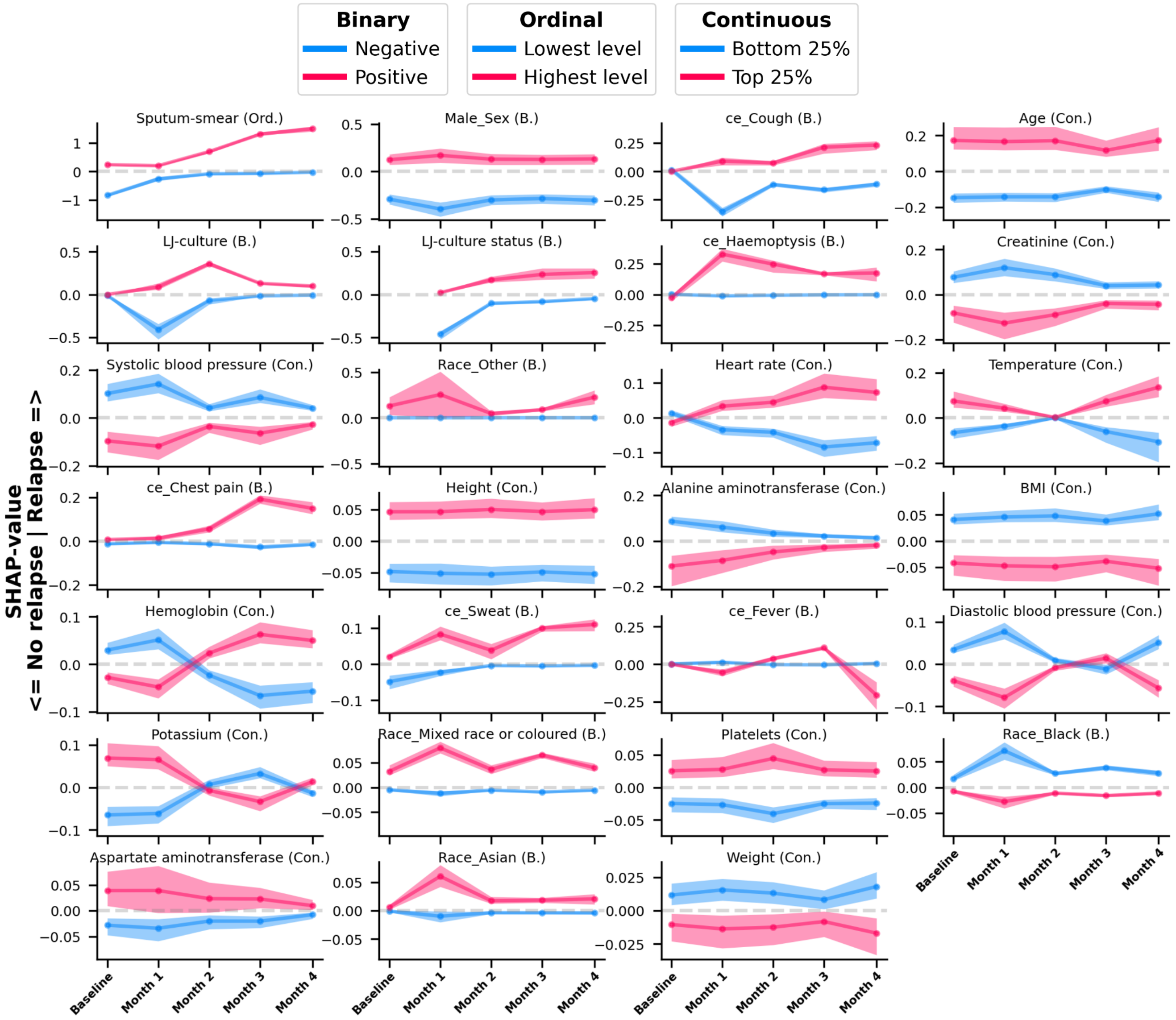


**Supplementary Figure 13**: **Population-level SHAP value trajectories across therapy periods for all clinical variables used for relapse prediction**. Population-level SHAP value trajectories from last-visit raw-data models for relapse prediction. Plots depict SHAP value trajectories of variables not shown in Figure 3. Patients were stratified by the distribution of the corresponding feature values at each time point: continuous variables were divided into quantile-based strata (bottom 25% and top 25%), while binary and ordinal variables were stratified into two groups (negative vs positive; lowest vs highest level). Lines represent the median SHAP value, and shaded bands indicate the interquartile range (25–75%) of SHAP values within each stratum at a given time point. Line and band colours encode the feature value strata (see legend). The horizontal dashed line denotes zero contribution to the model prediction. Positive SHAP values indicate increased contribution towards Relapse label whereas negative values indicate decreased contribution. B.: binary; Ord.: ordinal; Con.: continuous; ce: clinical event; LJ-culture: Löwenstein-Jensen culture; BMI: body mass index.

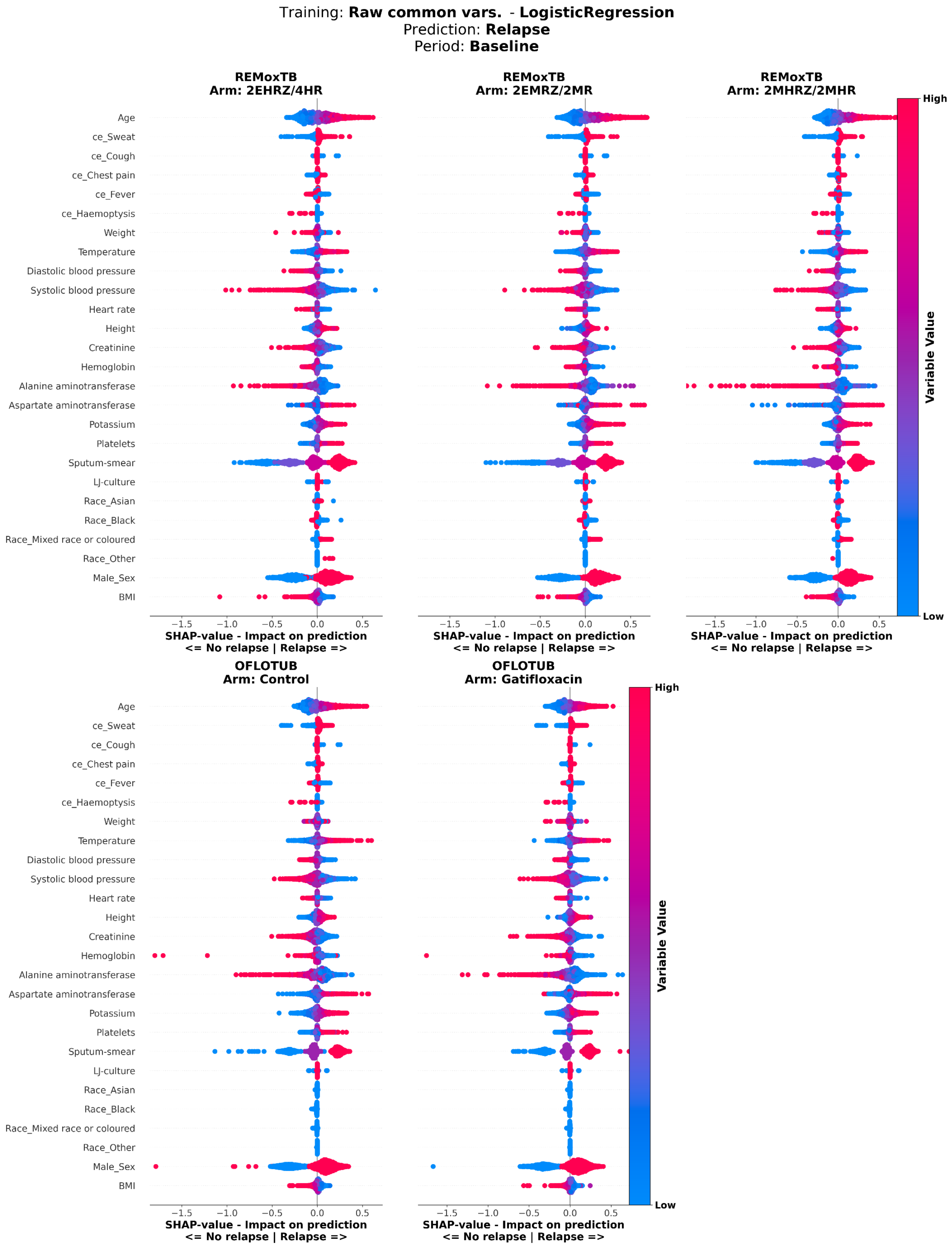


**Supplementary Figure 14**: **SHAP value analysis of logistic regresssion model predicting relapse, using raw common variable data at baseline.** SHAP values of logistic regression models predicting relapse, trained on common clinical and treatment variables across studies at baseline. SHAP values were calculated on test set patients. The y-axis shows variables included in the model; the x-axis shows SHAP values, where positive values indicate greater contribution to the positive label (Relapse), and negative values indicate lower contribution. Each dot represents a patient, with dot colour indicating the normalized value of the variable for that patient. Panels show patients stratified by treatment arm, with arm and study name indicated above each panel. mh.: medical history; ce: clinical event, BMI: body mass index.

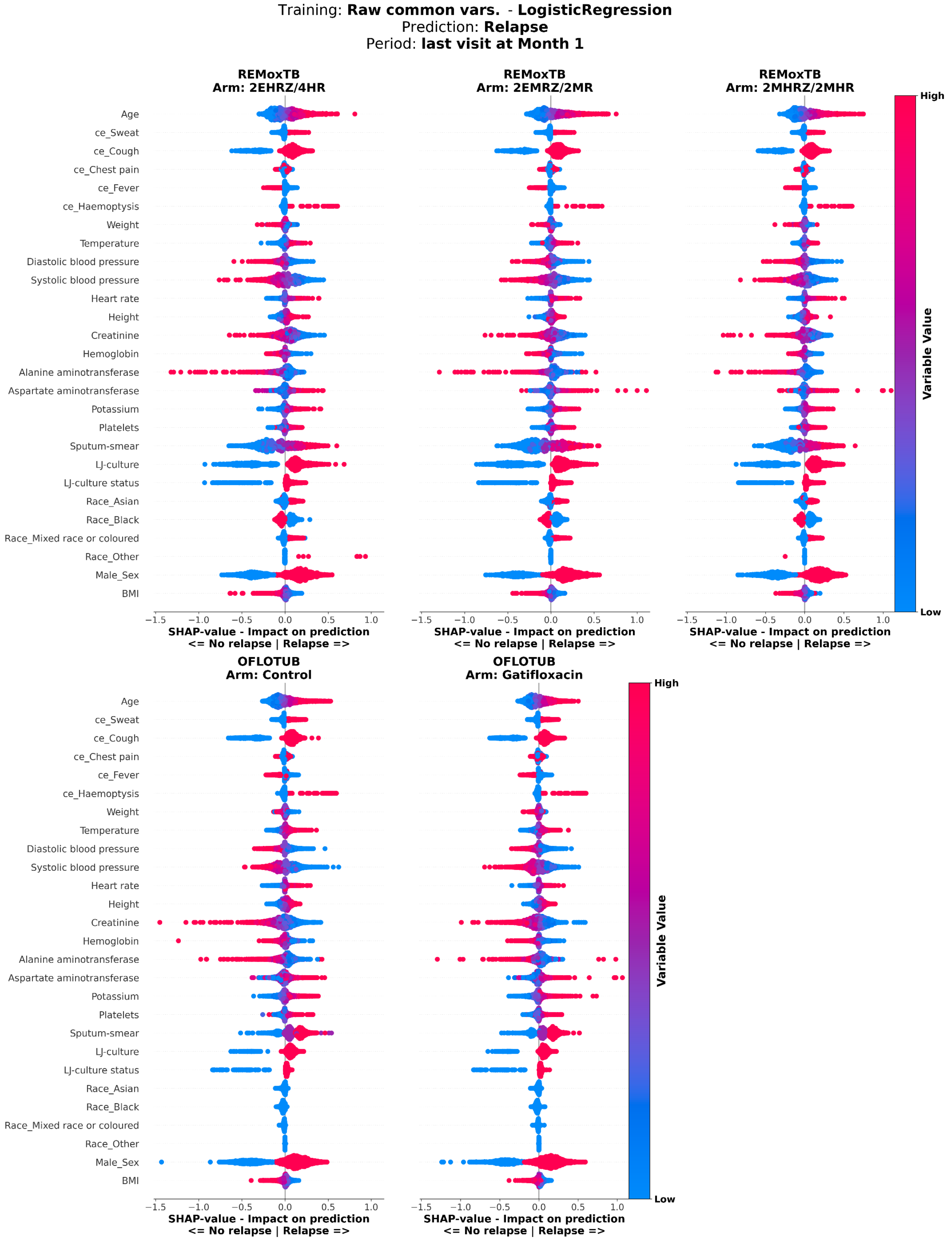


**Supplementary Figure 15**: **SHAP value analysis of logistic regresssion model predicting relapse, using raw common variable data from last visit at month 1.** SHAP values of logistic regression models predicting relapse, trained using on common clinical and treatment variables across studies from the last visit at month 1. SHAP values were calculated on test set patients. The y-axis shows variables included in the model; the x-axis shows SHAP values, where positive values indicate greater contribution to the positive label (Relapse), and negative values indicate lower contribution. Each dot represents a patient, with dot colour indicating the normalized value of the variable for that patient. Panels show patients stratified by treatment arm, with arm and study name indicated above each panel. mh.: medical history; ce: clinical event. , BMI: body mass index.

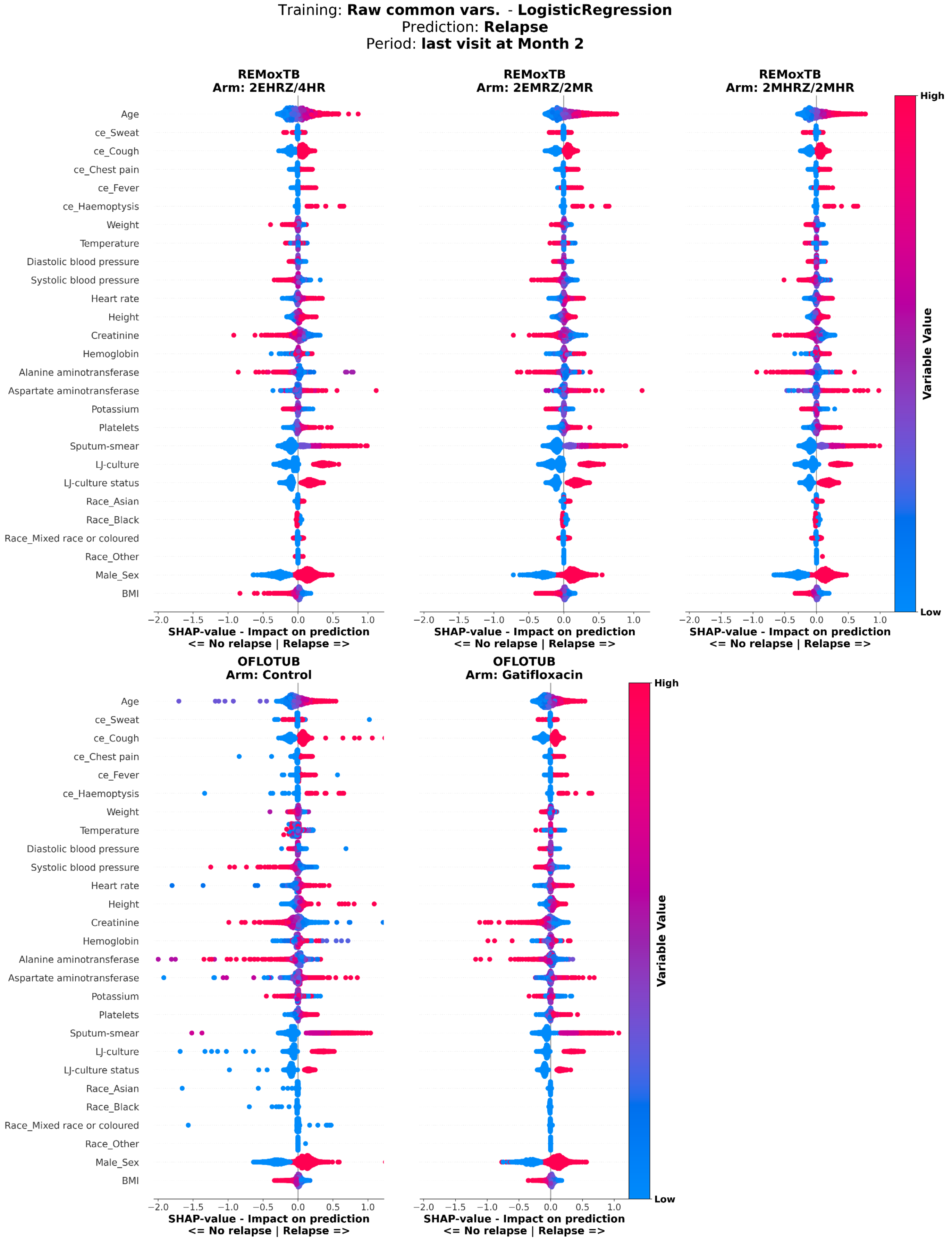


**Supplementary Figure 16**: **SHAP value analysis of logistic regresssion model predicting relapse, using raw common variable data from last visit at month 2.** SHAP values of logistic regression models predicting relapse, trained using on common clinical and treatment variables across studies from the last visit at month 2. SHAP values were calculated on test set patients. The y-axis shows variables included in the model; the x-axis shows SHAP values, where positive values indicate greater contribution to the positive label (Relapse), and negative values indicate lower contribution. Each dot represents a patient, with dot colour indicating the normalized value of the variable for that patient. Panels show patients stratified by treatment arm, with arm and study name indicated above each panel. mh.: medical history; ce: clinical event. , BMI: body mass index.

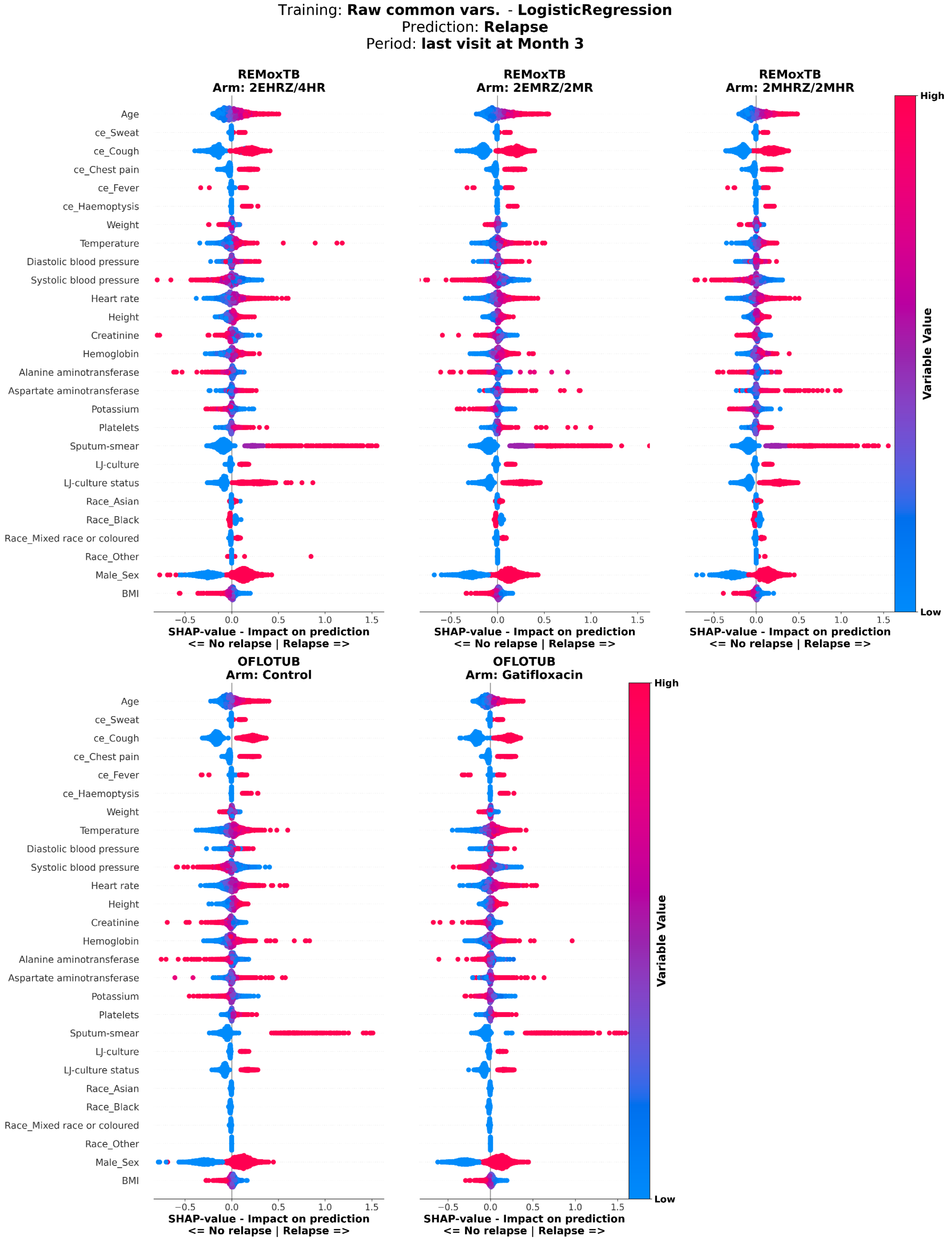


**Supplementary Figure 17**: **SHAP value analysis of logistic regresssion model predicting relapse, using raw common variable data from last visit at month 3.** SHAP values of logistic regression models predicting relapse, trained using on common clinical and treatment variables across studies from the last visit at month 3. SHAP values were calculated on test set patients. The y-axis shows variables included in the model; the x-axis shows SHAP values, where positive values indicate greater contribution to the positive label (Relapse), and negative values indicate lower contribution. Each dot represents a patient, with dot colour indicating the normalized value of the variable for that patient. Panels show patients stratified by treatment arm, with arm and study name indicated above each panel. mh.: medical history; ce: clinical event. , BMI: body mass index.

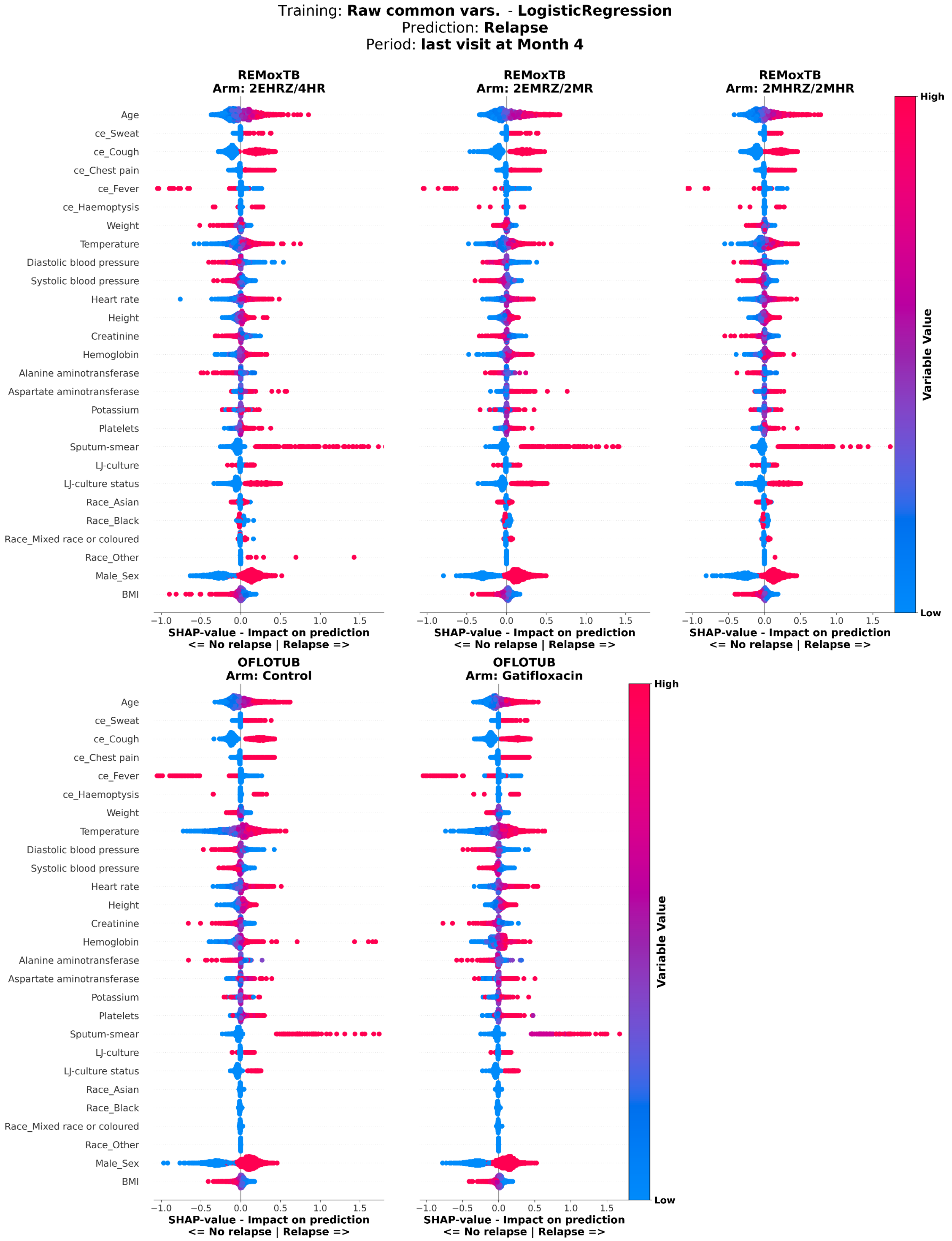


**Supplementary Figure 18**: **SHAP value analysis of logistic regresssion model predicting relapse, using raw common variable data from last visit at month 4.** SHAP values of logistic regression models predicting relapse, trained using on common clinical and treatment variables across studies from the last visit at month 4. SHAP values were calculated on test set patients. The y-axis shows variables included in the model; the x-axis shows SHAP values, where positive values indicate greater contribution to the positive label (Relapse), and negative values indicate lower contribution. Each dot represents a patient, with dot colour indicating the normalized value of the variable for that patient. Panels show patients stratified by treatment arm, with arm and study name indicated above each panel. mh.: medical history; ce: clinical event. , BMI: body mass index.

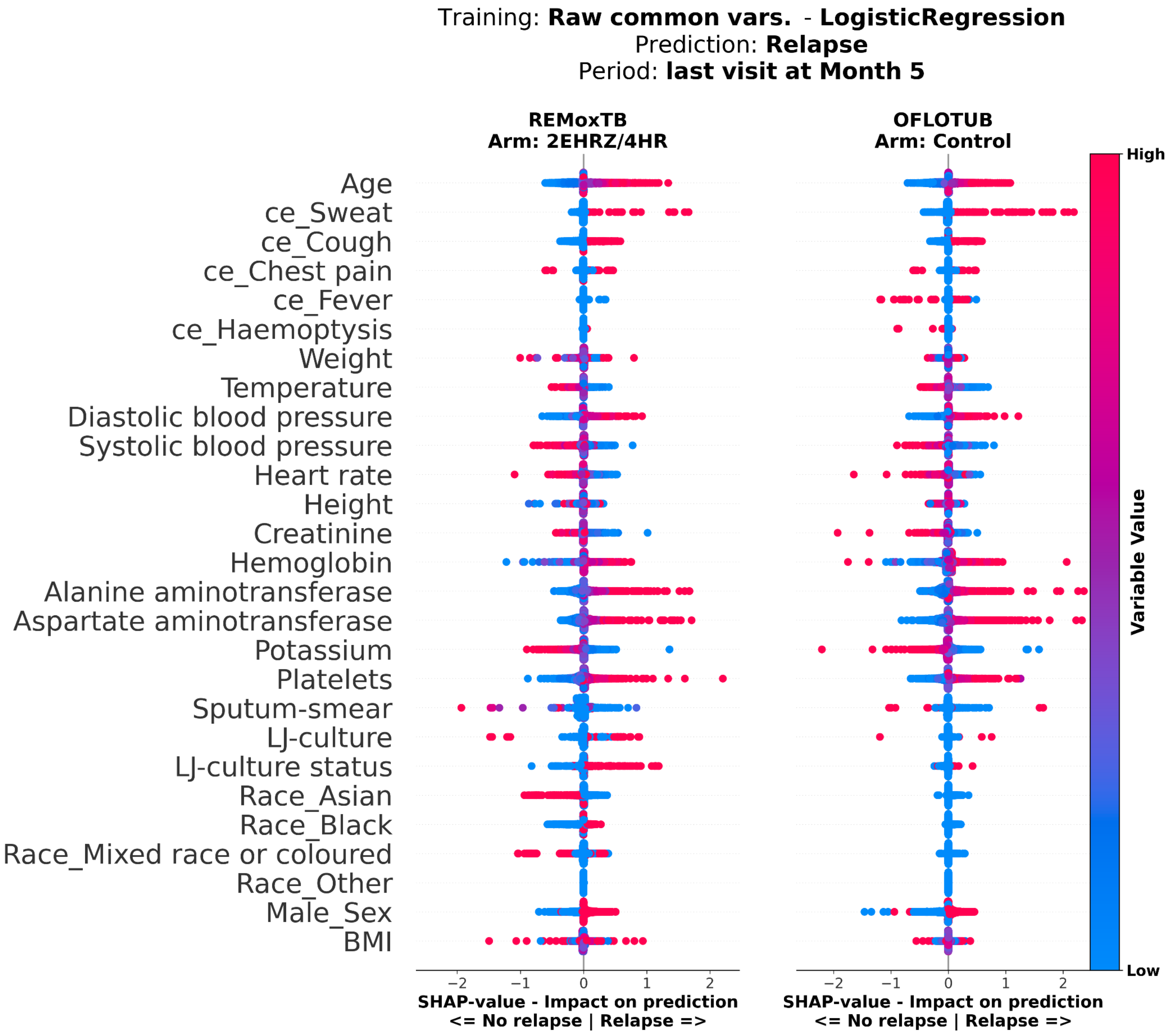


**Supplementary Figure 19**: **SHAP value analysis of logistic regresssion model predicting relapse, using raw common variable data from last visit at month 5.** SHAP values of logistic regression models predicting relapse, trained using on common clinical and treatment variables across studies from the last visit at month 5. SHAP values were calculated on test set patients. The y-axis shows variables included in the model; the x-axis shows SHAP values, where positive values indicate greater contribution to the positive label (Relapse), and negative values indicate lower contribution. Each dot represents a patient, with dot colour indicating the normalized value of the variable for that patient. Panels show patients stratified by treatment arm, with arm and study name indicated above each panel. mh.: medical history; ce: clinical event. , BMI: body mass index.

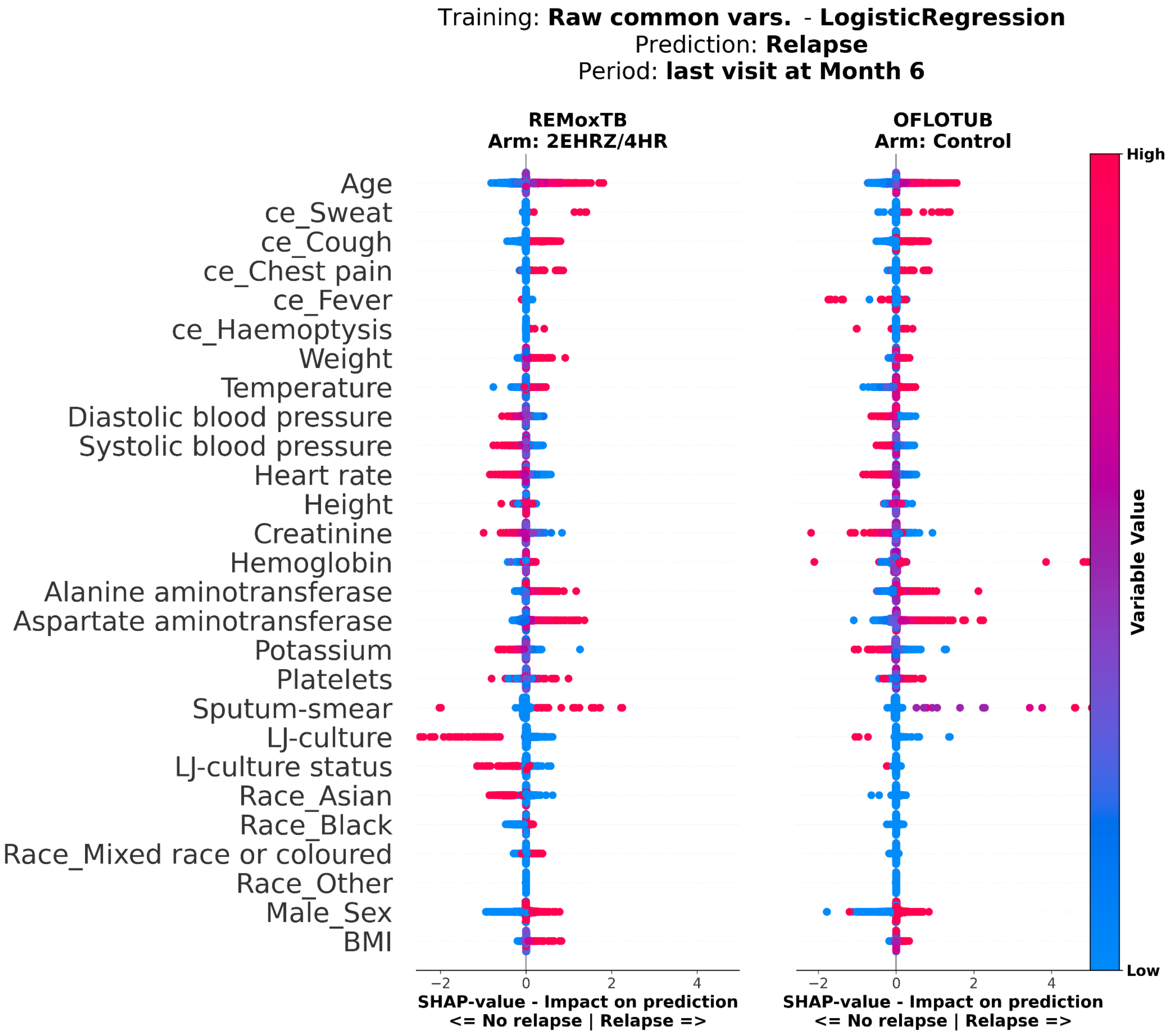


**Supplementary Figure 20**: **SHAP value analysis of logistic regresssion model predicting relapse, using raw common variable data from last visit at month 6.** SHAP values of logistic regression models predicting relapse, trained using on common clinical and treatment variables across studies from the last visit at month 6. SHAP values were calculated on test set patients. The y-axis shows variables included in the model; the x-axis shows SHAP values, where positive values indicate greater contribution to the positive label (Relapse), and negative values indicate lower contribution. Each dot represents a patient, with dot colour indicating the normalized value of the variable for that patient. Panels show patients stratified by treatment arm, with arm and study name indicated above each panel. mh.: medical history; ce: clinical event. , BMI: body mass index.

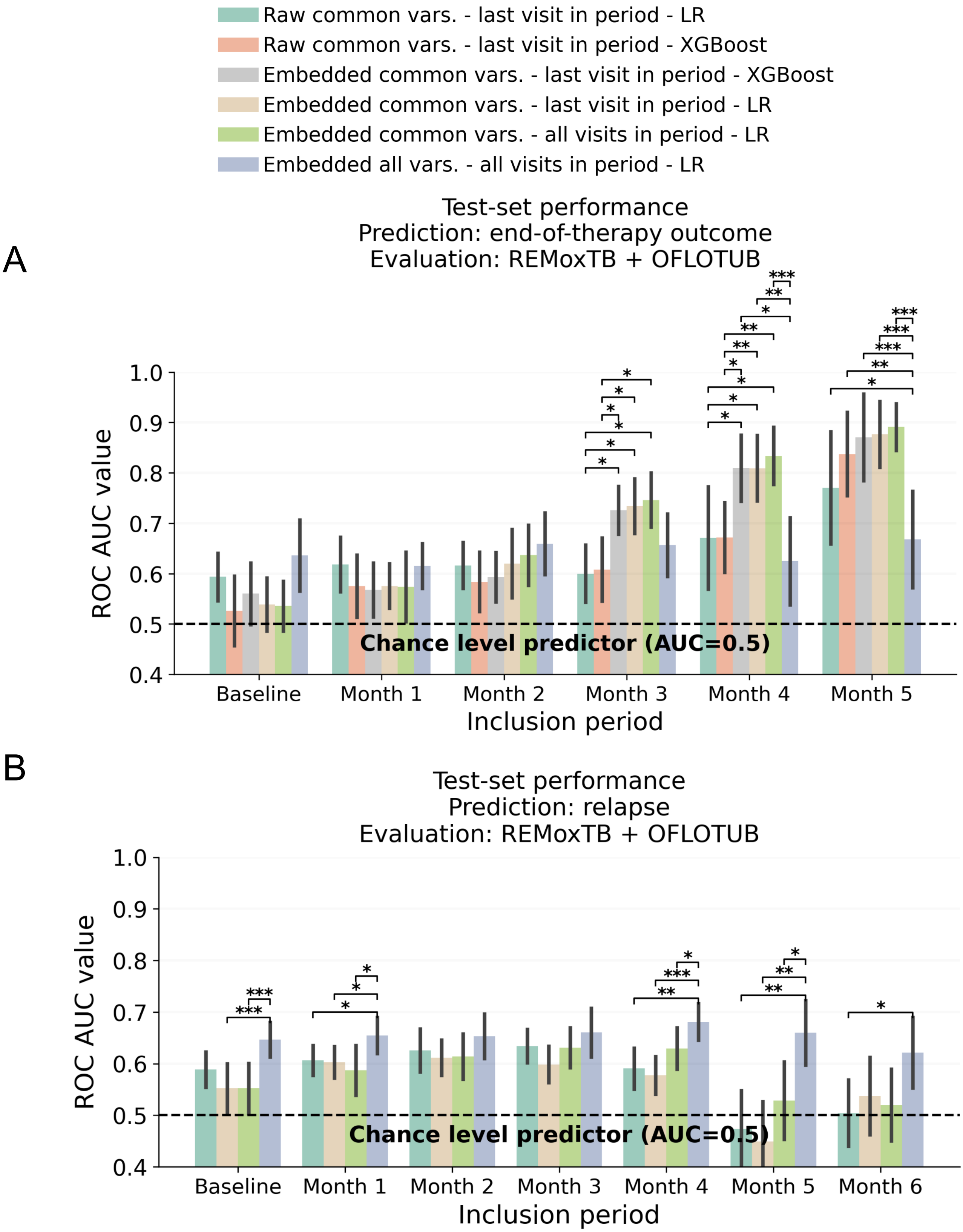


**Supplementary Figure 21: Comparative end-of-therapy outcome and relapse prediction using raw data and LLM embeddings.** Predictive performance (ROC-AUC) of XGBoost and logistic regression (LR) models trained on four input setups: raw tabular data of common clinical and treatment variables across studies, taken from the last visit in each period ("Raw common vars. – last visit in period"); LLM embeddings of the same variables and visits ("Embedded common vars. – last visit in period"); LLM embeddings of common variables containing all visits up to each period cutoff ("Embedded common vars. – all visits in period"); LLM embeddings incorporating all variables and all visits up to each period cutoff ("Embedded all vars. – all visits in period"). Subplot (A) shows results of end-of-therapy outcome, (B) results of relapse prediction. Legend above shows colour coding of the input setup-model combinations used for prediction. Bars represent mean ROC-AUC across 25 test set splits; error bars indicate standard deviation. Corrected resampled t-tests (Nadeau & Bengio, 2003) were used to compare setups within periods, accounting for the correlation between performance estimates induced by overlapping test sets across the 25 random train-test splits, with only significant comparisons shown after Benjamini-Hochberg correction (*: adj. $p<0.05$, **: adj. $p<0.01$, ***: adj. $p<0.001$). The dashed line indicates chance-level performance (AUC = 0.5)

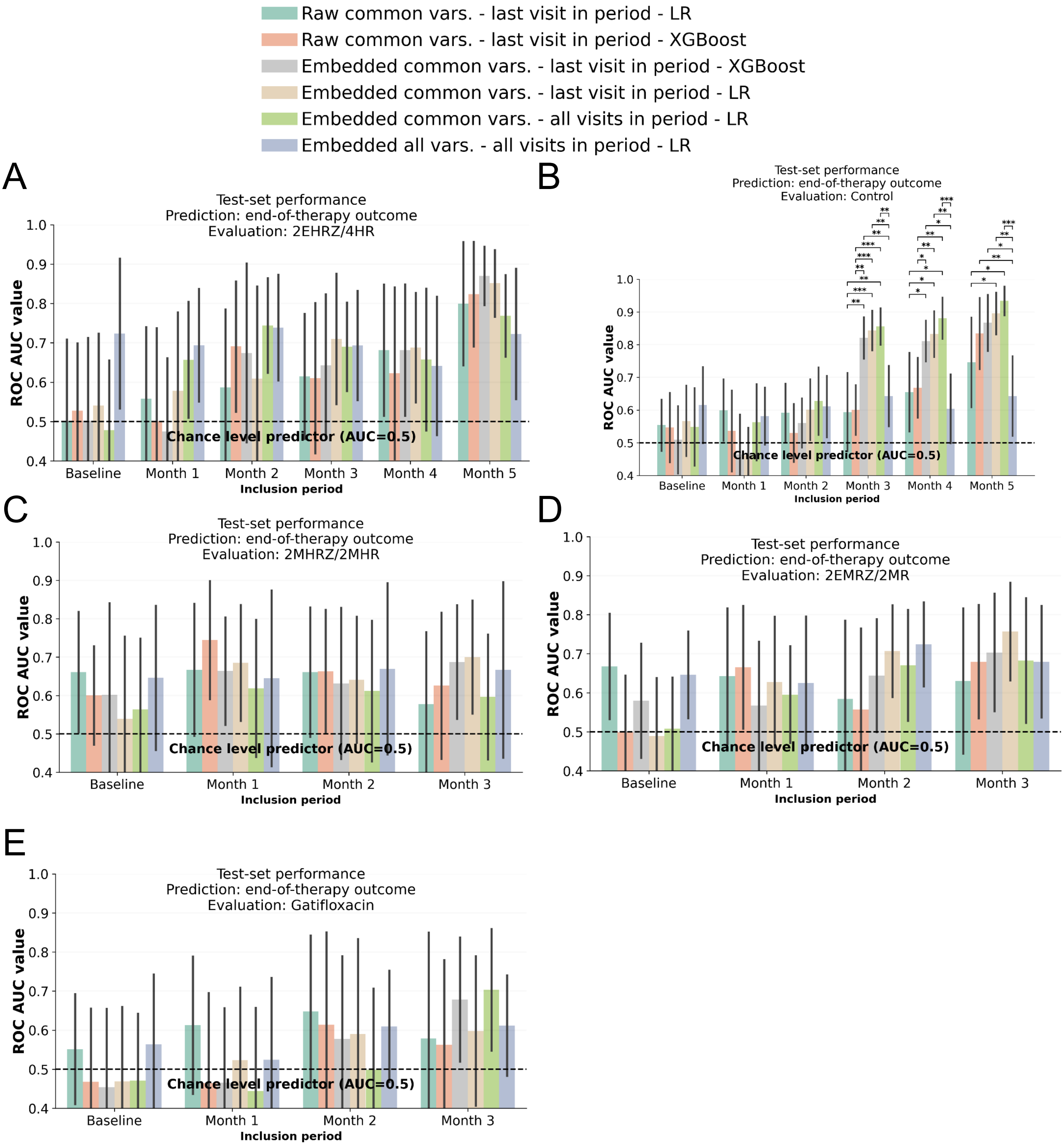


**Supplementary Figure 22: Within-arm comparison of end-of-therapy outcome prediction using raw data and LLM embeddings.** Predictive performance (ROC-AUC) of XGBoost and logistic regression (LR) models predicting end-of-therapy (EOT) outcome trained on four input setups: raw tabular data of common clinical and treatment variables across studies, taken from the last visit in each period ("Raw common vars. – last visit in period"); LLM embeddings of the same variables and visits ("Embedded common vars. – last visit in period"); LLM embeddings of common variables containing all visits up to each period cutoff ("Embedded common vars. – all visits in period"); LLM embeddings incorporating all variables and all visits up to each period cutoff ("Embedded all vars. – all visits in period"). (**A-E**) panels show the within-arm performance of models evaluated on patients from the corresponding treatment arms included in the analysis. Corresponding arm and study names are indicated above the subplots. Legend above shows colour coding of the input setup-model combinations used for prediction. Bars represent mean ROC-AUC across 25 test set splits; error bars indicate standard deviation. Corrected resampled t-tests (Nadeau & Bengio, 2003) were used to compare setups within periods, accounting for the correlation between performance estimates induced by overlapping test sets across the 25 random train-test splits, with only significant comparisons shown after Benjamini-Hochberg correction (*: adj. p<0.05, **: adj. p<0.01, ***: adj. p<0.001). The dashed line indicates chance-level performance (AUC = 0.5)

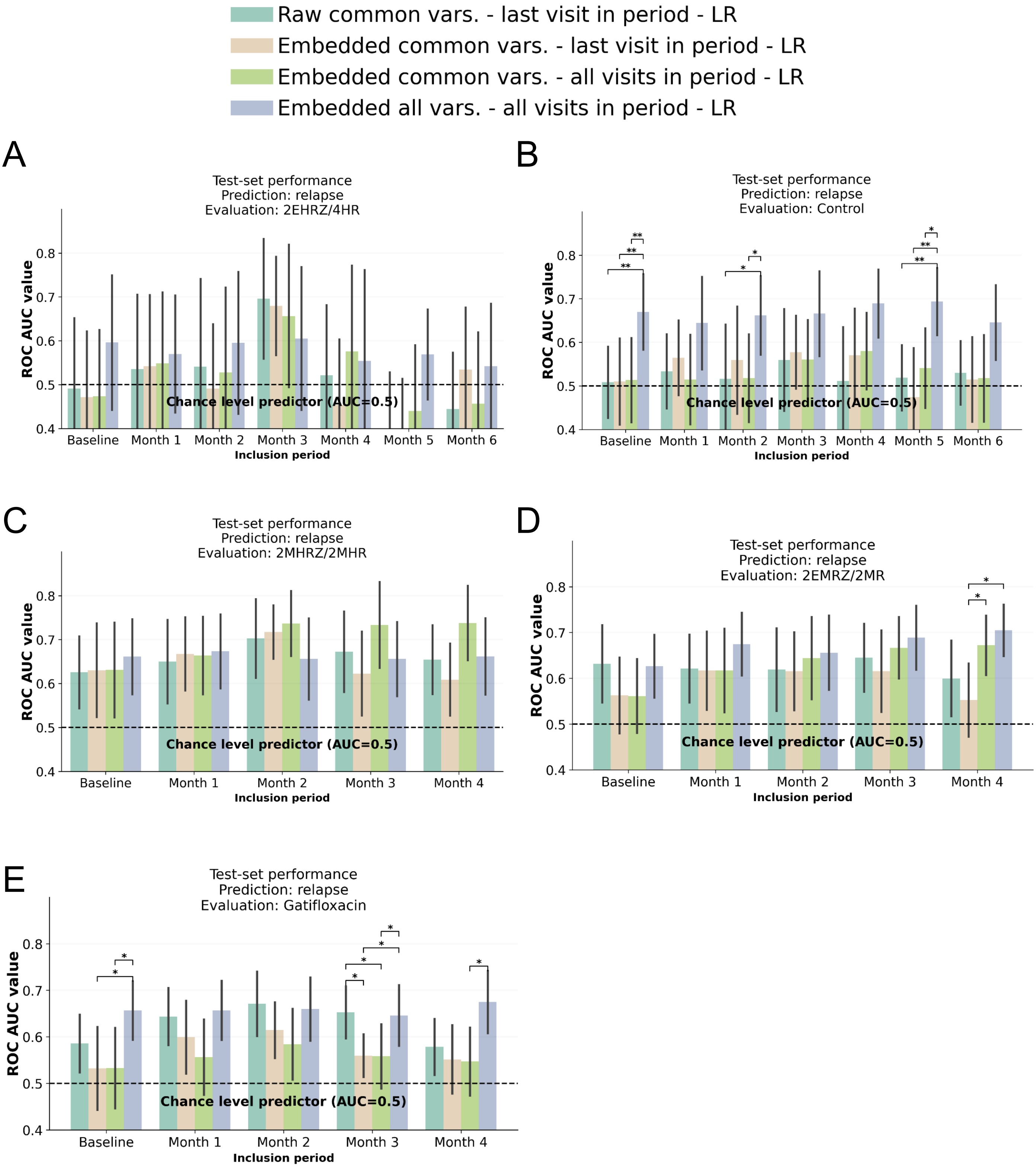


**Supplementary Figure 23: Within-arm comparison of relapse prediction using raw data and LLM embeddings.** Predictive performance (ROC-AUC) of XGBoost and logistic regression (LR) models predicting relapse trained on four input setups: raw tabular data of common clinical and treatment variables across studies, taken from the last visit in each period ("Raw common vars. – last visit in period"); LLM embeddings of the same variables and visits ("Embedded common vars. – last visit in period"); LLM embeddings of common variables containing all visits up to each period cutoff ("Embedded common vars. – all visits in period"); LLM embeddings incorporating all variables and all visits up to each period cutoff ("Embedded all vars. – all visits in period"). (**A-E**) panels show the within-arm performance of models evaluated on patients from the corresponding treatment arms included in the analysis. Corresponding arm and study names are indicated above the subplots. Legend above shows colour coding of the input setup-model combinations used for prediction. Bars represent mean ROC-AUC across 25 test set splits; error bars indicate standard deviation. Corrected resampled t-tests (Nadeau & Bengio, 2003) were used to compare setups within periods, accounting for the correlation between performance estimates induced by overlapping test sets across the 25 random train-test splits, with only significant comparisons shown after Benjamini-Hochberg correction (*: adj. p<0.05, **: adj. p<0.01, ***: adj. p<0.001). The dashed line indicates chance-level performance (AUC = 0.5)

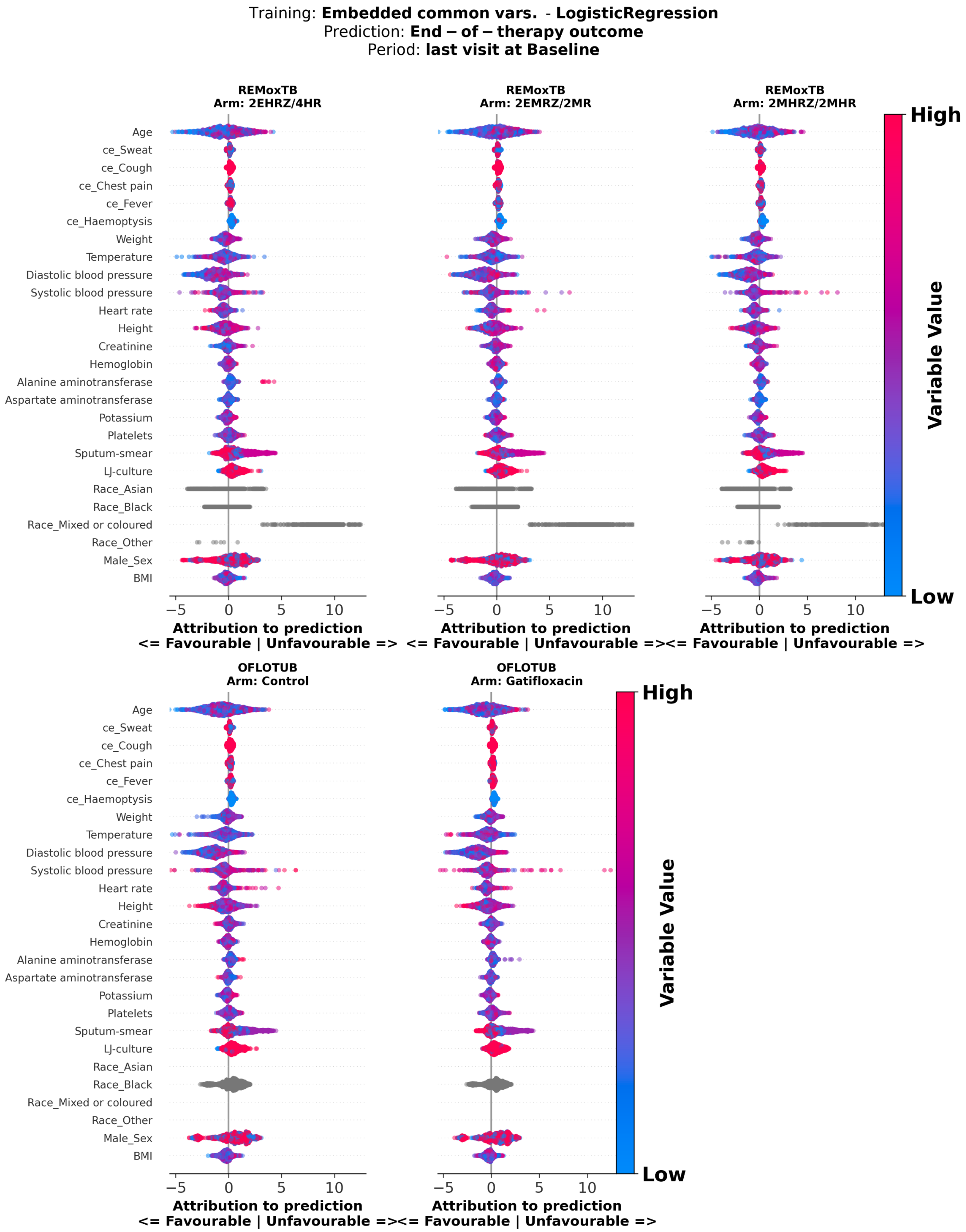


**Supplementary Figure 24**: **Attribution score analysis of logistic regresssion model predicting end-of-therapy outcome, using embeddings of common variable data at baseline.** Attribution scores of logistic regression models predicting end-of-therapy outcome, trained on embeddings of common clinical and treatment variables across studies at baseline. Attribution scores were calculated on test set patients. The y-axis shows variables included in the model; the x-axis shows attribution scores, where positive values indicate greater contribution to the positive label (Unfavourable), and negative values indicate lower contribution. Each dot represents a patient, with dot colour indicating the normalized value of the variable for that patient. Panels show patients stratified by treatment arm, with arm and study name indicated above each panel. mh.: medical history; ce: clinical event, BMI: body mass index.

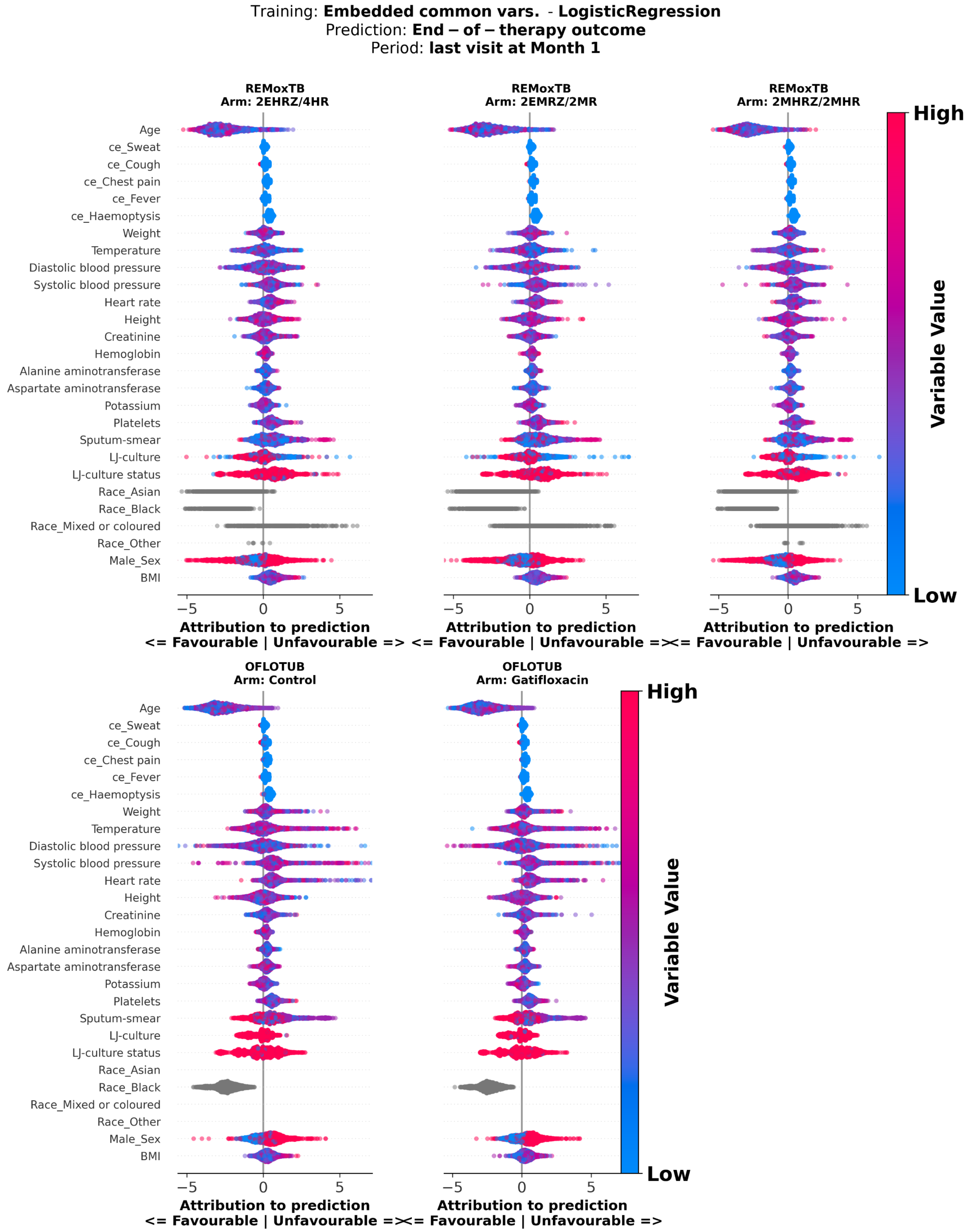


**Supplementary Figure 25**: **Attribution score analysis of logistic regresssion model predicting end-of-therapy outcome, using embeddings of common variable data from last visit at month 1.** Attribution scores of logistic regression models predicting end-of-therapy outcome, trained using on common clinical and treatment variables across studies from the last visit at month 1. Attribution scores were calculated on test set patients. The y-axis shows variables included in the model; the x-axis shows Attribution scores where positive values indicate greater contribution to the positive label (Unfavourable), and negative values indicate lower contribution. Each dot represents a patient, with dot colour indicating the normalized value of the variable for that patient. Panels show patients stratified by treatment arm, with arm and study name indicated above each panel. mh.: medical history; ce: clinical event, BMI: body mass index.

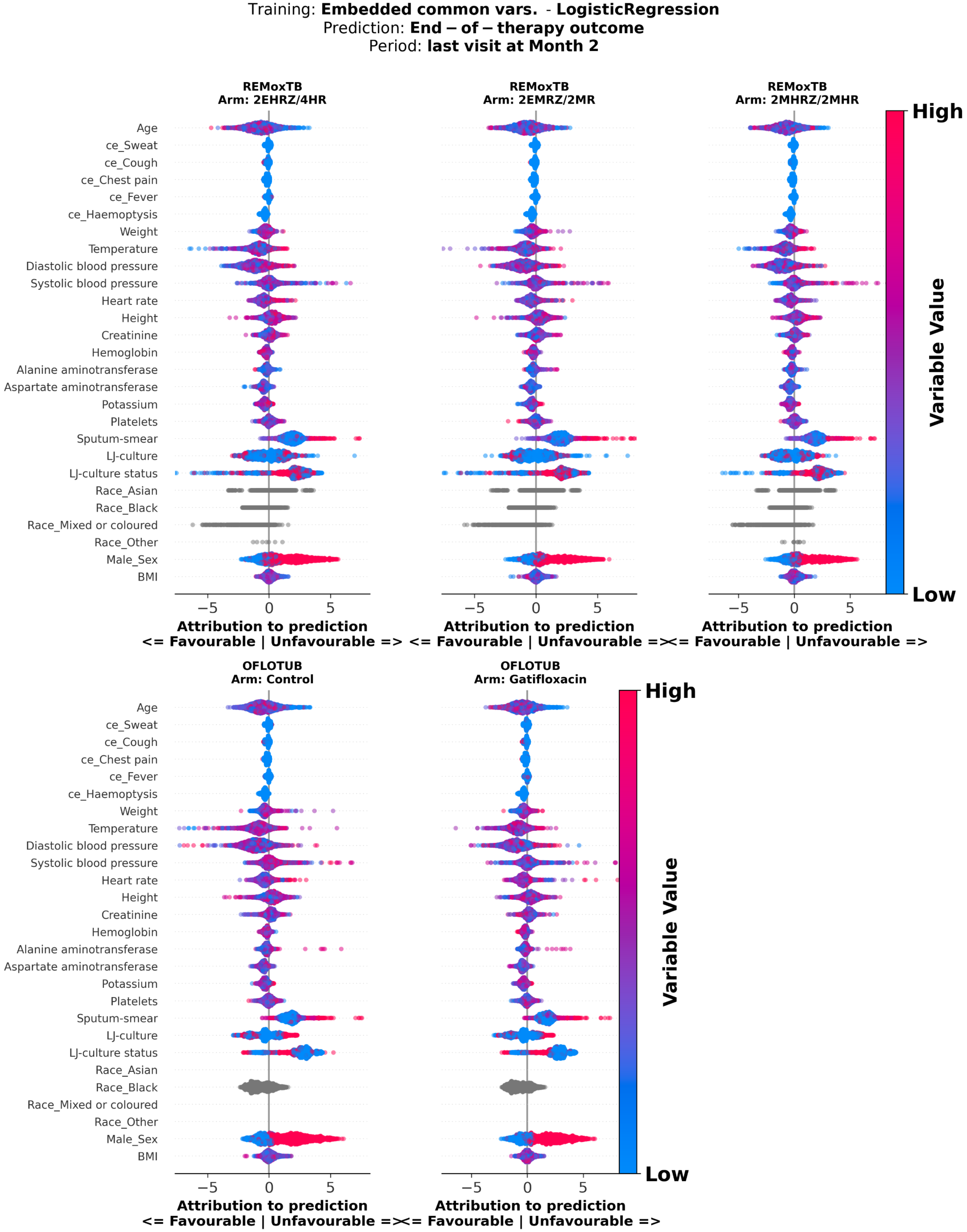


**Supplementary Figure 26**: **Attribution score analysis of logistic regresssion model predicting end-of-therapy outcome, using embeddings of common variable data from last visit at month 2.** Attribution scores of logistic regression models predicting end-of-therapy outcome, trained using on common clinical and treatment variables across studies from the last visit at month 2. Attribution scores were calculated on test set patients. The y-axis shows variables included in the model; the x-axis shows Attribution scores where positive values indicate greater contribution to the positive label (Unfavourable), and negative values indicate lower contribution. Each dot represents a patient, with dot colour indicating the normalized value of the variable for that patient. Panels show patients stratified by treatment arm, with arm and study name indicated above each panel. mh.: medical history; ce: clinical event, BMI: body mass index.

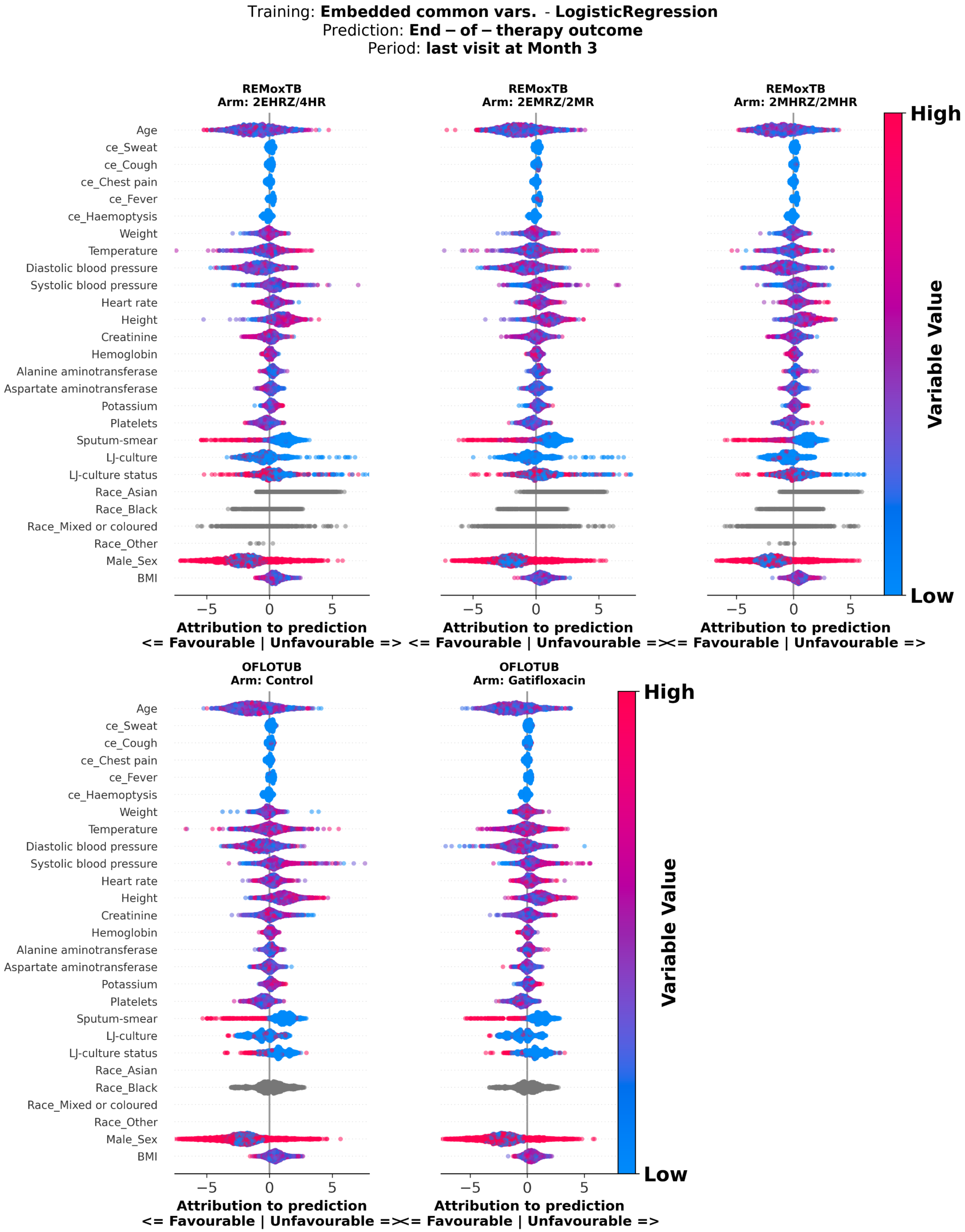


**Supplementary Figure 27**: **Attribution score analysis of logistic regresssion model predicting end-of-therapy outcome, using embeddings of common variable data from last visit at month 3.** Attribution scores of logistic regression models predicting end-of-therapy outcome, trained using on common clinical and treatment variables across studies from the last visit at month 3. Attribution scores were calculated on test set patients. The y-axis shows variables included in the model; the x-axis shows Attribution scores where positive values indicate greater contribution to the positive label (Unfavourable), and negative values indicate lower contribution. Each dot represents a patient, with dot colour indicating the normalized value of the variable for that patient. Panels show patients stratified by treatment arm, with arm and study name indicated above each panel. mh.: medical history; ce: clinical event, BMI: body mass index.

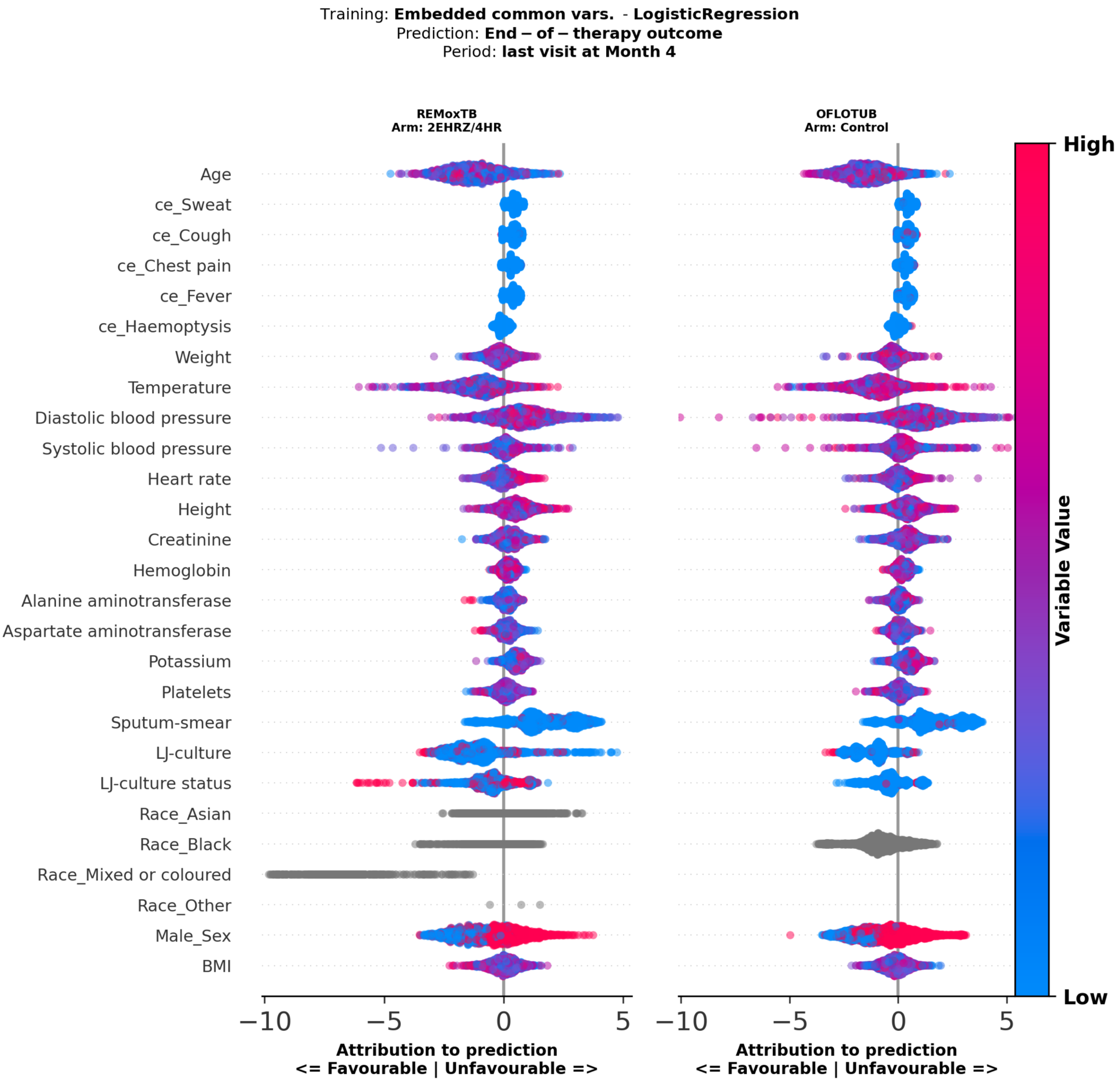


**Supplementary Figure 28**: **Attribution score analysis of logistic regresssion model predicting end-of-therapy outcome, using embeddings of common variable data from last visit at month 4.** Attribution scores of logistic regression models predicting end-of-therapy outcome, trained using on common clinical and treatment variables across studies from the last visit at month 4. Attribution scores were calculated on test set patients. The y-axis shows variables included in the model; the x-axis shows Attribution scores where positive values indicate greater contribution to the positive label (Unfavourable), and negative values indicate lower contribution. Each dot represents a patient, with dot colour indicating the normalized value of the variable for that patient. Panels show patients stratified by treatment arm, with arm and study name indicated above each panel. mh.: medical history; ce: clinical event, BMI: body mass index.

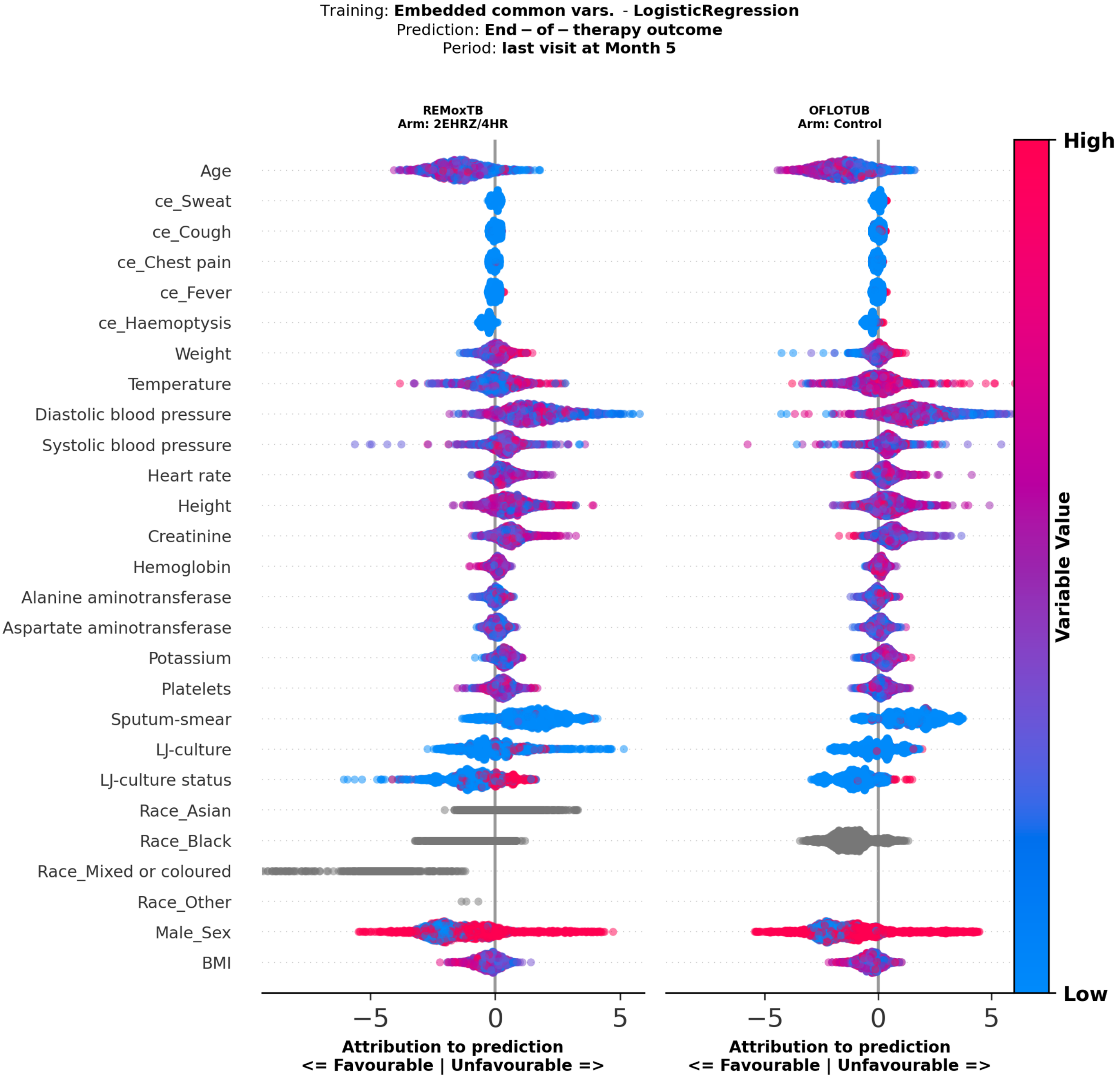


**Supplementary Figure 29**: **Attribution score analysis of logistic regresssion model predicting end-of-therapy outcome, using embeddings of common variable data from last visit at month 5.** Attribution scores of logistic regression models predicting end-of-therapy outcome, trained using on common clinical and treatment variables across studies from the last visit at month 5. Attribution scores were calculated on test set patients. The y-axis shows variables included in the model; the x-axis shows Attribution scores where positive values indicate greater contribution to the positive label (Unfavourable), and negative values indicate lower contribution. Each dot represents a patient, with dot colour indicating the normalized value of the variable for that patient. Panels show patients stratified by treatment arm, with arm and study name indicated above each panel. mh.: medical history; ce: clinical event, BMI: body mass index.

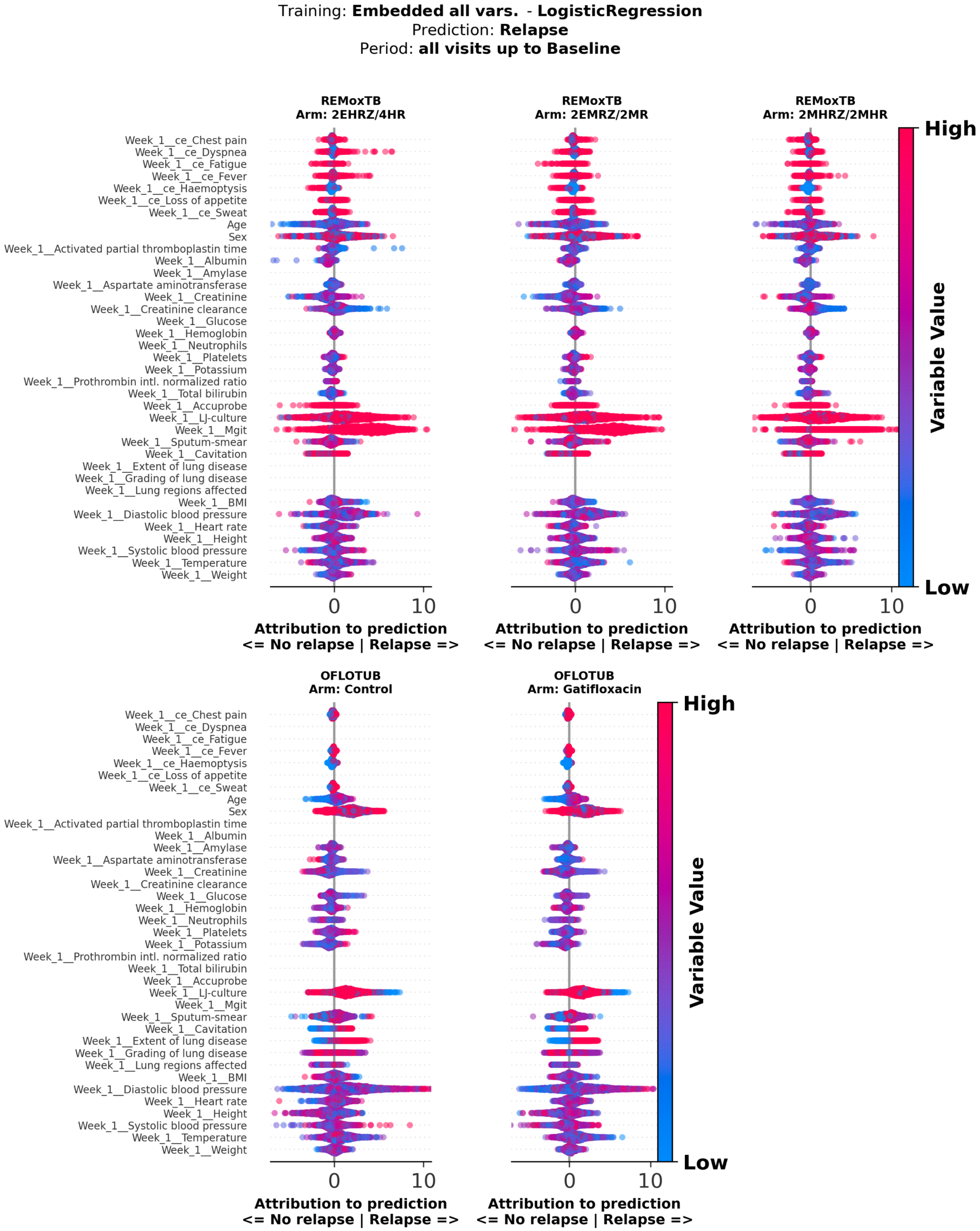


**Supplementary Figure 30**: **Attribution score analysis of logistic regresssion model predicting relapse, using embeddings of all variable data at baseline – variables with largest attributions.** Attribution scores of logistic regression models predicting relapse trained on embeddings of all clinical and treatment variables at baseline. Attribution scores were calculated on test set patients. The y-axis shows top 40 variables with largest mean absolute attribution scores; the x-axis shows attribution scores, where positive values indicate greater contribution to the positive label (Relapse), and negative values indicate lower contribution. Each dot represents a patient, with dot colour indicating the normalized value of the variable for that patient. Panels show patients stratified by treatment arm, with arm and study name indicated above each panel. ce: clinical event, BMI: body mass index, LJ: Lowenstein-Jensen, MGIT: Mycobacterium indicator growth tube

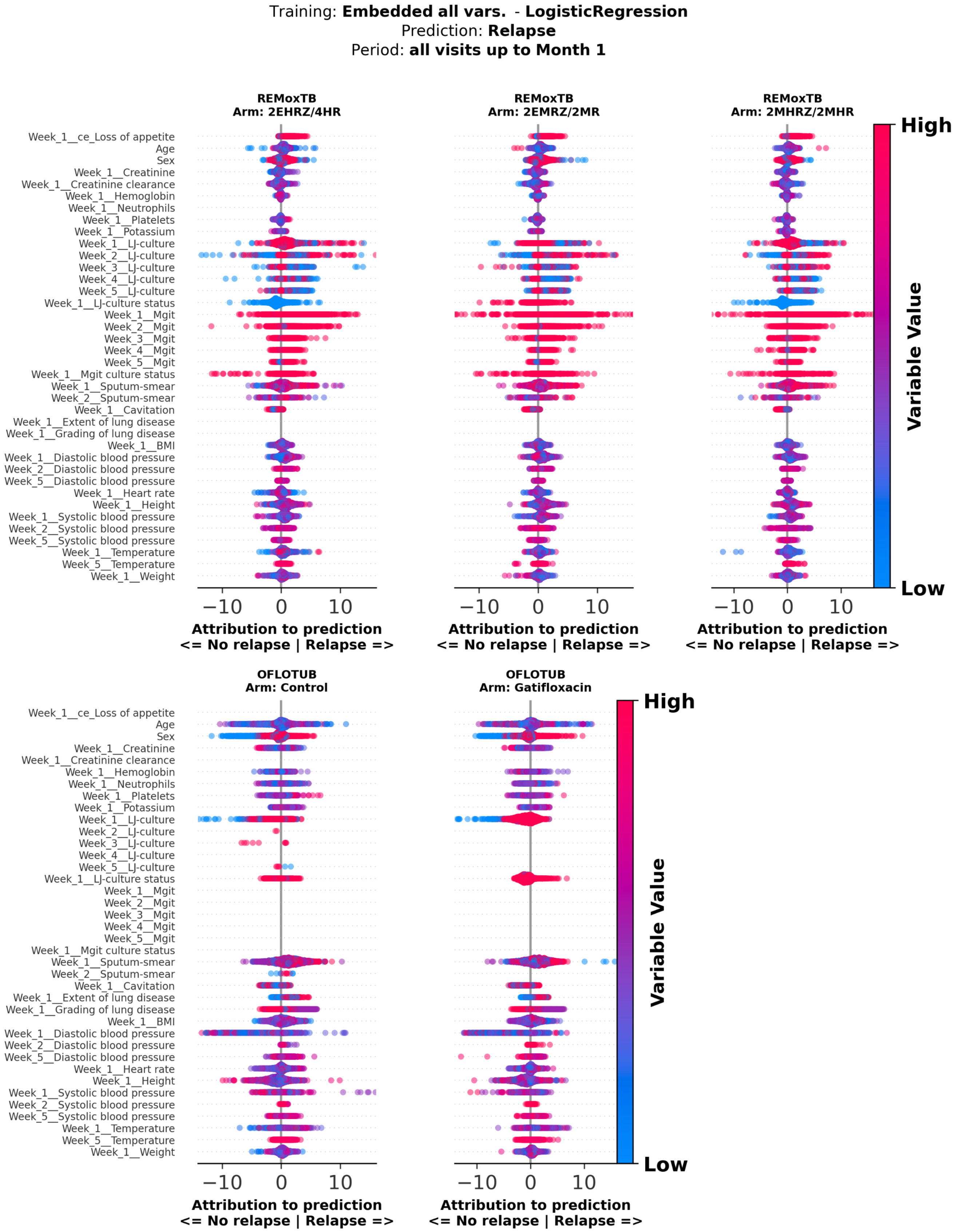


**Supplementary Figure 31**: **Attribution score analysis of logistic regresssion model predicting relapse, using embeddings of all variables and all visits up to month 1 – variables with largest attributions.** Attribution scores of logistic regression models predicting relapse trained on embeddings of all clinical and treatment variables, including all visits up to month 1. Attribution scores were calculated on test set patients. embeddings of all clinical and treatment variables at baseline. Attribution scores were calculated on test set patients. The y-axis shows top 40 variables with largest mean absolute attribution scores; the x-axis shows attribution scores, where positive values indicate greater contribution to the positive label (Relapse), and negative values indicate lower contribution. Each dot represents a patient, with dot colour indicating the normalized value of the variable for that patient. Panels show patients stratified by treatment arm, with arm and study name indicated above each panel. BMI: body mass index, LJ: Lowenstein-Jensen, MGIT: Mycobacterium indicator growth tube

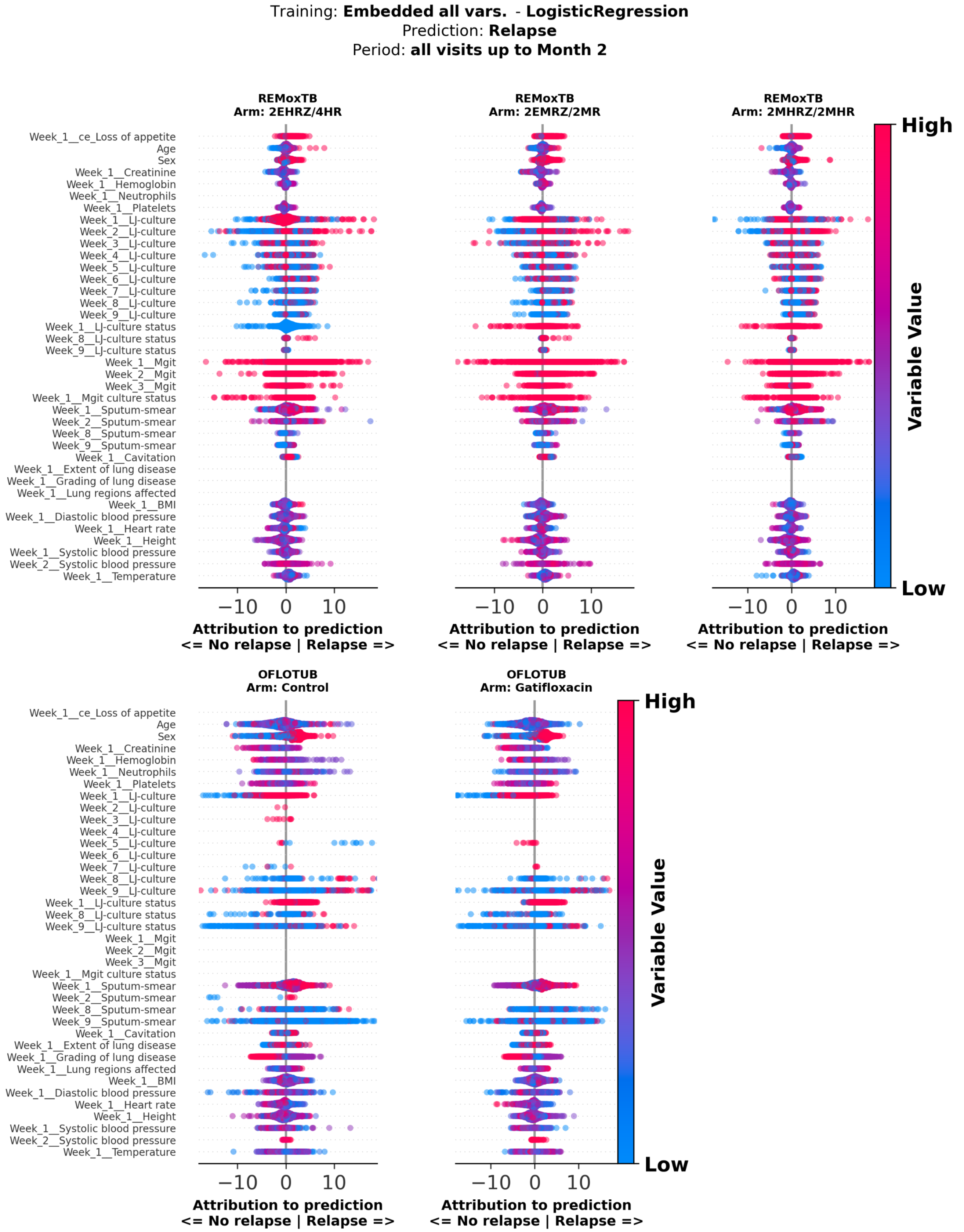


**Supplementary Figure 32**: **Attribution score analysis of logistic regresssion model predicting relapse, using embeddings of all variables and all visits up to month 2 – variables with largest attributions.** Attribution scores of logistic regression models predicting relapse trained on embeddings of all clinical and treatment variables, including all visits up to month 2. Attribution scores were calculated on test set patients. embeddings of all clinical and treatment variables at baseline. Attribution scores were calculated on test set patients. The y-axis shows top 40 variables with largest mean absolute attribution scores; the x-axis shows attribution scores, where positive values indicate greater contribution to the positive label (Relapse), and negative values indicate lower contribution. Each dot represents a patient, with dot colour indicating the normalized value of the variable for that patient. Panels show patients stratified by treatment arm, with arm and study name indicated above each panel. BMI: body mass index, LJ: Lowenstein-Jensen, MGIT: Mycobacterium indicator growth tube

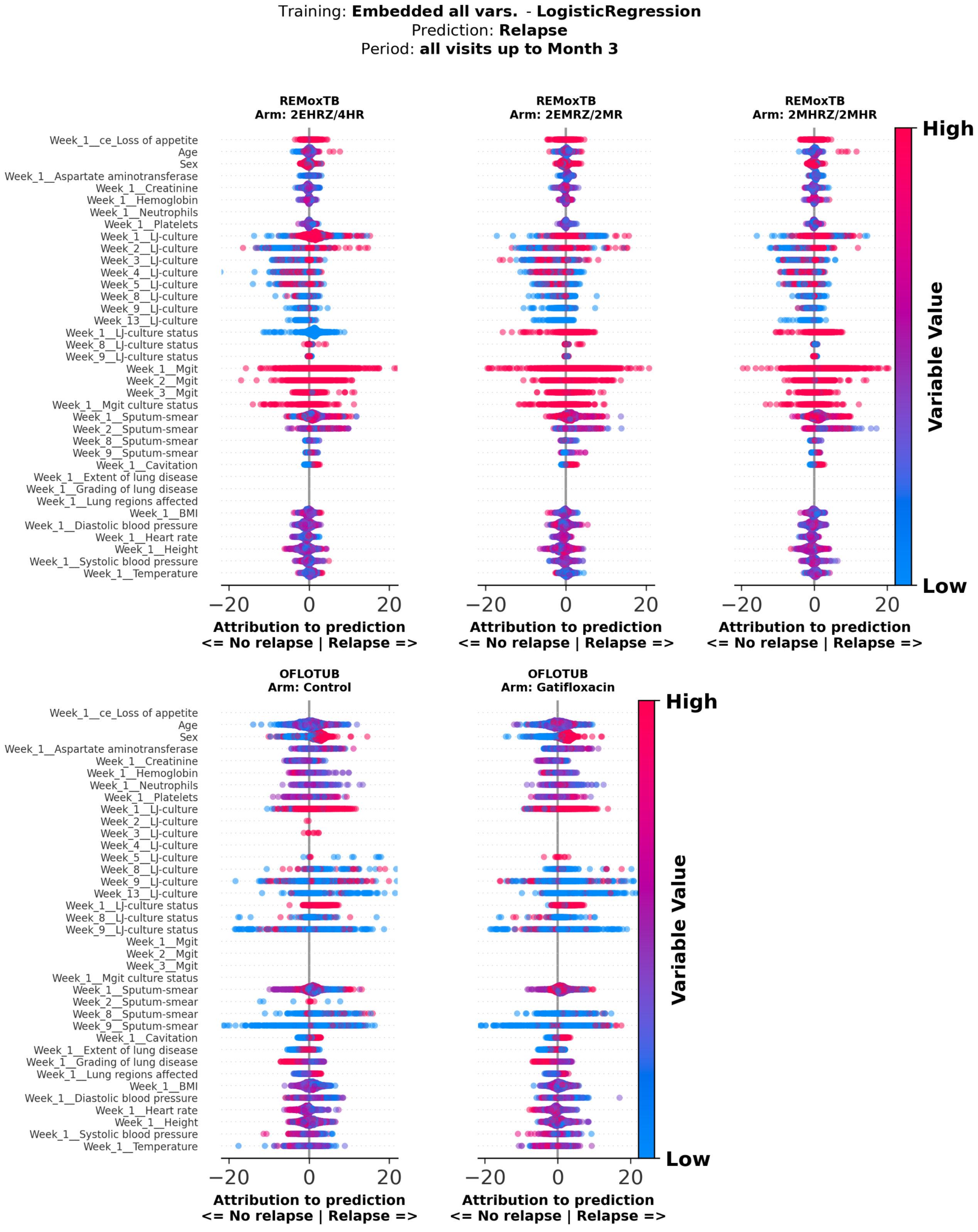


**Supplementary Figure 33**: **Attribution score analysis of logistic regresssion model predicting relapse, using embeddings of all variables and all visits up to month 3 – variables with largest attributions.** Attribution scores of logistic regression models predicting relapse trained on embeddings of all clinical and treatment variables, including all visits up to month 3. Attribution scores were calculated on test set patients. embeddings of all clinical and treatment variables at baseline. Attribution scores were calculated on test set patients. The y-axis shows top 40 variables with largest mean absolute attribution scores; the x-axis shows attribution scores, where positive values indicate greater contribution to the positive label (Relapse), and negative values indicate lower contribution. Each dot represents a patient, with dot colour indicating the normalized value of the variable for that patient. Panels show patients stratified by treatment arm, with arm and study name indicated above each panel. BMI: body mass index, LJ: Lowenstein-Jensen, MGIT: Mycobacterium indicator growth tube

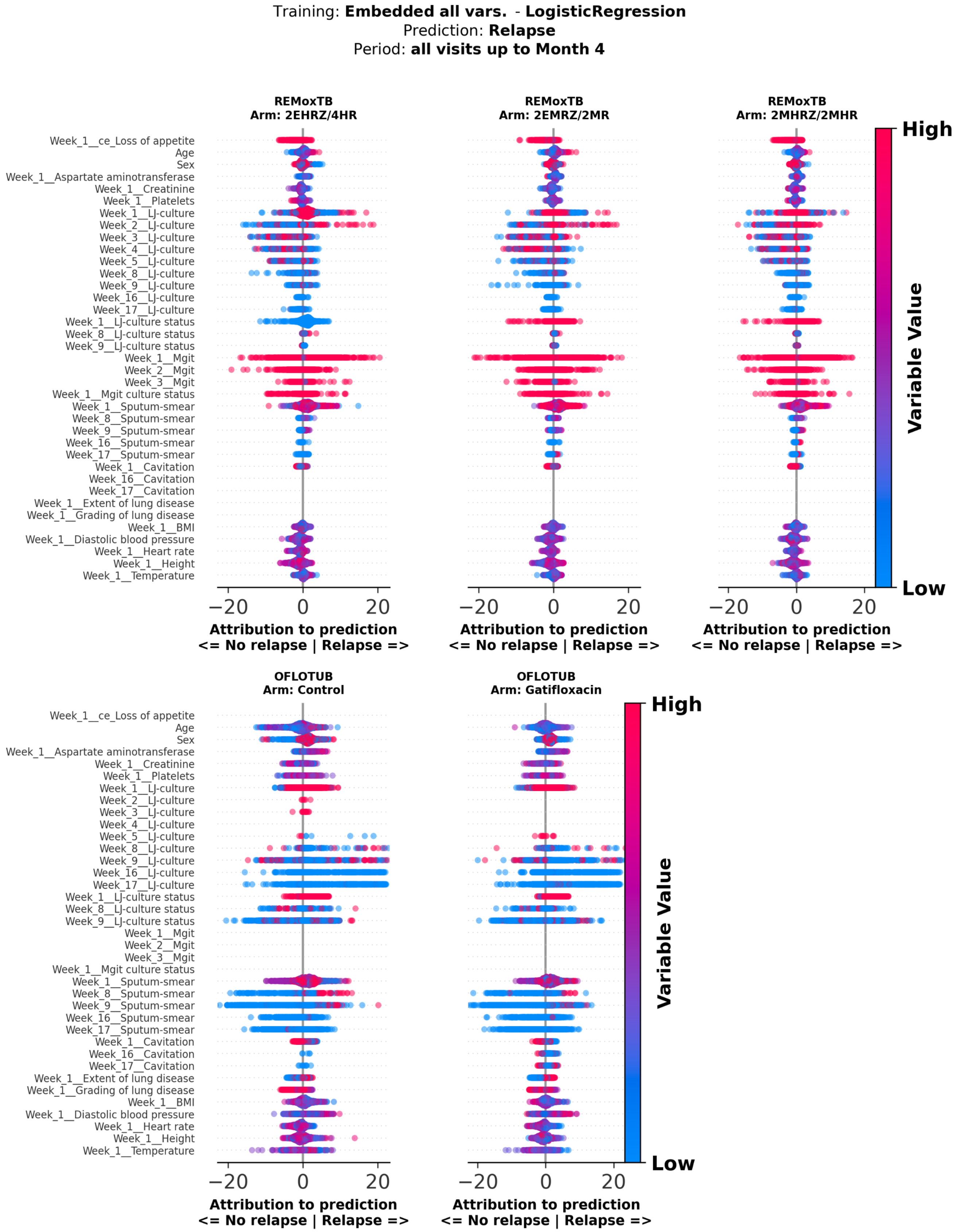


**Supplementary Figure 34**: **Attribution score analysis of logistic regresssion model predicting relapse, using embeddings of all variables and all visits up to month 4 – variables with largest attributions.** Attribution scores of logistic regression models predicting relapse trained on embeddings of all clinical and treatment variables, including all visits up to month 4. Attribution scores were calculated on test set patients. embeddings of all clinical and treatment variables at baseline. Attribution scores were calculated on test set patients. The y-axis shows top 40 variables with largest mean absolute attribution scores; the x-axis shows attribution scores, where positive values indicate greater contribution to the positive label (Relapse), and negative values indicate lower contribution. Each dot represents a patient, with dot colour indicating the normalized value of the variable for that patient. Panels show patients stratified by treatment arm, with arm and study name indicated above each panel. BMI: body mass index, LJ: Lowenstein-Jensen, MGIT: Mycobacterium indicator growth tube

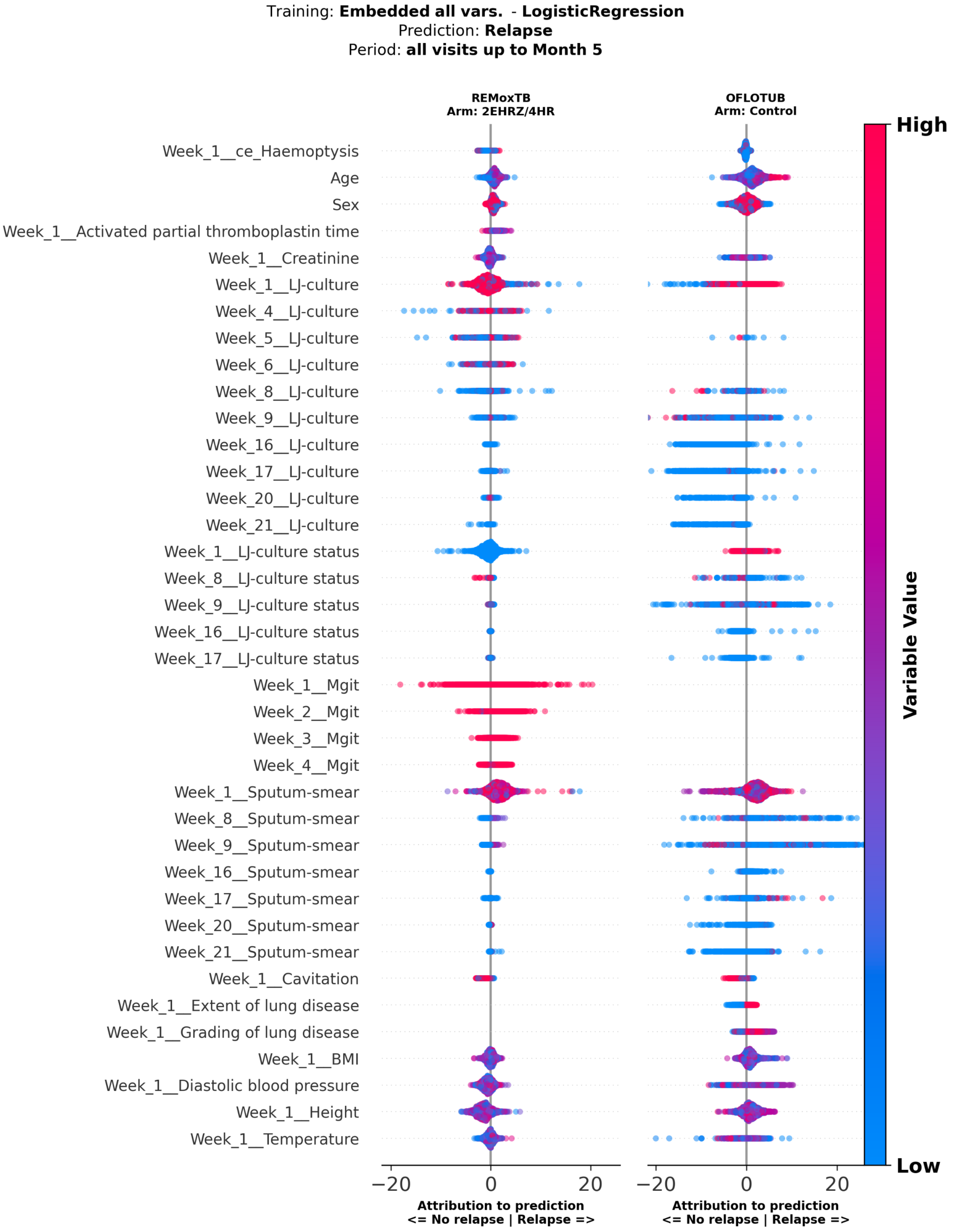


**Supplementary Figure 35**: **Attribution score analysis of logistic regresssion model predicting relapse, using embeddings of all variables and all visits up to month 5 – variables with largest attributions.** Attribution scores of logistic regression models predicting relapse trained on embeddings of all clinical and treatment variables, including all visits up to month 5. Attribution scores were calculated on test set patients. embeddings of all clinical and treatment variables at baseline. Attribution scores were calculated on test set patients. The y-axis shows top 40 variables with largest mean absolute attribution scores; the x-axis shows attribution scores, where positive values indicate greater contribution to the positive label (Relapse), and negative values indicate lower contribution. Each dot represents a patient, with dot colour indicating the normalized value of the variable for that patient. Panels show patients stratified by treatment arm, with arm and study name indicated above each panel. BMI: body mass index, LJ: Lowenstein-Jensen, MGIT: Mycobacterium indicator growth tube

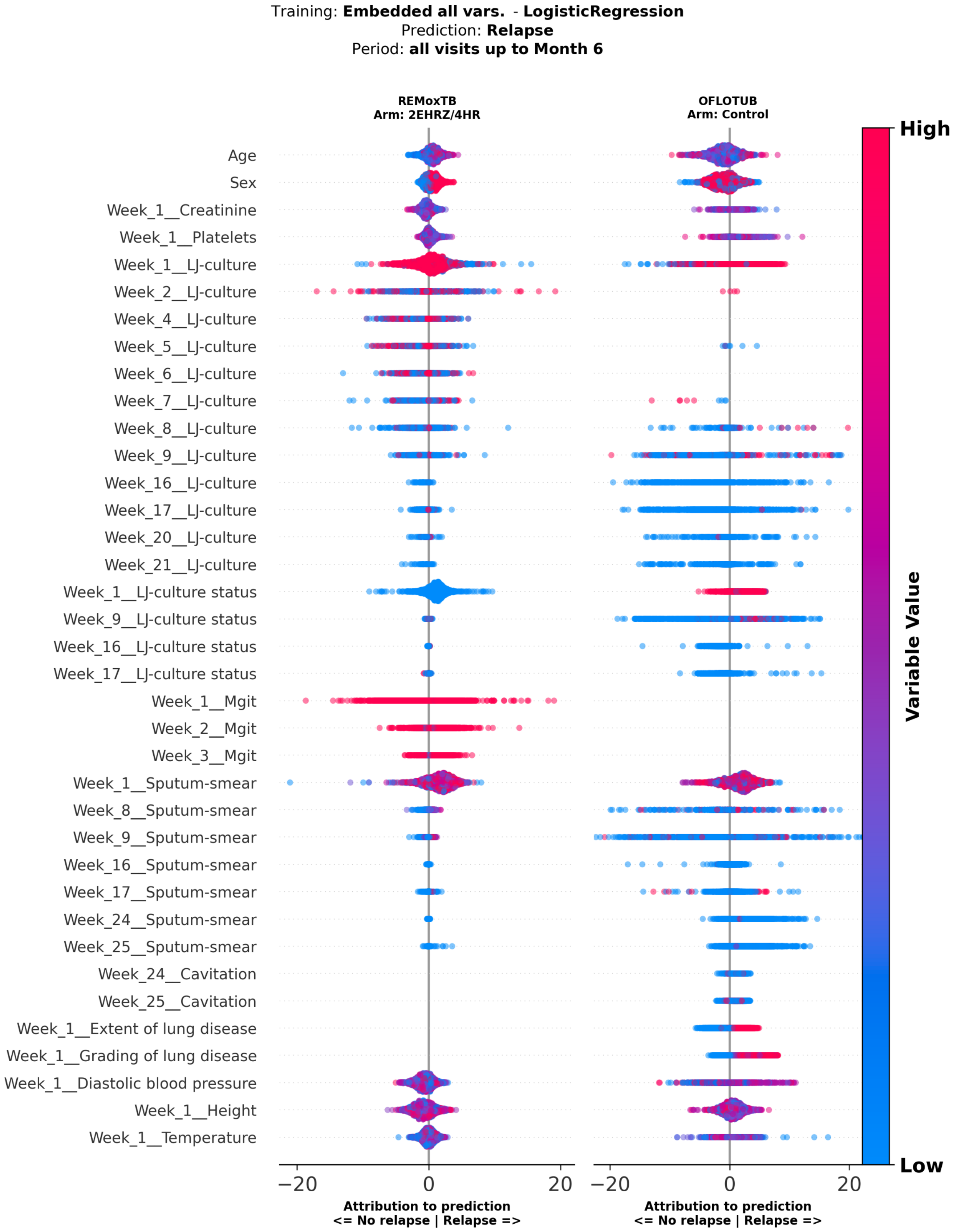


**Supplementary Figure 36**: **Attribution score analysis of logistic regresssion model predicting relapse, using embeddings of all variables and all visits up to month 6 – variables with largest attributions.** Attribution scores of logistic regression models predicting relapse trained on embeddings of all clinical and treatment variables, including all visits up to month 6. Attribution scores were calculated on test set patients. embeddings of all clinical and treatment variables at baseline. Attribution scores were calculated on test set patients. The y-axis shows top 40 variables with largest mean absolute attribution scores; the x-axis shows attribution scores, where positive values indicate greater contribution to the positive label (Relapse), and negative values indicate lower contribution. Each dot represents a patient, with dot colour indicating the normalized value of the variable for that patient. Panels show patients stratified by treatment arm, with arm and study name indicated above each panel. BMI: body mass index, LJ: Lowenstein-Jensen, MGIT: Mycobacterium indicator growth tube

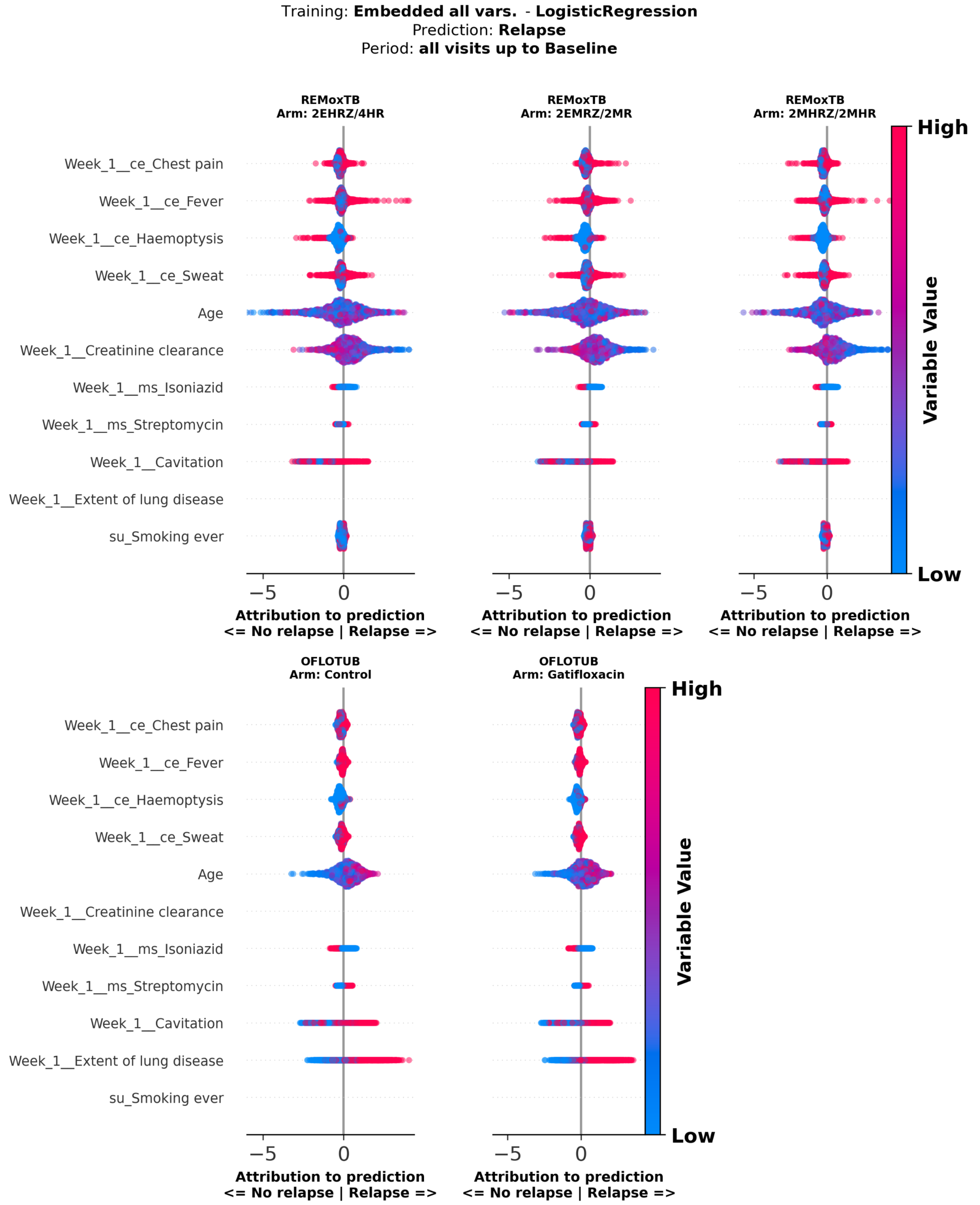


**Supplementary Figure 37**: **Attribution score analysis of logistic regresssion model predicting relapse, using embeddings of all variables at baseline – variables with largest correlations.** Attribution scores of logistic regression models predicting relapse trained on embeddings of all clinical and treatment variables at baseline. Attribution scores were calculated on test set patients. embeddings of all clinical and treatment variables at baseline. Attribution scores were calculated on test set patients. The y-axis shows top 40 variables with largest mean correlations between variable values and attribution scores; the x-axis shows attribution scores, where positive values indicate greater contribution to the positive label (Relapse), and negative values indicate lower contribution. Each dot represents a patient, with dot colour indicating the normalized value of the variable for that patient. Panels show patients stratified by treatment arm, with arm and study name indicated above each panel. mh.: medical history; ce: clinical event, ms: microbiological susceptibility test, su: substance use, LJ: Lowenstein-Jensen.

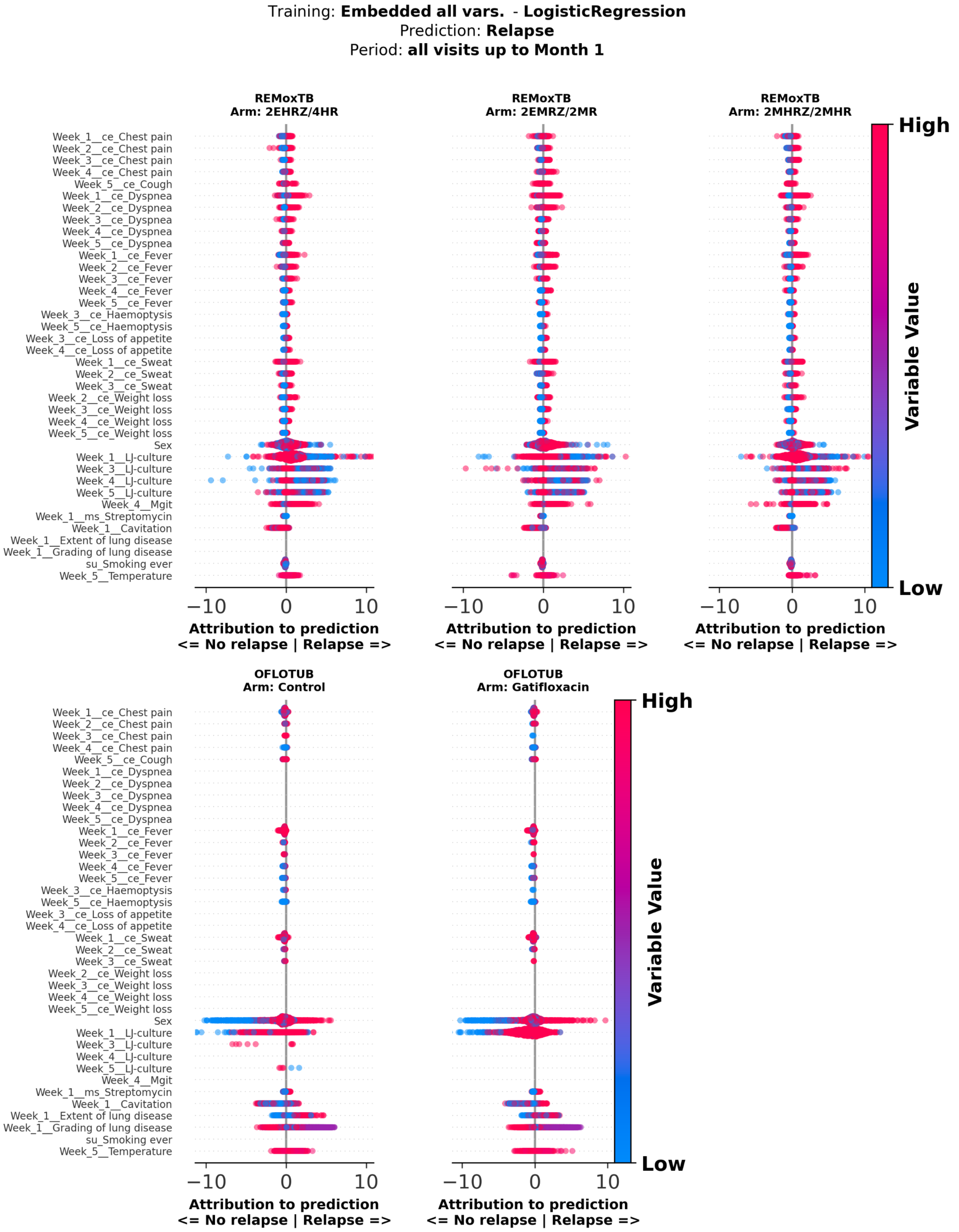


**Supplementary Figure 38**: **Attribution score analysis of logistic regresssion model predicting relapse, using embeddings of all variables and all visits up to month 1 – variables with largest correlations.** Attribution scores of logistic regression models predicting relapse trained on embeddings of all clinical and treatment variables, including all visits up to month 1. Attribution scores were calculated on test set patients. embeddings of all clinical and treatment variables at baseline. Attribution scores were calculated on test set patients. The y-axis shows top 40 variables with largest mean correlations between variable values and attribution scores; the x-axis shows attribution scores, where positive values indicate greater contribution to the positive label (Relapse), and negative values indicate lower contribution. Each dot represents a patient, with dot colour indicating the normalized value of the variable for that patient. Panels show patients stratified by treatment arm, with arm and study name indicated above each panel. mh.: medical history, ce: clinical event, ms: microbiological susceptibility test, su: substance use, cumul.: cumulative

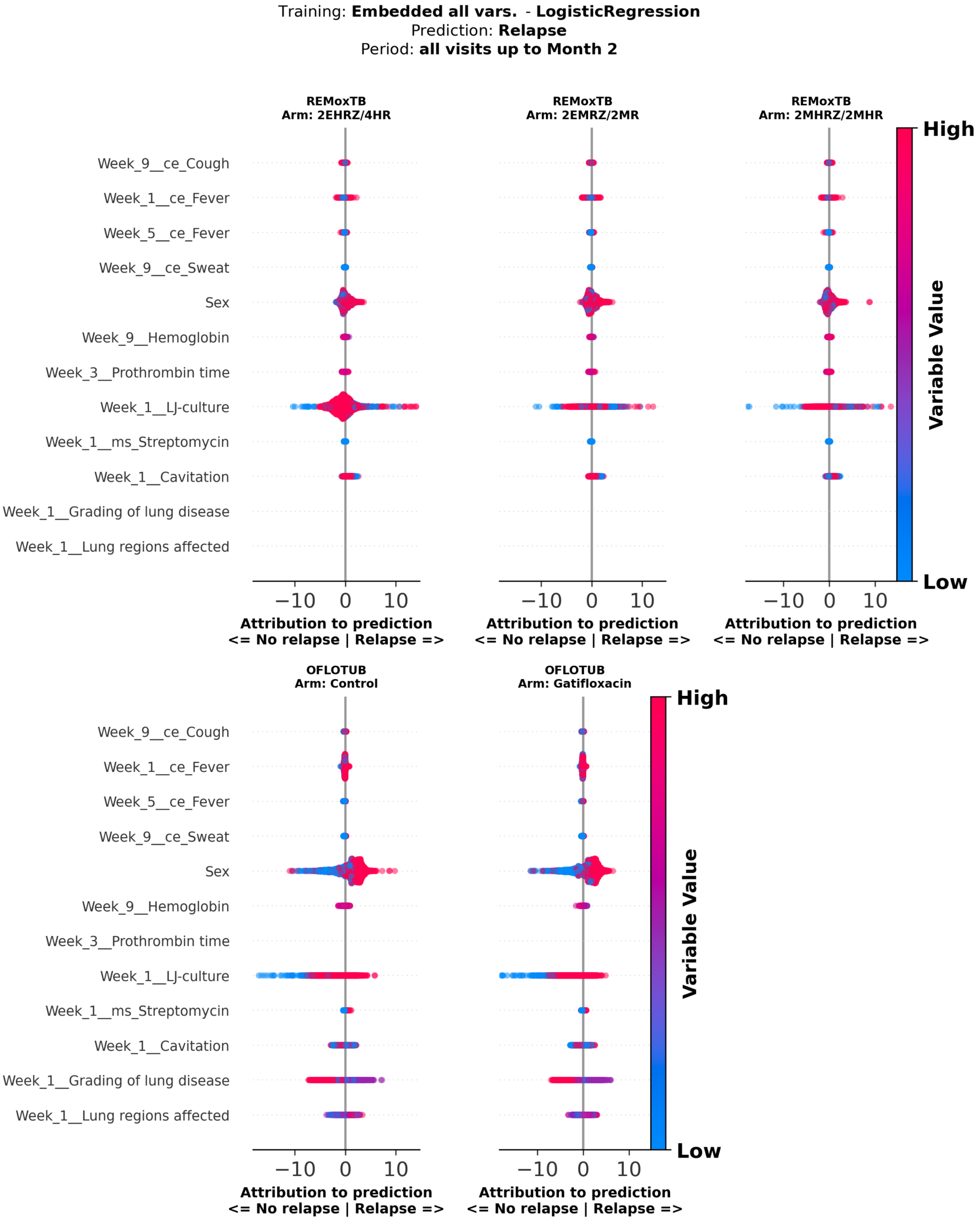


**Supplementary Figure 39**: **Attribution score analysis of logistic regresssion model predicting relapse, using embeddings of all variables and all visits up to month 2 – variables with largest correlations.** Attribution scores of logistic regression models predicting relapse trained on embeddings of all clinical and treatment variables, including all visits up to month 2. Attribution scores were calculated on test set patients. embeddings of all clinical and treatment variables at baseline. Attribution scores were calculated on test set patients. The y-axis shows top 40 variables with largest mean correlations between variable values and attribution scores; the x-axis shows attribution scores, where positive values indicate greater contribution to the positive label (Relapse), and negative values indicate lower contribution. Each dot represents a patient, with dot colour indicating the normalized value of the variable for that patient. Panels show patients stratified by treatment arm, with arm and study name indicated above each panel. mh.: medical history, ms: microbiological susceptibility test, cumul.: cumulative, LJ: Lowenstein-Jensen

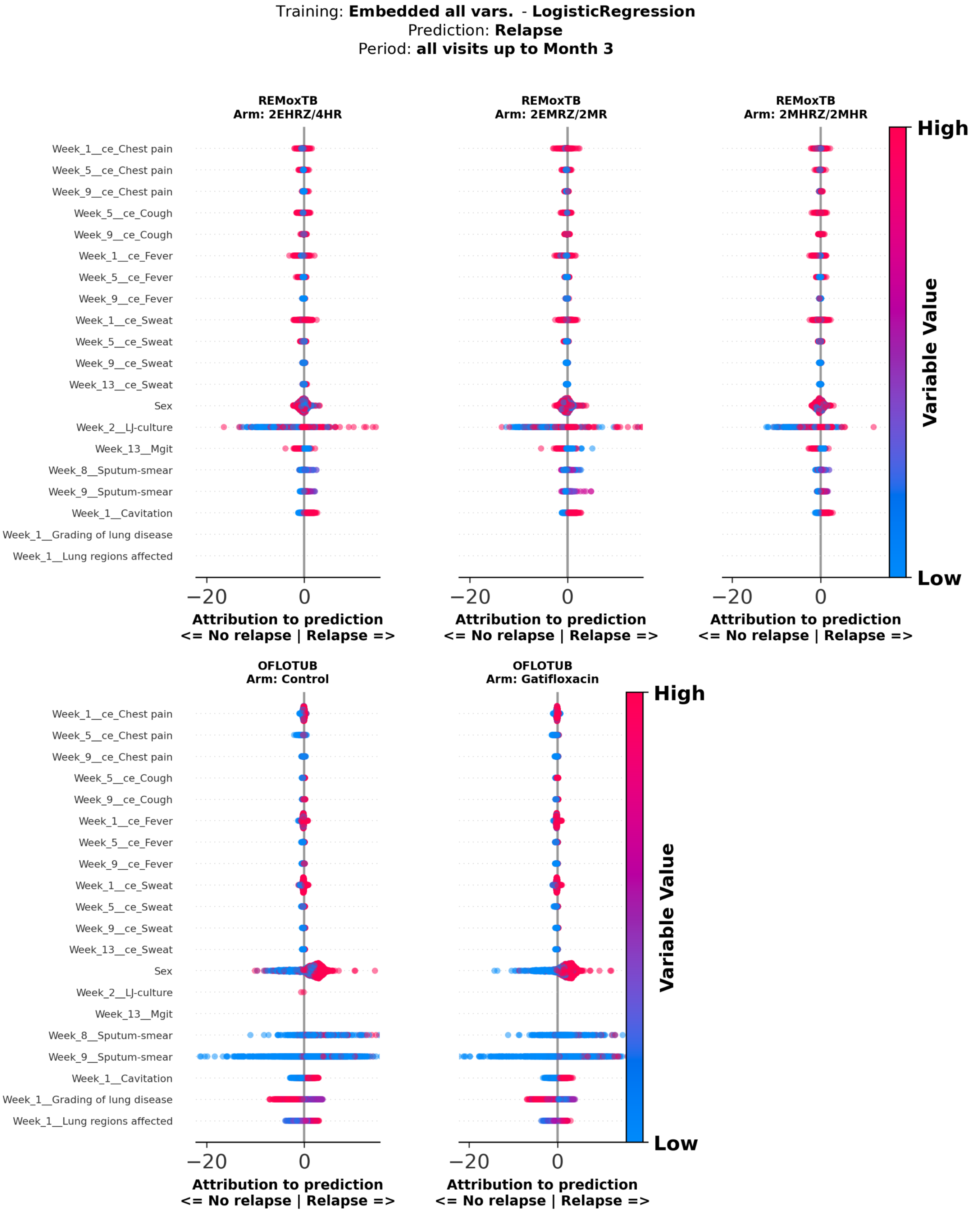


**Supplementary Figure 40**: **Attribution score analysis of logistic regresssion model predicting relapse, using embeddings of all variables and all visits up to month 3 – variables with largest correlations.** Attribution scores of logistic regression models predicting relapse trained on embeddings of all clinical and treatment variables, including all visits up to month 3. Attribution scores were calculated on test set patients. embeddings of all clinical and treatment variables at baseline. Attribution scores were calculated on test set patients. The y-axis shows top 40 variables with largest mean correlations between variable values and attribution scores; the x-axis shows attribution scores, where positive values indicate greater contribution to the positive label (Relapse), and negative values indicate lower contribution. Each dot represents a patient, with dot colour indicating the normalized value of the variable for that patient. Panels show patients stratified by treatment arm, with arm and study name indicated above each panel. mh.: medical history, ce: clinical event, ms: microbiological susceptibility test, cumul.: cumulative, LJ: Lowenstein-Jensen, MGIT: Mycobacterium indicator growth tube

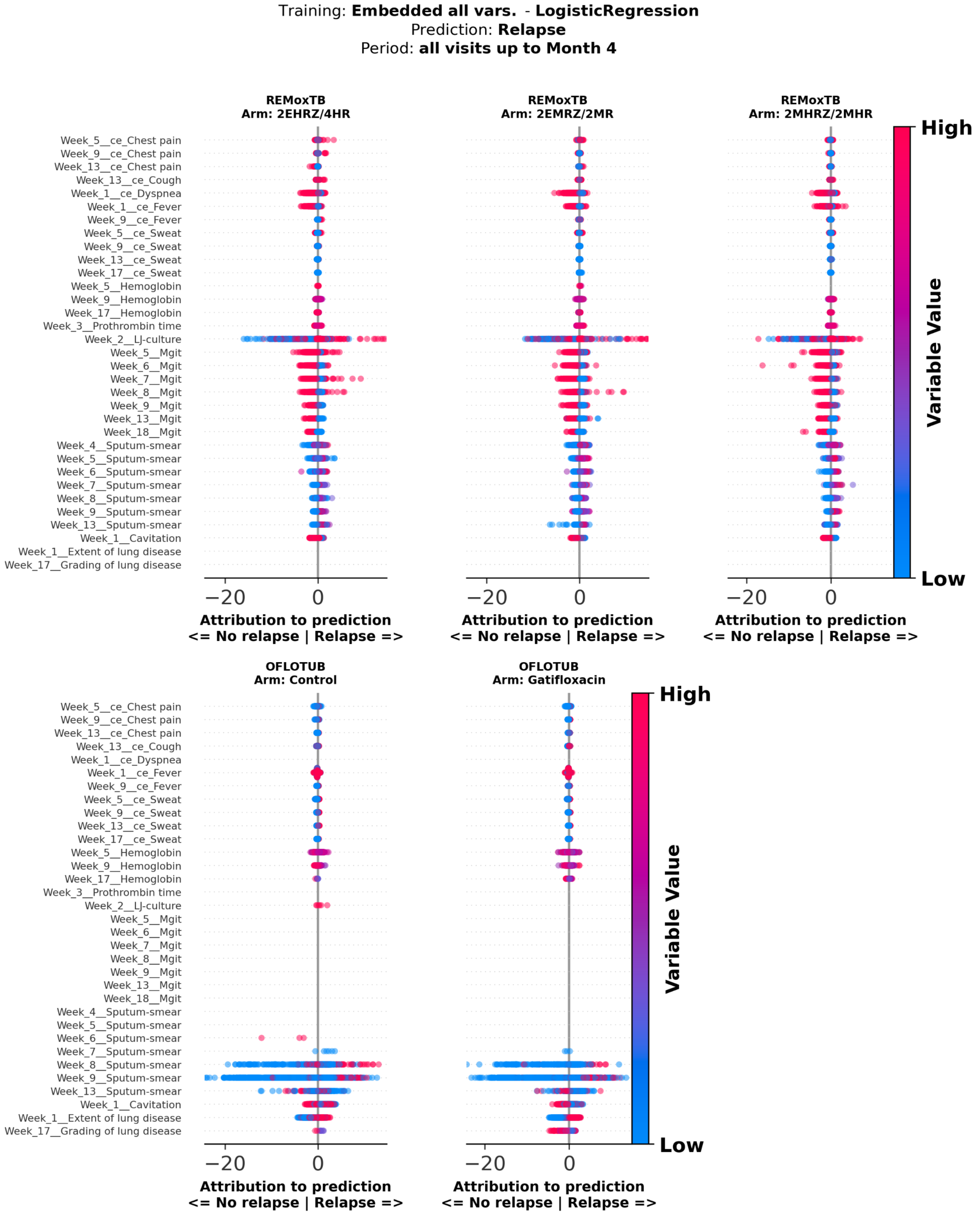


**Supplementary Figure 41**: **Attribution score analysis of logistic regresssion model predicting relapse, using embeddings of all variables and all visits up to month 4 – variables with largest correlations.** Attribution scores of logistic regression models predicting relapse trained on embeddings of all clinical and treatment variables, including all visits up to month 4. Attribution scores were calculated on test set patients. embeddings of all clinical and treatment variables at baseline. Attribution scores were calculated on test set patients. The y-axis shows top 40 variables with largest mean correlations between variable values and attribution scores; the x-axis shows attribution scores, where positive values indicate greater contribution to the positive label (Relapse), and negative values indicate lower contribution. Each dot represents a patient, with dot colour indicating the normalized value of the variable for that patient. Panels show patients stratified by treatment arm, with arm and study name indicated above each panel. mh.: medical history, ce: clinical event, cumul.: cumulative, LJ: Lowenstein-Jensen, MGIT: Mycobacterium indicator growth tube

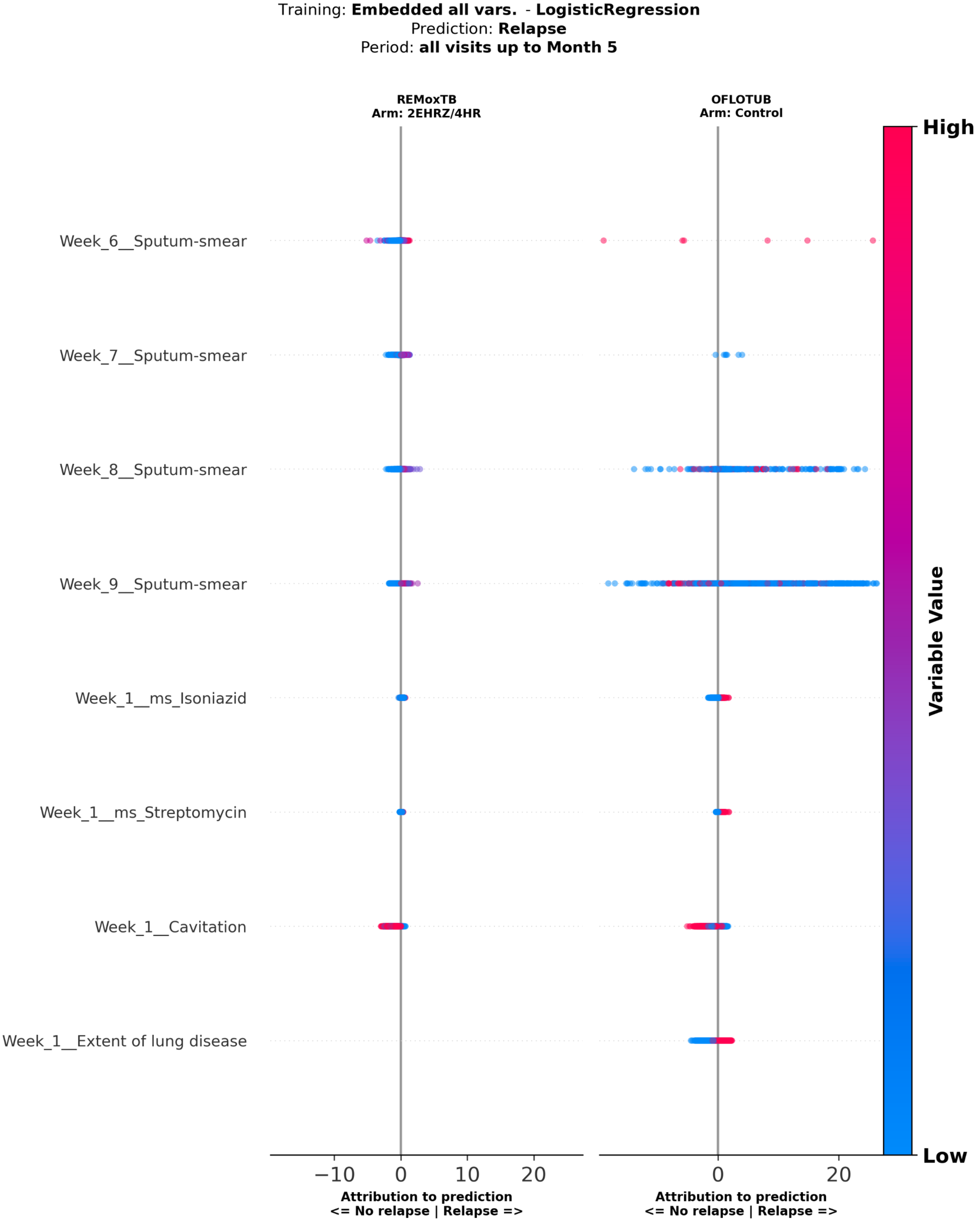


**Supplementary Figure 42**: **Attribution score analysis of logistic regresssion model predicting relapse, using embeddings of all variables and all visits up to month 5 – variables with largest correlations.** Attribution scores of logistic regression models predicting relapse trained on embeddings of all clinical and treatment variables, including all visits up to month 5. Attribution scores were calculated on test set patients. embeddings of all clinical and treatment variables at baseline. Attribution scores were calculated on test set patients. The y-axis shows top 40 variables with largest mean correlations between variable values and attribution scores; the x-axis shows attribution scores, where positive values indicate greater contribution to the positive label (Relapse), and negative values indicate lower contribution. Each dot represents a patient, with dot colour indicating the normalized value of the variable for that patient. Panels show patients stratified by treatment arm, with arm and study name indicated above each panel. mh.: medical history, ce: clinical event, ms: microbiological susceptibility test, cumul.: cumulative

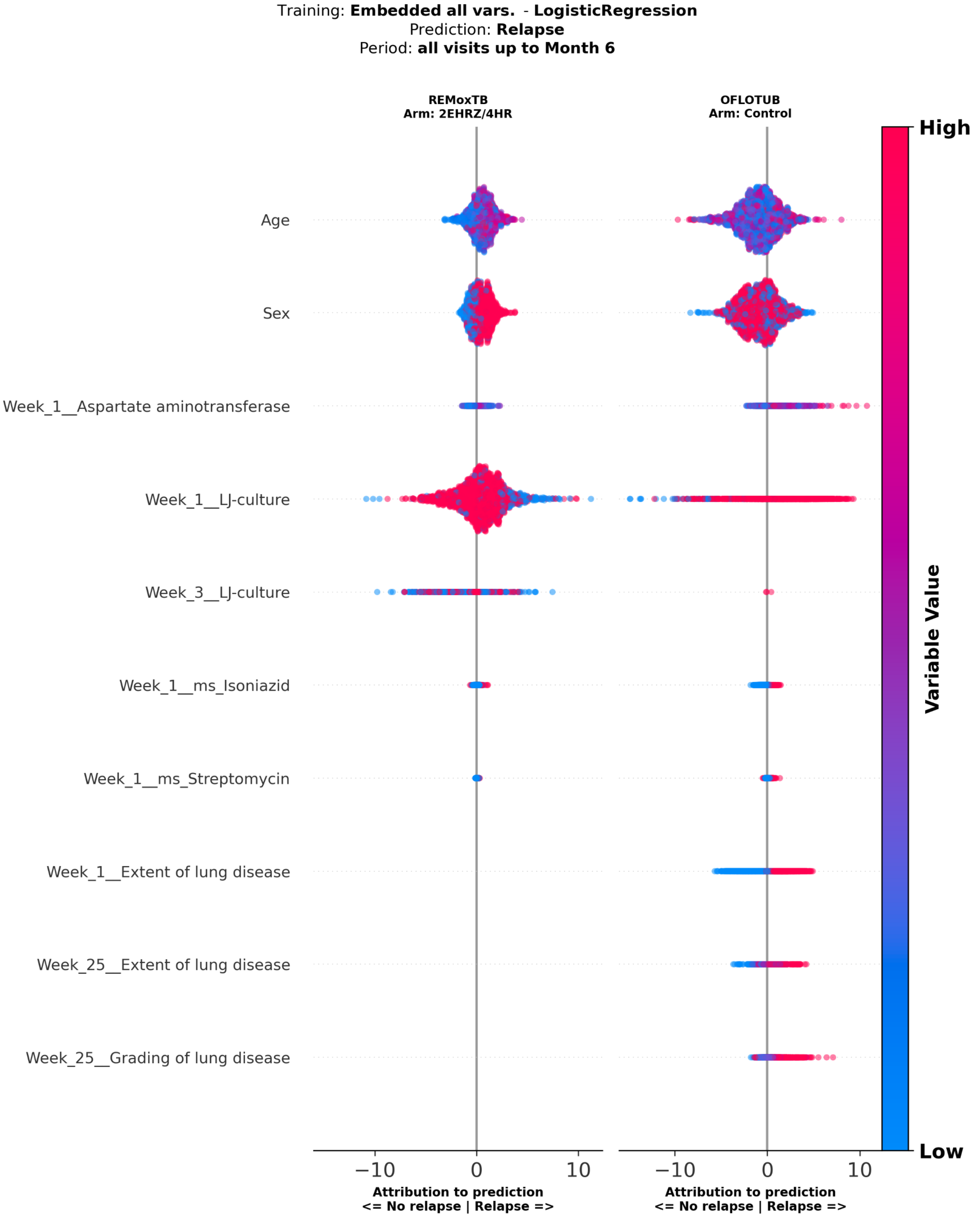


**Supplementary Figure 43**: **Attribution score analysis of logistic regresssion model predicting relapse, using embeddings of all variables and all visits up to month 6 – variables with largest correlations.** Attribution scores of logistic regression models predicting relapse trained on embeddings of all clinical and treatment variables, including all visits up to month 6. Attribution scores were calculated on test set patients. embeddings of all clinical and treatment variables at baseline. Attribution scores were calculated on test set patients. The y-axis shows top 40 variables with largest mean correlations between variable values and attribution scores; the x-axis shows attribution scores, where positive values indicate greater contribution to the positive label (Relapse), and negative values indicate lower contribution. Each dot represents a patient, with dot colour indicating the normalized value of the variable for that patient. Panels show patients stratified by treatment arm, with arm and study name indicated above each panel. ms: microbiological susceptibility test, cumul.: cumulative, LJ: Lowenstein-Jensen

A

**TB severity (baseline Cavitation & Sputum-smear)**

**4-month cohort**

Number at risk

| Months since end-of-therapy | 0 | 3 | 6 | 9 | 12 |
|---|---|---|---|---|---|
| Easy-to-treat | 721 | 715 | 679 | 663 | 659 |
| Hard-to-treat | 426 | 400 | 370 | 359 | 353 |

Cumulative number of relapses

| Months since end-of-therapy | 0 | 3 | 6 | 9 | 12 |
|---|---|---|---|---|---|
| Easy-to-treat | 0 | 6 | 42 | 58 | 62 |
| Hard-to-treat | 0 | 27 | 56 | 67 | 73 |

**6-month cohort**

Number at risk

| Months since end-of-therapy | 0 | 3 | 6 | 9 | 12 |
|---|---|---|---|---|---|
| Easy-to-treat | 552 | 550 | 542 | 539 | 419 |
| Hard-to-treat | 240 | 234 | 231 | 227 | 134 |

Cumulative number of relapses

| Months since end-of-therapy | 0 | 3 | 6 | 9 | 12 |
|---|---|---|---|---|---|
| Easy-to-treat | 0 | 1 | 9 | 12 | 19 |
| Hard-to-treat | 0 | 6 | 9 | 13 | 14 |

B

**Raw common vars. - last visit in period - LR**

**4-month cohort**

Number at risk

| Months since end-of-therapy | 0 | 3 | 6 | 9 | 12 |
|---|---|---|---|---|---|
| Low | 442 | 435 | 427 | 421 | 419 |
| High | 452 | 426 | 378 | 363 | 355 |

Cumulative number of relapses

| Months since end-of-therapy | 0 | 3 | 6 | 9 | 12 |
|---|---|---|---|---|---|
| Low | 0 | 7 | 15 | 21 | 23 |
| High | 0 | 26 | 74 | 89 | 97 |

**6-month cohort**

Number at risk

| Months since end-of-therapy | 0 | 3 | 6 | 9 | 12 |
|---|---|---|---|---|---|
| Low | 318 | 315 | 313 | 311 | 229 |
| High | 280 | 276 | 271 | 266 | 158 |

Cumulative number of relapses

| Months since end-of-therapy | 0 | 3 | 6 | 9 | 12 |
|---|---|---|---|---|---|
| Low | 0 | 2 | 4 | 6 | 7 |
| High | 0 | 4 | 9 | 14 | 17 |

C

**Embedding model: all vars. - all visits in period - LR**

**4-month cohort**

Number at risk

| Months since end-of-therapy | 0 | 3 | 6 | 9 | 12 |
|---|---|---|---|---|---|
| Low | 393 | 389 | 381 | 378 | 375 |
| High | 487 | 458 | 407 | 391 | 384 |

Cumulative number of relapses

| Months since end-of-therapy | 0 | 3 | 6 | 9 | 12 |
|---|---|---|---|---|---|
| Low | 0 | 4 | 12 | 15 | 18 |
| High | 0 | 30 | 80 | 96 | 103 |

**6-month cohort**

Number at risk

| Months since end-of-therapy | 0 | 3 | 6 | 9 | 12 |
|---|---|---|---|---|---|
| Low | 296 | 293 | 292 | 292 | 208 |
| High | 278 | 271 | 265 | 259 | 169 |

Cumulative number of relapses

| Months since end-of-therapy | 0 | 3 | 6 | 9 | 12 |
|---|---|---|---|---|---|
| Low | 0 | 3 | 4 | 4 | 5 |
| High | 0 | 6 | 12 | 18 | 21 |

**Supplementary Figure 44**: **Risk stratification of post-treatment tuberculosis relapse: number at risk and cumulative relapse counts across models and treatment durations.** Tables summarising the number of patients at risk and cumulative number of relapses over time following end-of-therapy (EOT), stratified by treatment duration and subgroup classification approach. Subgroups were defined as easy- versus hard-to-treat according to Imperial et al. (A), or as low- versus high-risk based on predictions from raw last-visit models (B), or LLM-based models incorporating all variables across all visits (C). Results are shown separately for the 4-month (left) and 6-month (right) cohorts at 0, 3, 6, 9, and 12 months after EOT. LR: logistic regression.

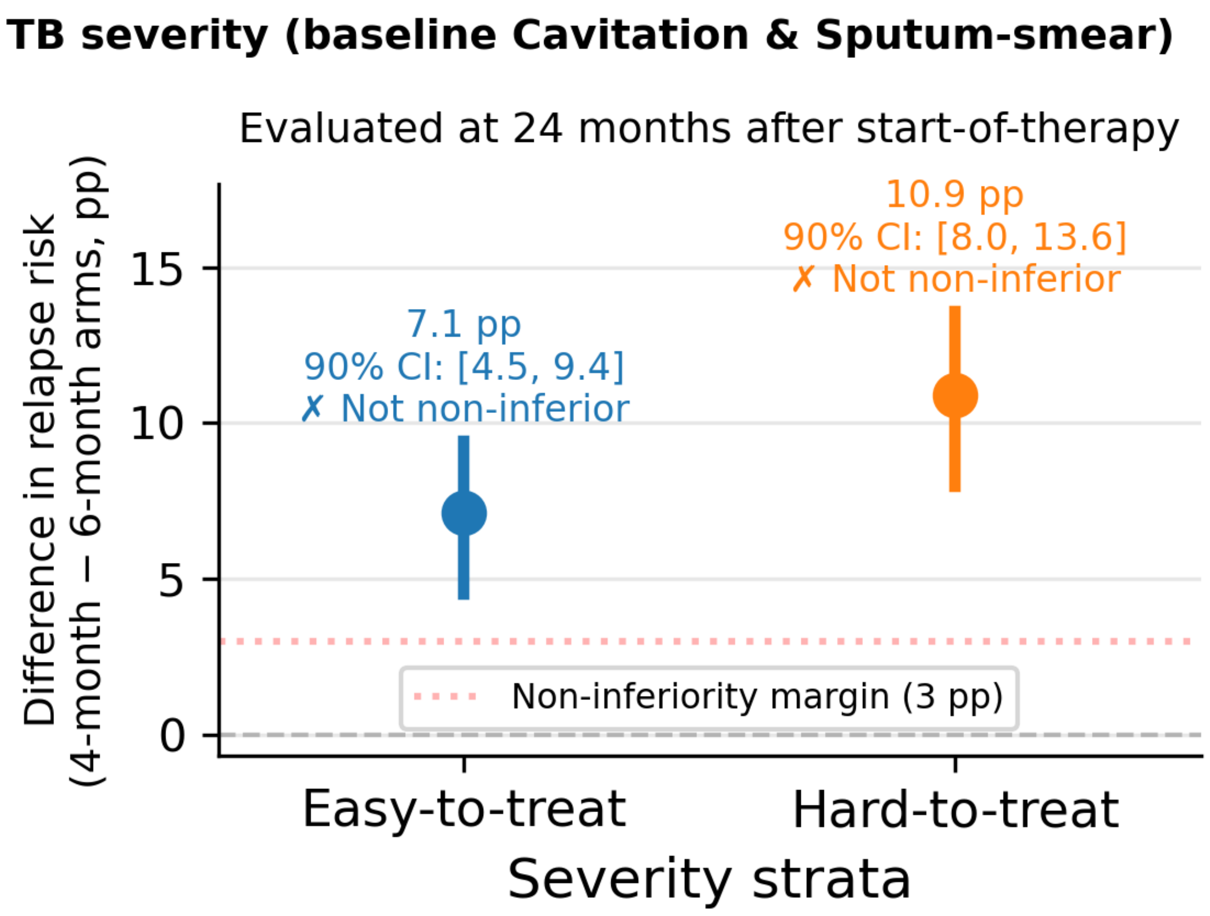


**Supplementary Figure 45**: **Non-inferiority analysis of relapse risk by tuberculosis severity strata at 24 months after start-of-treatment.** Inverse probability-of-study weighted estimates of the difference in relapse risk between the 4-month (experimental) and 6-month (control) treatment arms, evaluated at 24 months after start of therapy. Results are shown stratified by tuberculosis severity proposed by Imperial et al. (easy-to-treat and hard-to-treat), defined based on baseline cavitation and sputum smear status. Points represent the estimated absolute difference in relapse risk (percentage points, pp), calculated as the difference in inverse probability-of-study weighted Kaplan-Meier estimated relapse-free survival between treatment arms (4-month minus 6-month). Vertical bars indicate two-sided 90% confidence intervals derived from stratified bootstrap resampling (n=500), preserving study- and therapy duration level structure. The horizontal dashed line at 0 indicates no difference between treatment arms, and the dotted line represents the pre-specified non-inferiority margin of 3 percentage points. Non-inferiority is concluded if the upper bound of the 90% confidence interval lies below this margin. CI: confidence interval.

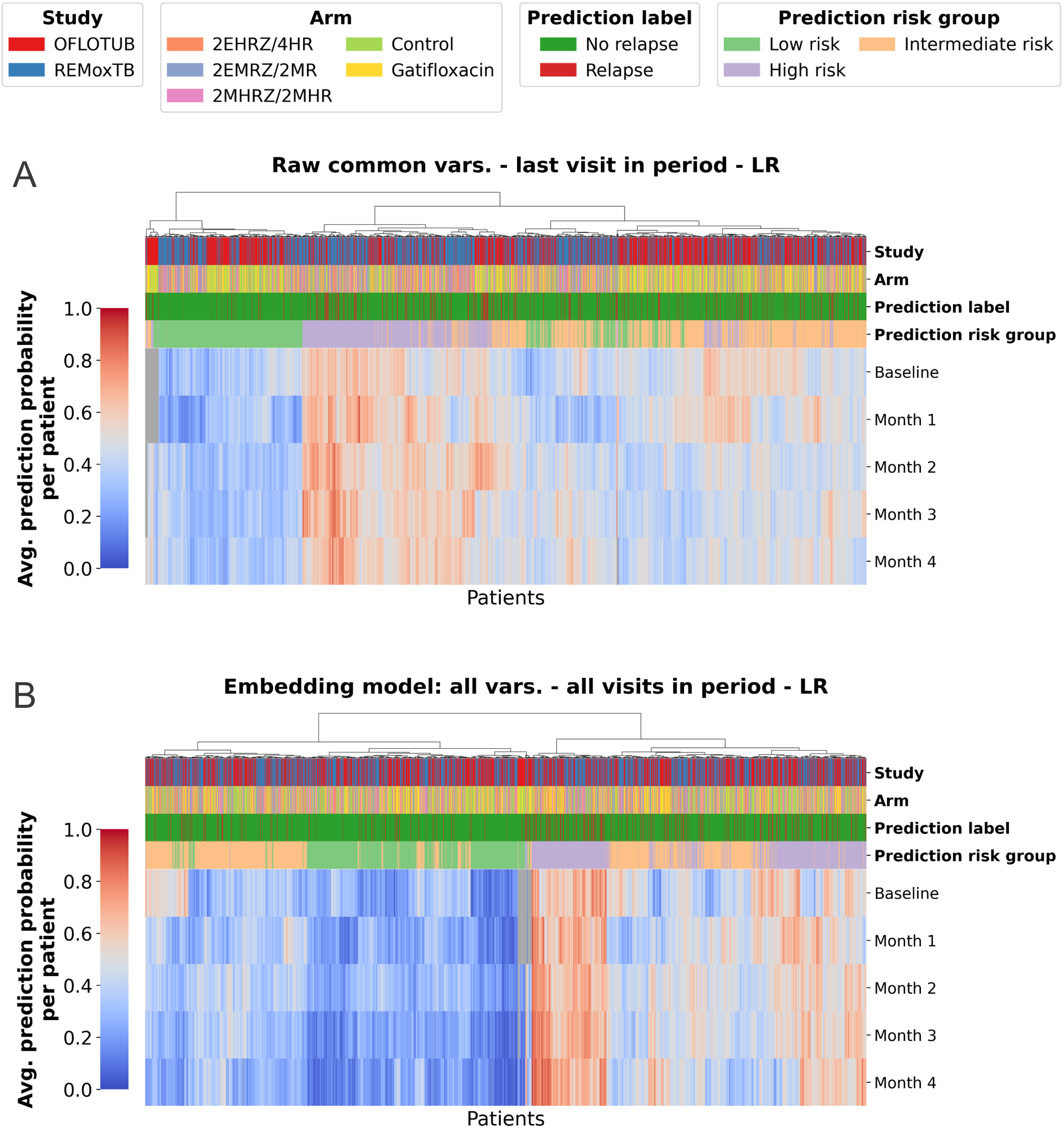


**Supplementary Figure 46**: **Relapse prediction probability of test-set patients across time periods.** (A) Heatmap of patient-specific average prediction probability across treatment timepoints for relapse prediction using raw tabular clinical variables (common variables, last visit in period) with a logistic regression model. Each column represents a patient, ordered by hierarchical clustering of prediction probability trajectories, and rows correspond to baseline and monthly treatment cutoffs. Annotation tracks indicate study (REMoxTB, OFLOTUB), treatment arm, true relapse outcome. (B) Corresponding heatmap for an embedding-based model using LLM-based representations constructed from all available variables across all visits within each period
